# The penetrance of rare variants in cardiomyopathy-associated genes: a cross-sectional approach to estimate penetrance for secondary findings

**DOI:** 10.1101/2023.03.15.23287112

**Authors:** Kathryn A. McGurk, Xiaolei Zhang, Pantazis Theotokis, Kate Thomson, Andrew Harper, Rachel J. Buchan, Erica Mazaika, Elizabeth Ormondroyd, William T. Wright, Daniela Macaya, Chee Jian Pua, Birgit Funke, Daniel G. MacArthur, Sanjay Prasad, Stuart A. Cook, Mona Allouba, Yasmine Aguib, Magdi H. Yacoub, Declan P. O’Regan, Paul J. R. Barton, Hugh Watkins, Leonardo Bottolo, James S. Ware

## Abstract

Understanding the penetrance of pathogenic variants identified as secondary findings (SFs) is of paramount importance with the growing availability of genetic testing. We estimated penetrance through large-scale analyses of patients referred for diagnostic sequencing for hypertrophic cardiomyopathy (HCM; 10,400 cases, 1,340 variants) and dilated cardiomyopathy (DCM; 2,564 cases, 665 variants), using a cross-sectional approach comparing allele frequencies against reference populations (293,226 participants from UK Biobank and gnomAD). We generated updated prevalence estimates for HCM (1:543) and DCM (1:220).

In aggregate, the penetrance by late adulthood of rare, pathogenic variants (23% for HCM, 35% for DCM) and likely pathogenic variants (7% for HCM, 10% for DCM) was substantial for dominant CM. Penetrance was significantly higher for variant subgroups annotated as loss of function or ultra-rare, and for males compared to females for variants in HCM-associated genes.

We estimated variant-specific penetrance for 316 recurrent variants most likely to be identified as SFs (51% HCM and 17% DCM cases). 49 variants were observed at least ten times (14% of cases) in HCM-associated genes. Median penetrance was 14.6% (±14.4% SD). We explore estimates of penetrance by age, sex, and ancestry, and simulate the impact of including future cohorts.

This dataset is the first to report penetrance of individual variants at scale and will inform the management of individuals undergoing genetic screening for SFs. While most variants had low penetrance and the costs and harms of screening are unclear, some carriers of highly penetrant variants may benefit from SFs.

**Graphical Abstract:** A flowchart of the estimated penetrance for dominant cardiomyopathy by late adulthood for a variant of interest. The estimates of penetrance in this study are for carriers identified from unselected populations (e.g., consumer-initiated elective genomic testing or as secondary (2°) findings in clinical settings). If the variant is ultra-rare (i.e., identified once or less in population datasets), only estimates by variant subgroup in aggregate are available. If the variant is identified multiple times in both case and population datasets, variant-specific penetrance estimates may be available. If the variant is curated as a variant of uncertain significance (VUS), the penetrance estimate is low. If the variant is a likely pathogenic predicted loss of function (pLoF) variant and is identifiable multiple times in cases and population cohorts, penetrance estimates vary by gene (**Figure 2**). High aggregate penetrance represents an estimate of >25%; moderate aggregate penetrance represents 10%-25%, and low penetrance represents <10%. This flowchart was created with draw.io.

## Introduction

Cardiomyopathies (CM) are diseases of the heart muscle, characterised by abnormal cardiac structure and function that is not due to coronary disease, hypertension, valve disease, or congenital heart disease. Many cases have a monogenic etiology with autosomal dominant inheritance. Penetrance is incomplete and age-related, and expressivity is highly variable. These features present huge challenges for disease management. In particular, the penetrance of variants in CM-associated genes is incompletely characterised and poorly understood, especially when identified in an asymptomatic individual without family history of CM. With the growing availability of whole exome sequencing in wider clinical settings and consumer-initiated elective genomic testing^1^, the importance of estimating the penetrance of individual variants identified as secondary findings (SFs) to guide intervention is ever-increasing.

SFs are genetic variants that are actively sought out (as opposed to incidental findings), but which are unrelated to the clinical indication for genetic testing and can therefore be considered as opportunistic genetic screening. Genes associated with inherited CMs make up one-fifth of the 78 genes recommended by the American College of Medical Genetics and Genomics (ACMG SF v3.1) for reporting SFs during clinical sequencing^2^. It is recommended to return variants that would be classified as pathogenic or likely pathogenic in an affected individual and where there is >90% confidence that the variant is causing the observed disease^3^. This is independent of the probability that an individual carrying the variant will develop disease (penetrance). The ACMG SF guidelines have not yet been adopted globally, with the European Society of Human Genetics recommending a cautious approach with limited capacity, but responsive to accumulating evidence^4,5^.

We are concerned that the costs, harms, and benefits, have not been fully characterised. We have previously discussed issues with the recommendations based on the lack of estimates of the harms and cost of this approach for variants in specific genes^6^. These estimates are required to conform to the ninth rule of Wilson and Jugner’s principles of screening^7^. The burden of the implementation of reporting SFs in specific healthcare systems remains unassessed. There is little evidence for clinical utility and limited justification for use of resources^5^. Research is beginning to become available on patient perspectives and impact^8–12^.

Subclinical phenotypic expressivity of rare variants in CM-associated genes has been demonstrated in the UK Biobank population cohort^13–15^. Causes of variability in penetrance may include: i) genetic and allelic heterogeneity, with different alleles having different consequences on protein function, ii) environmental modifiers altering genetic influence (e.g., age, sex, hypertension, lifestyle), and iii) additional genetic modifiers with additive or epistatic interactions with the variant of interest (other variants or combinations of genetic factors, e.g. polygenic risk, variants in *cis* that drive allelic imbalance, imprinting, epigenetic regulation, compensation, threshold model, and transcript isoform expression)^16–22^.

Variant-specific estimates of penetrance are required to appropriately inform clinical practice and to fully utilise genetics as a tool to individualise the risk of developing disease in asymptomatic carriers^6,23^. It is challenging to estimate the penetrance of individual rare variants through other study methods as longitudinal population studies require very large sample sizes and long-term follow-up is required if penetrance is age-related. Where data is available for rare variants in CM-associated genes, reported penetrance is mostly estimated from family-based studies. These may be affected by ascertainment biases and secondary genetic and environmental factors^24^ and thus less applicable to SFs. Penetrance has been estimated in aggregate by gene and by disease^13,25,26^. Variant-specific penetrance in the general adult population for rare variants in CM-associated genes is unknown.

Here, we apply a cross-sectional approach, using a method that compares the allele frequency of individual rare variants in large cohorts of cases and reference populations to estimate penetrance. As well as providing aggregated penetrance estimates for groups of rare variants (e.g., those curated as pathogenic), this approach can estimate the penetrance of *individual* rare alleles. Importantly, these estimates represent variants in the general population, rather than in families ascertained for disease.

## Methods

### Case cohort

Sequencing data for 10,400 individuals referred for HCM gene panel sequencing, and 2,564 individuals referred for DCM gene panel sequencing was collected from seven international testing centres: three UK-based - the NIHR Royal Brompton Biobank, Oxford Molecular Genetics Laboratory, and Belfast Regional Genetics Laboratory; two US-based – the Partners Laboratory of Molecular Medicine and GeneDx; and the National Heart Centre, Singapore, and Aswan Heart Centre, Egypt. Although the diagnosis cannot directly be reconfirmed, given genetic testing guidelines (e.g.,^3,27^), a clinical diagnosis of CM is implicit. For information on DNA sequencing and data obtained for analyses, see the supplementary materials.

For each variant observed in one or more individuals referred for CM sequencing, we calculated the allele count (AC) and allele number (AN), and further stratified by reported age, sex, and ancestry, where the data allowed. All patients gave written informed consent, and the study was approved by the relevant regional research ethics committees.

### Population cohort

167,478 participants of the UK Biobank (UKBB) with whole exome sequencing data available for analyses and 125,748 exome sequenced participants of the Genome Aggregation Database (gnomAD; version v2.1.1) were included in this study.

Briefly, the UK Biobank recruited participants aged 40–69 years old from across the UK between 2006 and 2010^28^, of which the 200,571 exome tranche of individuals that had not withdrawn, were included in this study^29^. The maximal subset of unrelated participants was used, identified by those included in the UKBB PCA analysis (S3.3.2^28^, n=167,478). Age at recruitment, genetic sex, and genetic (for European (EUR) and British ancestry) or reported ancestry information (for other global ancestries: AFR, African, Caribbean (n=2,903); SAS, Indian, Pakistani, Bangladeshi (n=3,136); EAS, Chinese (n=605)) were incorporated.

The gnomAD database contains sequencing information for unrelated individuals sequenced as part of various disease-specific and population genetic studies^30^. The version 2 short variant dataset spans 125,748 exomes from unrelated individuals sequenced as part of various disease-specific and population genetic studies. Ensembl Variant Effect Predictor^31^ (VEP, version 105) was used to incorporate the variant-specific summary counts. Variants flagged by gnomAD as AC0 were excluded from gnomAD counts. For more information on the incorporation of these datasets, please see the supplementary materials.

### Variant annotation

VEP (105) was used to annotate the case and population datasets, with additional plugins: gnomAD^30^ (version r2.1), LOFTEE^30^, SpliceAI^32^ (1.3.1), REVEL^33^ (1.3), and ClinVar^34^ (20220115). The data was organised using PLINK^35^ (1.9) and the VEP output was analysed using R (4.1.2).

Protein altering variants, defined with respect to MANE transcripts, that were annotated as high or moderate impact by SO and ENSEMBL were included in the analysis. Analysis was restricted to genes with strong or definitive evidence of causing CM following ClinGen guidance^36,37^ and expert curation (gene2phenotype Cardiac Panel; https://www.ebi.ac.uk/gene2phenotype/), to include 8 sarcomeric HCM-associated genes (*MYH7, MYBPC3, MYL2, MYL3, ACTC1, TNNI3, TNNT2, TPM1*) and 11 DCM-associated genes (*BAG3, DES, DSP, LMNA, MYH7, PLN, RBM20, SCN5A, TNNC1, TNNT2, TTN*_*PSI>90%*_), with the exception of *FLNC* that was not included on the panel sequencing of the DCM case cohort (**Table S4**). Variants with consequences consistent with the known disease-causing mechanism were retained.

Further manual annotation was undertaken following ACMG guidelines using ClinVar^34^ and Cardioclassifier^38^, as previously published^13^. For analyses of variants in aggregate, the UKBB data was filtered following the same thresholds and used to estimate aggregate penetrance.

### Statistical analysis

Estimation of penetrance and 95% confidence interval Penetrance is expressed as a probability function on a scale of 0%-100%. Penetrance was estimated from case-population data in a Binomial framework following Bayes’ theorem for conditional probability^26^

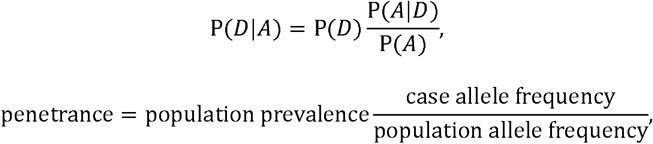

where, *D*, disease; *A*, allele; and P, probability, P(*D*|*A*) = penetrance (probability of disease given a risk allele), P(*D*) = fixed population baseline risk of disease (probability of disease), P(*A*|*D*) = allele frequency in the case cohort (probability of the allele given disease), P(*A*) = allele frequency in the population cohort (probability of the allele).

We define penetrance in this setting as the probability of dominant CM by late adulthood (UKBB had a mean age of 56 years old at recruitment). We assume the independence of the random variables in the penetrance equation above to derive the 95% confidence interval for penetrance as the product and ratio of binomial proportions. We used the specialised version of the Central Limit Theorem, the Delta method, on the log-transformed random variable log(*D*|*A*) *=* log(*D*) + log(*A*|*D*) − log (*A*) with an improved mean approximation and adjustment for degeneracy (as allele frequency tends to 0 for rare variants). Please see additional methods and alternative approaches considered (**Supplementary methods, Table S3, Lists S1-S2, Figures S4-S5**).

For estimates of penetrance by sex, we adjusted all terms of the penetrance equation by values for sex-specific parameters. For estimates of penetrance by ancestry, we kept P(*D*) as estimated for CM (there are few estimates of the prevalence of CM in specific ancestries) and proportioned P(*A*|*D*) and P(*A*) by reported ancestry. For estimates of penetrance by age, we normalised P(*D*) by the number diagnosed in the case cohort by a particular age in a cumulative fashion, with P(*A*|*D*) by a particular age, and P(*A*) fixed as total population allele frequency (**Supplementary methods**).

### Estimated cardiomyopathy prevalence

To incorporate P(*D*) in our penetrance analysis, we estimated the uncertainty surrounding the reported prevalence of CM (**Tables S1-S2, Figures S1-S3**). For HCM, we meta-analysed four imaging-based prevalence estimates^13,39–41^ excluding studies with potential selection biases. From the meta-analysis estimate (*p*_*D*_ ≡ P(*D*)) and its confidence interval, we derived values of allele count, *x*_*D*_, and allele number, *n*_*D*_ (where 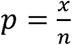). A literature review was also completed for DCM, but there were not enough imaging-based prevalence estimates in literature, so 39,003 participants of the UK Biobank imaging cohort were used to estimate phenotypic DCM^42–44^ (**Supplementary methods**). Using the same methods and included studies, we derived estimates for male- and female-specific HCM and DCM prevalence.

## Results

### Case cohort summary information

Sequencing data for 10,400 individuals referred for HCM genetic panel sequencing and 2,564 individuals referred for DCM genetic panel sequencing were included in the analysis. Aggregated frequency of rare protein altering variants in well-established disease genes was 42% for HCM and 33% for DCM in the respective case cohorts (**Tables S6-S7**). Of the cohorts with age, sex, and ancestry information available (20% of HCM cases, 42% of DCM cases), 35% and 32% were female, 93% and 91% were of EUR ancestry, and mean age was 48 and 49 years old, for HCM and DCM, respectively (**Table S5**).

### New estimates of the prevalence of cardiomyopathies

To estimate the prevalence of CMs, a literature review and meta-analysis were undertaken (**Tables S1-S2, Figures S1-S3**). Prevalence is underestimated when derived from national cohorts using coding systems such as ICD codes, due to incomplete ascertainment through diagnostic and procedure coding^45^. We would therefore expect the most accurate estimates of the prevalence of CM to come from imaging studies in populations, where echocardiogram or cardiac magnetic resonance imaging was used to identify CM within a population sample that is representative. The estimates are not generalisable if the prevalence is estimated for selected subgroups of individuals, such as young, elderly, or athletic cohorts. We therefore meta-analysed four imaging-based prevalence estimates which resulted in a HCM population prevalence estimate of 1 in 543 individuals (*p*_*D*_ = 0.18% (95% *CI*_*D*_ = 0.15%-0.23%))^13,39–41^. The well reported estimate of 1 in 500 individuals for HCM prevalence (0.20%) is within this confidence interval.

A literature review revealed insufficient imaging-based estimates to undertake a direct meta-analysis of the prevalence of DCM. Instead, we used 39,003 participants of the UK Biobank imaging cohort to estimate phenotypic DCM^42–44^. This derived a DCM population prevalence of 1 in 220 individuals (*p*_*D*_ *=* 0.45% (95% *CI*_*D*_ *=* 0.39%-0.53%)), which includes the well reported estimate of 1 in 250 (0.40%)^46^ within the confidence interval.

We also estimated sex-specific CM prevalence. This resulted in a HCM population prevalence of ∼1 in 1,300 females (*p*_*D*_ *=* 0.08% (95% *CI*_*D*_ *=* 0.04%-0.12%)) and ∼1 in 360 males (*p*_*D*_ *=* 0.28% (95% *CI*_*D*_ *=* 0.22%-0.35%)), and a DCM population prevalence of ∼1 in 340 females (*p*_*D*_ *=* 0.30% (95% *CI*_*D*_ *=* 0.23%-0.38%)) and ∼1 in 160 males (*p*_*D*_ *=* 0.63% (95% *CI*_*D*_ *=* 0.52%-0.75%)).

### Estimated penetrance of rare variants in aggregate

In individuals with cardiomyopathy referred for diagnostic sequencing, we identified 1,340 rare (inclusive population allele frequency of <0.1%) variants in HCM-associated genes (4,314 observations, case frequency 42%) and 665 rare variants in DCM-associated genes (833 observations, case frequency 33%) (**Tables S6-S9**). The UKBB dataset was filtered following the same pipeline. 1,735 rare variants in HCM-associated genes (9,196 observations, 5.5% population frequency) and 4,586 rare variants in DCM-associated genes (22,232 observations; 13.3% population frequency) were used to estimate aggregate penetrance of rare variant subgroups. Variants with a pathogenic classification in ClinVar were the most penetrant subgroup by ACMG classification^45^ (HCM 22.5% (17.5%-28.8%), DCM 35.0% (21.6%-56.8%); **Figure 1, Table S15**).

**Figure 1.**
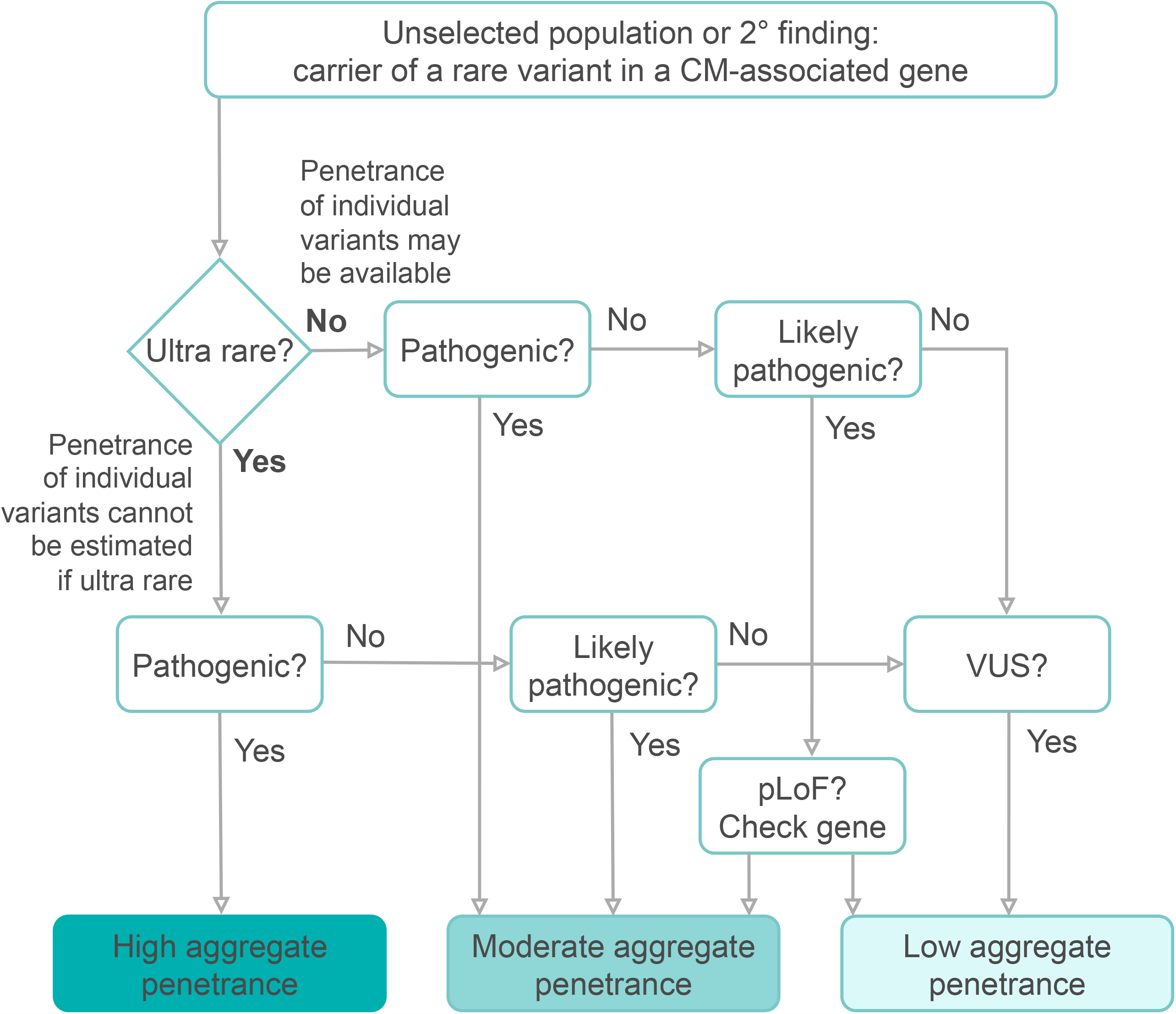
Aggregated penetrance of rare variants by variant curation, rarity, age, and sex. In aggregate, variants curated as pathogenic and variants that are particularly rare (gnomAD AC=0) were most penetrant. The plot depicts aggregated estimated penetrance for rare variants in HCM-(A, B, C, D) and DCM-associated (E, F, G, H) genes. Variant curation was assessed following ACMG guidelines through ClinVar and CardioClassifier software with additional manual curation of variants with conflicting evidence (A, E (for HCM cases: 173 P variants, 316 LP, 832 VUS, 19 LB; for the UK Biobank: 30 P, 98 LP, 1552 VUS, 53 LB, 2 B; for DCM cases: 21 P, 245 LP, 358 VUS, 37 LB, 4 B; for the UK Biobank: 15 P, 505 LP, 3951 VUS, 108 LB, 7 B)). The variants were assessed for rarity by gnomAD allele count (AC) bins, where 0 is not identified in the gnomAD dataset (B, F). Age was assessed in decades based on the cumulative proportion of cases analysed by each age timepoint (C, G). Sex was estimated using all parameters stratified by reported sex (D, H).

**Figure 2.**
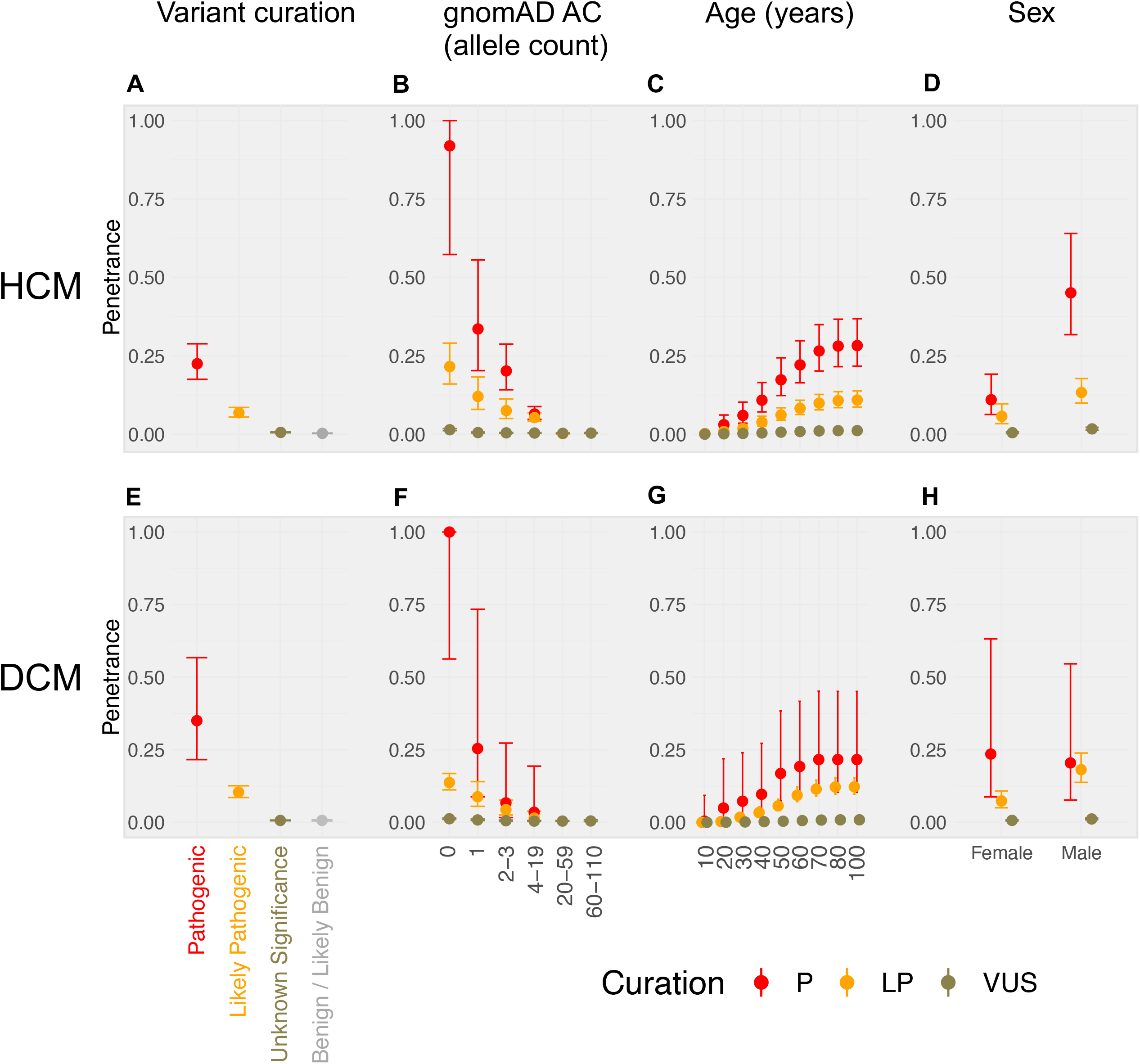
The aggregate estimates of penetrance of “loss of function” variants are high for specific genes. The plot depicts estimated penetrance of HCM-associated (left) and DCM-associated (right) rare variants. pLoF and non-pLoF variant groups are plotted in green and blue, respectively. pLoF, predicted loss of function variants; *, TTNtv that are PSI>90%. Pathogenic *TNNT2* inframe deletions caused an increased penetrance signal for inframe deletions for both HCM and DCM (see **Figure S12**).

An estimate of the aggregate penetrance of both pathogenic and likely pathogenic variants in HCM was 10.7% (8.6%-13.3%) using this approach, concordant to a recent estimate derived using direct assessment of cardiac imaging in UKBB (10.8%; variant carriers with LVH ≥13mm without hypertension or valve disease; binomial 95% confidence interval of 3.0%-25.4%; n=4/37)^11^. This concordance was also observed for other variants in the same paper (e.g., VUS) which we estimated penetrance as 0.55% (0.45%-0.68%) compared to 0.57% (0.07%-2.03%, n=2/353)^11^.

The aggregate penetrance of pathogenic and likely pathogenic variants in DCM was 11.3% (9.3%-13.6%). Population penetrance of rare variants in DCM-associated genes in UKBB has previously been estimated at ≤30%^47^ for a clinical or subclinical diagnosis in an analysis of 44 DCM-associated genes, and in the range of 5%-6% for TTNtvs (1.9%-12.8%; 877 carriers)^6^ depending on the definition used. We report a concordant penetrance estimate from our analysis of strong- and definitive-evidence DCM genes only, and 9.8% (8.0%-12.1%) for TTNtvs (**Figure 2, Figure S12**).

Variants predicted to result in premature termination codons (PTCs; nonsense mediated decay competent or incompetent^48^) in *MYBPC3, BAG3, DSP*, and *LMNA*, were the most penetrant (**Figure 2, Figure S12, Tables S13-S14, Tables S18-S19**). Inframe deletions in *TNNT2* were highly penetrant for both HCM and DCM. TTNtvs and missense variants predicted to be damaging in *TPM1* and *TNNC1*, had moderate penetrance.

Stratification by variant rarity showed that variants absent from the gnomAD database were the most penetrant subgroup (HCM pathogenic 91.9% (57.3%-100.0%), HCM likely pathogenic 20.2% (14.2%-28.7%2), DCM pathogenic 100.0% (56.3%-100.0%), DCM likely pathogenic 13.7% (11.2%-16.8%); **Figure 1, Table S16**). Stratification of penetrance by sex identified increased penetrance for males compared to females for rare variants in HCM-associated genes (**Figure 1, Figure S13, Table S20**). Penetrance was estimated as <20% up to 50 years of age by modelling the penetrance of CM as an age-related cumulative frequency using the proportion of cases referred at each age decile (**Figure 1, Table S17**).

While there are limitations to the cohort size when split by reported ancestry and we are unable to rule out local ancestry mismatches between case and population datasets, there was no significant difference in the penetrance of truncating variants in *TTN* (TTNtvs_PSI>90%_) between African (5.7% (2.9%-10.9%)), European (6.9% (5.5-8.5%)), East Asian (6.1% (3.0%-12.4%)), and South Asian (5.7% (2.1%-15.8%)) ancestries, as previously suggested^49^.

### Estimated penetrance of individual rare variants

Of the variants identified and used to estimate penetrance in aggregate, we report four subgroups of variants in our case series (**Figure 3**):

**Figure 3.**
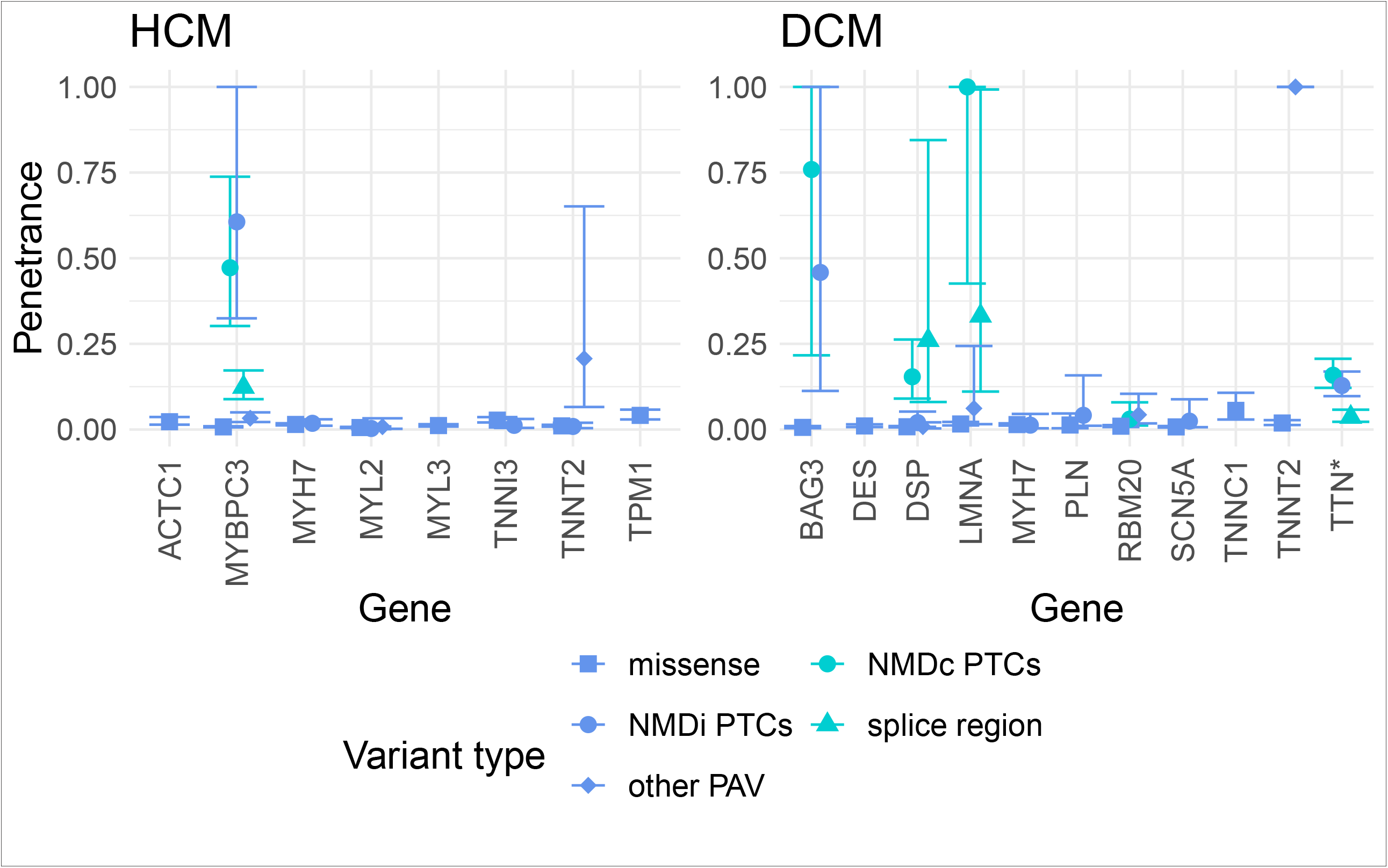
Penetrance of individual variants could be estimated for 316 recurrently observed rare variants from group 2. The figure shows variant counts and subgroups for rare variants in HCM-associated (left) and DCM-associated (right) genes. The colour of the description matches the subgroup of variants depicted. The pie charts plot the proportion of all variant observations in each subgroup (also denoted as ‘G+’). The observations approximate to the number of variant carriers, though a small number of individuals may carry more than one variant. G+, denotes the proportion of all variants identified; all, frequency of the variant in cases; obs, observations of allele count.

Group 1 consisted of 339 variants that were found in more than one case (case allele count (AC) ≥2) and were ultra-rare in population reference sets (population AC (popAC) ≤1). Penetrance cannot be estimated with precision for individual variants in this group, since the population allele frequency (AF) cannot be estimated with precision. When considered in aggregate this group have high penetrance (**Figures S14**). For HCM, 294 variants in group 1 were identified 1,322 times (13% case frequency, 31% observations). 29% were curated as pathogenic (P, n=84, 41% of HCM group 1 observations), 34% were likely pathogenic (LP, n=100, 36% observations), and 37% were curated as uncertain significance (VUS, n=110, 23% observations). For DCM, 45 variants in group 1 were identified 132 times (5% case frequency, 16% observations). 18% of these were P (n=8, 20% DCM group 1 observations), 49% LP (n=22, 55% observations), and 33% VUS (n=15, 25% observations).

Group 2 included 316 variants found multiple times in both cases and population reference datasets (caseAC≥2, popAC≥2). This group is expected to include variants with intermediate penetrance, including founder effect variants. For this group, we can estimate AF in both populations, and therefore can estimate penetrance (**Figure 4, Interactive Figure 4, Tables S10-S11**). These account for more than half of all variants identified in HCM-associated genes and include those most likely to be identified as SFs. For HCM, 257 variants were identified a total of 2,203 times (21% case frequency, 51% observations). 11% were P (n=29, 37% HCM group 2 observations), 25% LP (n=64, 31% observations), 59% VUS (n=151, 29% observations), and 5% likely benign (LB, n=13, 3% observations). 49 of these variants were recurrent at least ten times and described a large portion of observations (case AC≥10; found 1,424 times, 33.0% of case cohort observations, case frequency of 13.7%). The median penetrance of these was 14.6% (±14.4% SD). For DCM, 59 variants were identified 140 times (6% case frequency, 17% observations). None were curated as P, 24% were LP (n=14, 22% DCM group 2 observations), 56% VUS (n=33, 53% observations), 17% LB (n=10, 21% observations), and 3% B (n=2, 4% observations). With the current DCM case cohort size, no variant was identified ten or more times.

**Figure 4.**
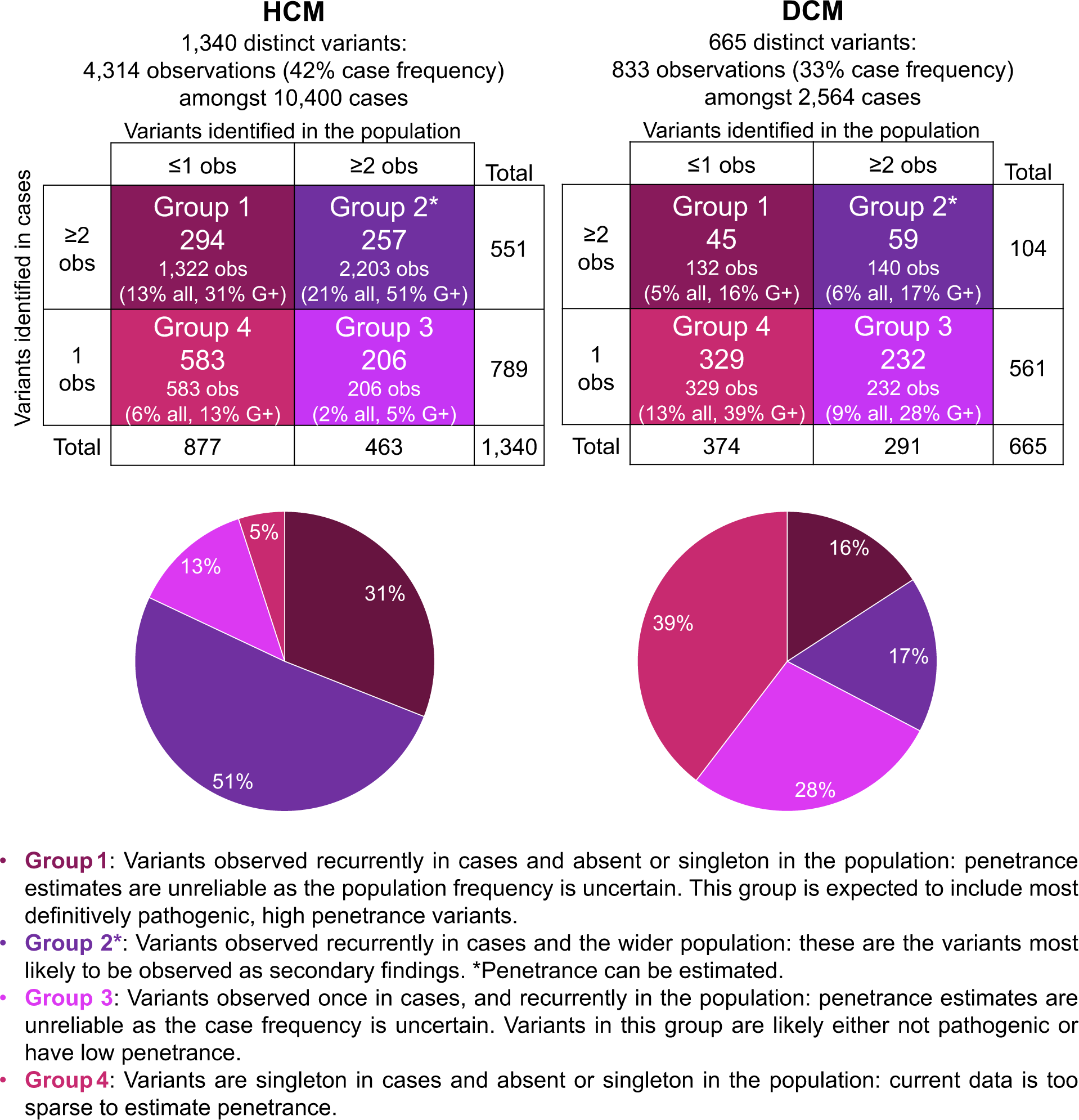
Variant-specific estimates of penetrance for the 316 recurrently observed rare variants in CM-associated genes from group 2. An interactive widget is available for browsing the individual variants in this figure (**see Figure4_interactive.html**). The variants depicted (HCM n=257 (top), DCM n=59 (bottom)) were identified multiple times in cases and population reference datasets and penetrance could therefore be estimated. The x-axis denotes the number of times the variant was observed in each case cohort. AC, allele count; B/LB, benign/likely benign; VUS, variant of uncertain significance; LP, likely pathogenic; P, pathogenic.

The final two groups consisted of 1,350 variants with only a single observation in our case series. This does not provide a reliable of case frequency so penetrance estimates would lack precision. Group 3 variants were those identified multiple times in the population (popAC≥2), and consisted mostly of VUSs: For HCM, 206 variants were identified (2% case frequency, 5% of case observations). This included 0.5% P (n=1; MYBPC3:c.3297dup:p.(Tyr1100Valfs*49)), 5% LP (n=10), 92% VUS (n=189), and 3% LB (n=6). For DCM, 232 variants were identified (9% case frequency, 28% observations). 1% were P (n=3), 7% LP (n=17), 79% VUS (n=183), 12% LB (n=27), and 1% B (n=2).

Group 4 variants are those observed once in cases, and rarely in the population reference dataset (popAC≤1). A substantial portion of these were P/LP: for HCM, 583 variants were identified (6% case frequency, 13% observations). 10% were P (n=59), 24% LP (n=142), and 66% VUS (n=382). For DCM, 329 variants were identified (13% case frequency, 40% observations). 3% were P (n=10), 58% LP (n=192), and 39% VUS (n=127).

### The impact of age, sex, and ancestry, on variant-specific penetrance estimates

For group 2, where age-related penetrance could be derived, we estimated the penetrance of specific variants by decade of age (e.g., **Figure 5**). For some variants (e.g., MYBPC3:c.1624G>C:p.(Glu542Gln)), the age-related penetrance curve shows infrequent onset before middle age. These curves may inform surveillance strategies in variant carriers unaffected at first assessment.

**Figure 5.**
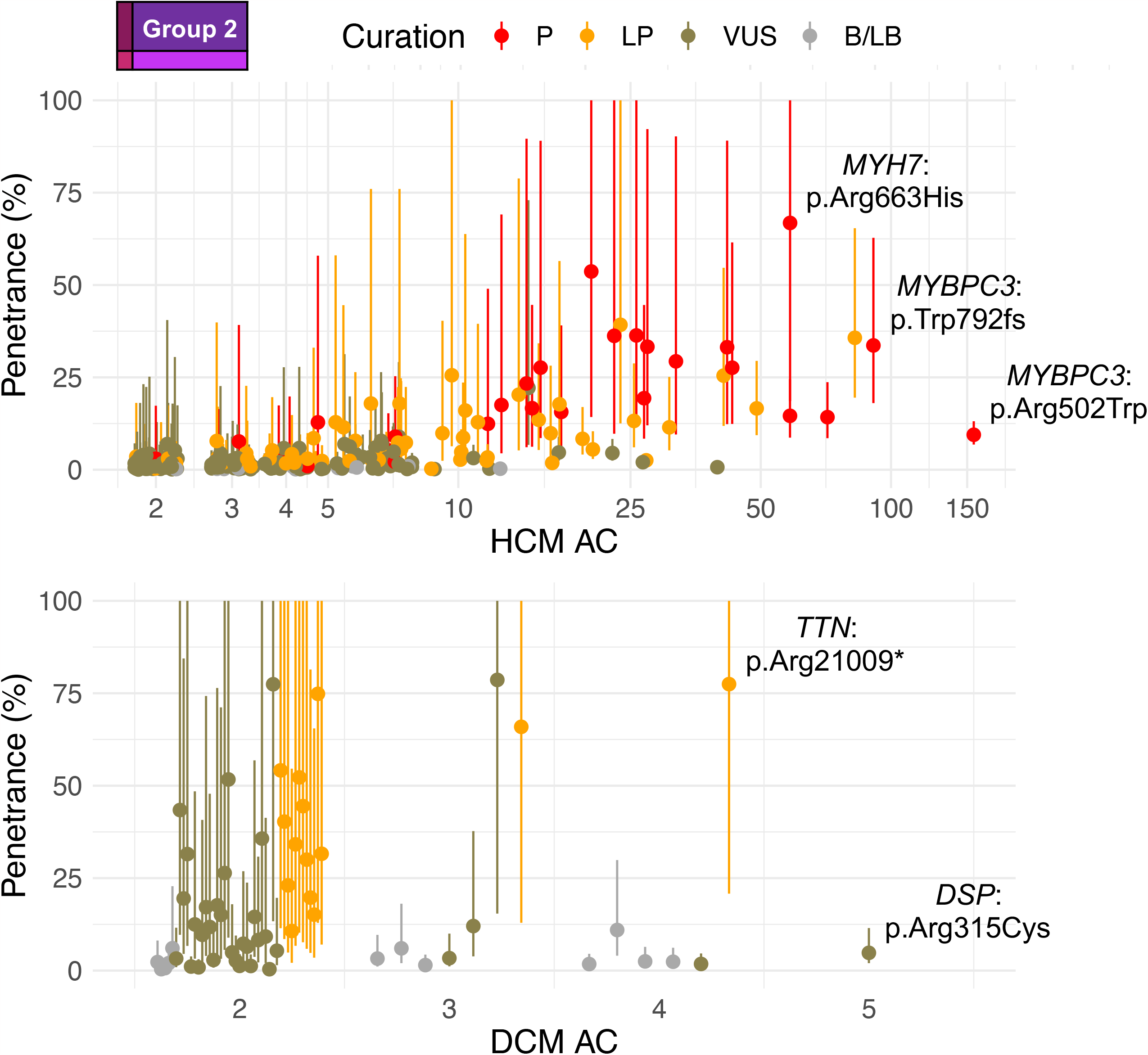
Variant-specific estimates of penetrance by age can now be derived. The plot depicts the age-related cumulative penetrance of five HCM-associated rare variants across age deciles from variant group 2. The x-axis starts in the decade of the 20s as the analysis of these variants was underpowered for teens and younger. “20s” here means “by 30 years old”.

We identified rare variants in HCM-associated genes where estimated penetrance for males was significantly increased compared to females (**Figure S13**). Identification of such variants allows for future investigations of modifiers protecting female carriers from disease.

For estimates of penetrance by ancestry, variants that were nominally more common in AFR, EAS, or SAS ancestries compared to EUR ancestry were identified (**Table S12**). We interpret these as more consistent with inaccurate penetrance estimation arising from ancestries where the variant is sparsely observed, rather than true differences in penetrance on different ancestral background. For example, MYBPC3:c.1544A>G:p.(Asn515Ser) was identified 5/492 times in AFR cases (AF=0.005) and 33/10,655 times in the AFR population participants (AF=0.0016; penetrance of 0.6% (0.2%-1.5%)) compared to 1/9,692 times in EUR cases (AF=0.00005) and not observed in 211,532 EUR population participants. Even when ancestry is nominally matched, broad continental groupings hide great diversity and results may be misleading due to stratification between cases datasets (mostly North AFR from Egypt) and population reference datasets (e.g., UK Biobank participants from the Caribbean) (**Box 1**).

### Clinical impact of specific variants now shown to have low penetrance

We can define the upper bound of the penetrance estimate for some variants. 162 rare variants in HCM-associated genes (63% of variants, observed 745 times (7% case frequency; 17% of observations) have a penetrance of ≤10%, according to the upper limit (UCI) of the 95% CI for our estimate. These included two variants previously curated as definitively pathogenic, and 25 variants curated as likely pathogenic.

One of the pathogenic variants is splice acceptor MYBPC3:c.26-2A>G which has an estimated penetrance of 1.0% (0.4%-2.8%) or 0.9% (0.3%-2.5%) in EUR ancestry, as it was identified 4 times in EUR cases and 20 times in population participants (90% were EUR). The potential for this variant to have incomplete penetrance has been noted previously through identified asymptomatic carriers (see ClinVar ID 42644). There is in silico evidence of an alternate splice site downstream that could result in an in-frame deletion of two amino acids.

The second pathogenic variant identified with a UCI of ≤10% is the missense variant MYH7:c.3158G>A:p.R1053Q which is a Finnish founder mutation. This variant had an estimate penetrance of 2.2% (0.9%-5.2%), as it was identified 7 times in EUR cases and 17 times in the population cohort (16 Finnish from gnomAD, 1 NWE from UK Biobank). Estimates of penetrance are sensitive to allele frequency differences across ancestries. Analysis of founder mutations in the population they derive from would provide additional confidence in their penetrance estimates.

For DCM, 17 rare variants (29% of variants) observed 45 times (2% case frequency; 5% of observations) met this criterion. None of the 17 variants were curated as P/LP.

#### Box 1

Case study: The MYBPC3:c.1504C>T:p.(Arg502Trp) Northwestern European variant

The variant MYBPC3:c.1504C>T:p.R502W was found in our cohort 159 times in patients referred for HCM genetic panel sequencing (3.7% of total observations; 1.5% total case frequency). To date, the variant has been classified on ClinVar fifteen times as pathogenic (ClinVar ID 42540). Penetrance has been previously estimated as ∼50% (increased relative risk of 340) by 45 years old in a clinical setting, with major adverse clinical events in carriers significantly more likely when another sarcomeric variant is present^50^.

In our case cohort, carriers of this variant were reported as broadly European ancestry (Oxford, n=59; London, n=11; Belfast, n=30; LMM, n=45; GDX, n=14). In gnomAD, the variant was identified 10 times, of which 7 carriers were non-Finnish Northwestern Europeans (NWE; plus 1 African; 1 South Asian, and 1 other), and in the UK Biobank, the variant was found 77 times, of which 68 carriers were NWE (plus 8 other Europeans, and 1 other). The population frequency of the variant in Ensembl population genetics showed that the variant (rs375882485) is only found multiple times in NWE ancestry sub cohorts. Thus, the variant is most common in NWE populations: the UK, Ireland, Belgium, the Netherlands, Luxembourg, Northern France, Germany, Denmark, Norway, Sweden, and Iceland.

We use this relatively common variant to highlight the effect of ancestry on estimated variant penetrance (**Figure 6**): the penetrance was estimated as 6.4% (4.6%-9.0%) using the UK Biobank cohort (93% European) and this is inflated to 35.1% (18.2%-67.5%) when estimating the penetrance using the gnomAD dataset (45% European), due to the difference in the proportion of individuals with NWE ancestry. In individuals of NWE ancestry only, the penetrance of this variant is 6.4% (4.6%-9.0%). Penetrance estimated from the NWE subset of gnomAD or UKB do not differ significantly.

**Figure 6.**
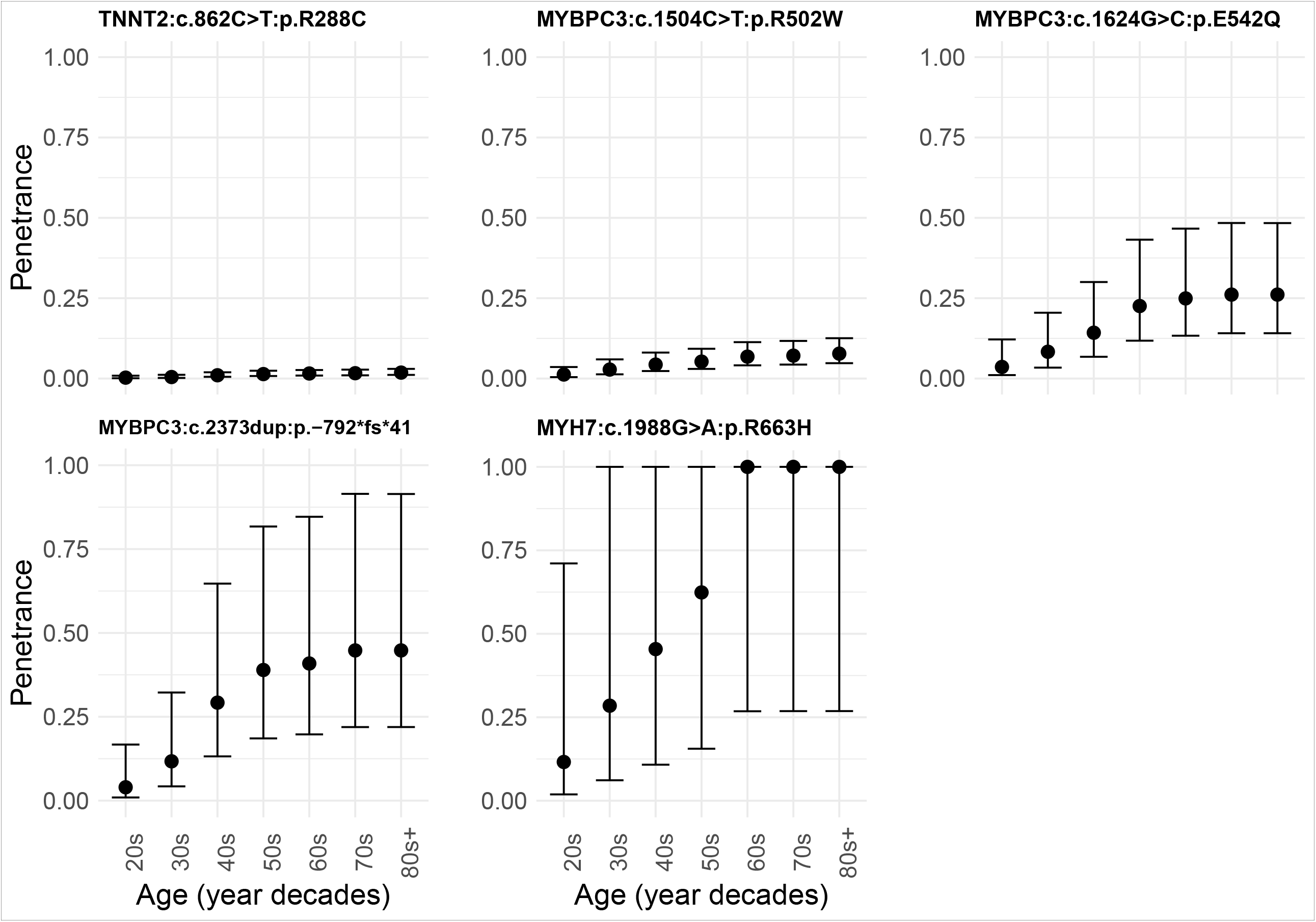
Penetrance estimates are inflated with underestimated population frequency. The map of the world emphasises the large proportion of observations of *MYBPC3*:c.1504C>T:p.(Arg502Trp) in HCM cases of Northwestern European (NWE) ancestry. The numbers on the map are the counts of rare variant genotype positive observations (n≈cohort participants) from each cohort with the specified ancestry, and the percentages derive the proportion of observations that are due to the *MYBPC3*:c.1504C>T:p.(Arg502Trp) variant. The graph shows the penetrance estimated for the variant based on subgroups of reference dataset participants included. The penetrance is inflated when estimated using gnomAD due to the variant being most common in participants with NWE ancestry (which dominates the UKBB dataset). Population frequency of gnomAD, UK Biobank, and Ensembl population genetics showed that this variant (rs375882485) is only found multiple times in NWE ancestry sub cohorts. The map excludes Antarctica for figure clarity. A limitation is the low sample sizes for AFR, SAS, and EAS, ancestries.

As access to larger genomic datasets becomes available, including more diverse ancestries, we can increase the precision of these variant-specific penetrance estimates by gaining further confidence in maximum population allele frequencies^51^.

### Penetrance estimate simulations of increased cohort sizes

We anticipate two benefits to estimating the penetrance of rare variants from increasing cohort sizes: i) there will be more variants that are observed recurrently in cases and populations, permitting AF estimates and hence penetrance estimates, and ii) the precision of our penetrance estimates will increase as AF of rare variants is ascertained with greater precision.

We sought to understand whether it would be more valuable to focus resources on aggregating data from larger numbers of cases (plausible ∼100,000 cases with global collaboration efforts), and/or from larger numbers of population participants with near-term publicly available population datasets (∼5,000,000 participants).

Efforts to increase reference population sample size will provide additional confidence in penetrance estimates once case aggregation to 10,000 cases is reached (**Figure S6**). There is substantial confidence to be gained by increasing the population cohort size: we found that increasing the population dataset from 300,000 participants to 4.5 million participants could provide ∼20% certainty, depending on the penetrance of the variant (**Figures S7-S11**). The increase in confidence gained from increasing the case cohort sample size from 10,000 cases to 100,000 cases was limited (with the caveat that more variants will be identified).

## Discussion

We show that some subgroups of rare variants in the population are penetrant and for these it may be reasonable to return as SFs. These include ultra-rare variants, predicted PTCs in certain genes where loss of function is a known disease mechanism, and variants with enough evidence to have been classified as definitively pathogenic previously (**Graphical abstract**).

There is still uncertainty regarding the penetrance of individual ultra-rare variants, and the implications of returning SFs in healthcare systems have yet to be estimated. While we have previously attempted to assess the burden of long-term surveillance for DCM^6^, cost-effect analyses are vital to fully understand the risks and benefits of reporting SFs in different healthcare systems. For variant types with low penetrance, it is very uncertain that the benefit of returning SFs will outweigh harms and justify costs.

Here, we provide the first at-scale estimates of variant-specific penetrance for variants in CM-associated genes that include those likely to be most frequently identified as SFs. Most have low estimated penetrance, where an asymptomatic individual without family history for disease may choose no or less frequent surveillance, depending on the healthcare system and follow-up cost.

Population penetrance estimates derived from unselected individuals (with certain caveats^52^) that are agnostic to personal or family history of disease, should provide a better estimate of the probability of manifesting disease when a variant is identified as a SF. Importantly, the penetrance of variants found in CM patients and relatives in a clinical setting is increased compared to the penetrance of variants estimated for those identified through SFs (e.g., MYBPC3:c.1504C>T:p.R502W with estimated penetrance of 50% in HCM patients and 6% here, in the population).

While published data is sparse and heterogeneous, overall estimates of penetrance by adulthood in the general population are lower than family-based studies. An abstract reported the penetrance of asymptomatic carriers referred to hospital for predictive testing after identification of a genotype- and CM-positive relative^53^. For HCM, 17 of 65 carriers (26.2%) were diagnosed with HCM (10 on first clinical evaluation, 7 during 2 years of follow up). For DCM, 2 of 22 carriers (9.1%) were diagnosed with DCM (2 on first clinical evaluation, 0 during 2 years of follow up (excluding 5 with hypokinetic non-dilated cardiomyopathy and 4 with isolated left ventricular dilatation))^53^. Additionally, a study of carriers identified during family screening who did not fulfill diagnostic criteria for HCM at first evaluation identified HCM or an abnormal ECG in 127 of 285 carriers (44.6%; 82 at baseline, 45 over a median of 8 years follow-up)^25^. First degree relatives in the same household may be at increased risk of disease due to shared environment and other genetic factors.

The ACMG guidelines for reporting “medically actionable” variants in 78 genes come with the caution that evaluating SFs requires an increased amount of supportive evidence of pathogenicity, given the low prior likelihood that variants unrelated to the indication are pathogenic^54^. Here, we show that variants with a definitive pathogenic assertion in ClinVar had the highest penetrance estimates. This may be because penetrant variants are more likely to yield sufficient evidence for confident interpretations, especially family segregation data.

Genetic laboratories communicate their confidence on whether a variant has a role in disease (i.e., pathogenicity), but do not consistently indicate the penetrance. Pathogenicity addresses whether a variant explains the aetiology of a patient. Penetrance addresses the probability of future disease in carriers. The ClinGen consortium Low-Penetrance/Risk Allele Working Group recommends providing penetrance estimates on clinical reports (aggregate gene-level or individual variants) and noting when penetrance is assumed or where current information is limited/unavailable.

TTNtv are common in the general population (∼1 in 250 for variants in exons constitutively expressed in the adult heart), and we show that the penetrance in aggregate of TTNtvs is reduced compared to predicted loss of function variants in other CM-associated, haploinsufficient genes. While recent work has increased our understanding of the functional mechanisms of TTNtvs in disease^55,56^, future work is required to identify modifiers of TTNtvs to understand this reduced penetrance in the population.

The penetrance of a variant may depend on characteristics of the variant itself and modulating effects of genetic background and environment. This study characterises individual variants, while ongoing work is dissecting the role of secondary genetic influences. Polygenic scores may identify individuals at particular risk of disease, modifying the estimated penetrance of a single dominant variant.

We present two novel dimensions to estimates of penetrance: the penetrance in the general population and variant-specific penetrance. As described, the results of this method are concordant with previous population estimates of aggregated penetrance in the UKBB population derived using independent approaches, providing confidence in the methods. In addition, we provide updated estimates for the population prevalence of HCM and DCM and stratify by sex. The addition of future, publicly available, large-scale, global population datasets and biobanks will aid this area of research by allowing for increased confidence in ancestry-specific population allele frequencies and CM prevalence. We provide the summary counts for each variant via an online browser and the function to estimate penetrance in R for transferability and use in other diseases and datasets.

### Limitations

This study has not been undertaken without careful consideration of the limitations. This method cannot quantify the penetrance of pathogenic variants that are absent/singleton in the population, while in aggregate the penetrance of this group of variants is significant.

Comparisons of case and control allele frequency are vulnerable to confounding by population stratification, and we have explored some examples in this manuscript. We do not have genome-wide variation data to directly assess genetic ancestry for the case cohort, so this is based on data reported by the referring clinician. As the EUR participants dominate our case and population datasets, greater representation of diverse ancestral backgrounds is essential for equitable access to genomic medicine. Estimates of the penetrance of variants and the prevalence of cardiomyopathies in more ancestral groups are required. The current data for both comes from UK Biobank which has limitations^52^.

In the absence of genome-wide data we cannot exclude the possibility of unrecognised or cryptic relatedness within the case cohort. As described by Minikel et al. (2016)^26^, when a variant is highly penetrant, cryptically related individuals are likely included in case series and if a disease is fatal, population cohorts are likely depleted of causal variants.

Case allele frequency in unrelated cases may not be a fair estimate of the case allele frequency in all cases observed in the clinic. Our estimate of case allele frequency, and therefore of penetrance, is influenced by genetic testing referral practice. If clinicians are cautious and only refer selected high confidence cases for testing, case allele frequency and estimated penetrance will be high, whereas if clinicians were to test widely and indiscriminately then our apparent case allele frequency would be lower, resulting in lower penetrance estimates^18^.

Current diagnostic data assumes that the testing centre obtained complete coverage of the gene. Limited data was available on age and sex for large portions of the case cohorts. Our DCM referred cohort was only moderate in size and thus, increases in sample size here through global collaboration would aid our estimates of penetrance for variants in DCM-associated genes.

Finally, the UKBB volunteer population cohort is healthier than the average individual^52^, and the gnomAD consortium includes some individuals with severe disease but likely at a frequency equivalent to or lower than the general population^30^. The proposed penetrance model is an approximation since in reality the three parameters used on the right-hand side of the penetrance equation share some degree of dependence.

## Conclusion

This is the first evaluation of the penetrance of individual rare variants in CM-associated genes at scale. These recurrent variants are those that are likely to generate SFs. Variants previously annotated as pathogenic, loss-of-function variants in specific genes susceptible to haploinsufficiency, and those that are the rarest in the population, have high penetrance, similar to observations from family studies. This first attempt at estimating the penetrance of rare variants has highlighted the requirement for large case and population datasets with known genetic ancestry. We are now able to start putting bounds on the estimate of penetrance for a specific variant identified as a secondary finding: for some, including those expected to be most penetrant, we do not currently have enough data, for others, we can provide asymptomatic variant carriers with an estimated probability of manifesting disease.

## Supporting information

Interactive Figure 4

## Data Availability

All case cohort data arising from this analysis is available through DECIPHER. Both gnomAD and UK Biobank population reference datasets are publicly available. Analysis code is available on GitHub.

https://www.deciphergenomics.org/

https://gnomad.broadinstitute.org/

https://www.ukbiobank.ac.uk/

https://github.com/ImperialCardioGenetics/variantfx/tree/main/PenetrancePaper

## Data and code availability

All case cohort data arising from this analysis is available through DECIPHER (https://www.deciphergenomics.org/). Both gnomAD (https://gnomad.broadinstitute.org/) and UK Biobank (https://www.ukbiobank.ac.uk/) population reference datasets are publicly available. Analysis code is available on GitHub (https://github.com/ImperialCardioGenetics/variantfx/tree/main/PenetrancePaper).

## Acknowledgements and funding sources

This work was supported by the Wellcome Trust [107469/Z/15/Z; 200990/A/16/Z], Medical Research Council (UK) [MC_UP_1605/13], British Heart Foundation [RG/19/6/34387, RE/18/4/34215, FS/IPBSRF/22/27059], and the NIHR Imperial College Biomedical Research Centre. The views expressed in this work are those of the authors and not necessarily those of the funders. For open access, the authors have applied a CC BY public copyright license to any Author Accepted Manuscript version arising from this submission.

## CrediT statement

Conceptualization: J.S.W, K.A.M.; Methodology: J.S.W., K.A.M., L.B., X.Z., P.T.; Formal Analysis: K.A.M., L.B., P.T., K.T., A.H.; Resources: K.T., R.B., W.T.W., D.M., P.C.J., B.F., D.M., S.P., S.C., M.A., Y.A., M.H.Y., D.O’R., H.W., J.SW.; Data curation: K.A.M., P.T., E.M.; Writing – original draft: K.A.M.; Writing – review & editing: (all authors); Visualization: K.A.M., J.S.W.; Supervision: J.S.W., H.W.; Project administration: J.S.W., P.B.

## Ethics declaration

All research participants provided written informed consent, and the studies were reviewed and approved by the relevant research ethics committee (*Aswan Heart Centre*: FWA00019142, Research Ethics Committee code 20130405MYFAHC_CMR_20130330; *NIHR Royal Brompton Biobank*: South Central – Hampshire B Research Ethics Committee, 09/H0504/104+5, 19/SC/0257; *National Heart Centre Singapore*: Singhealth Centralised Institutional Review Board 2020/2353 and Singhealth Biobank Research Scientific Advisory Executive Committee SBRSA 2019/001v1; *UK Biobank*: National Research Ethics Service 11/NW/0382, 21/NW/0157, under terms of access approval number 47602).

In addition, diagnostic laboratories (Oxford Molecular Genetics Laboratory, Belfast Regional Genetics Laboratory, the Partners Laboratory of Molecular Medicine, and GeneDx) provided aggregated (and therefore fully anonymous) cohort-level summaries of variant data collected for clinical purposes during routine healthcare. Secondary use of this data did not require research consent from individuals, and approval for public release of the data followed local governance procedures. Data are publicly available, through DECIPHER (https://www.deciphergenomics.org/). Analyses of these data do not require Research Ethics Committee approval.

---

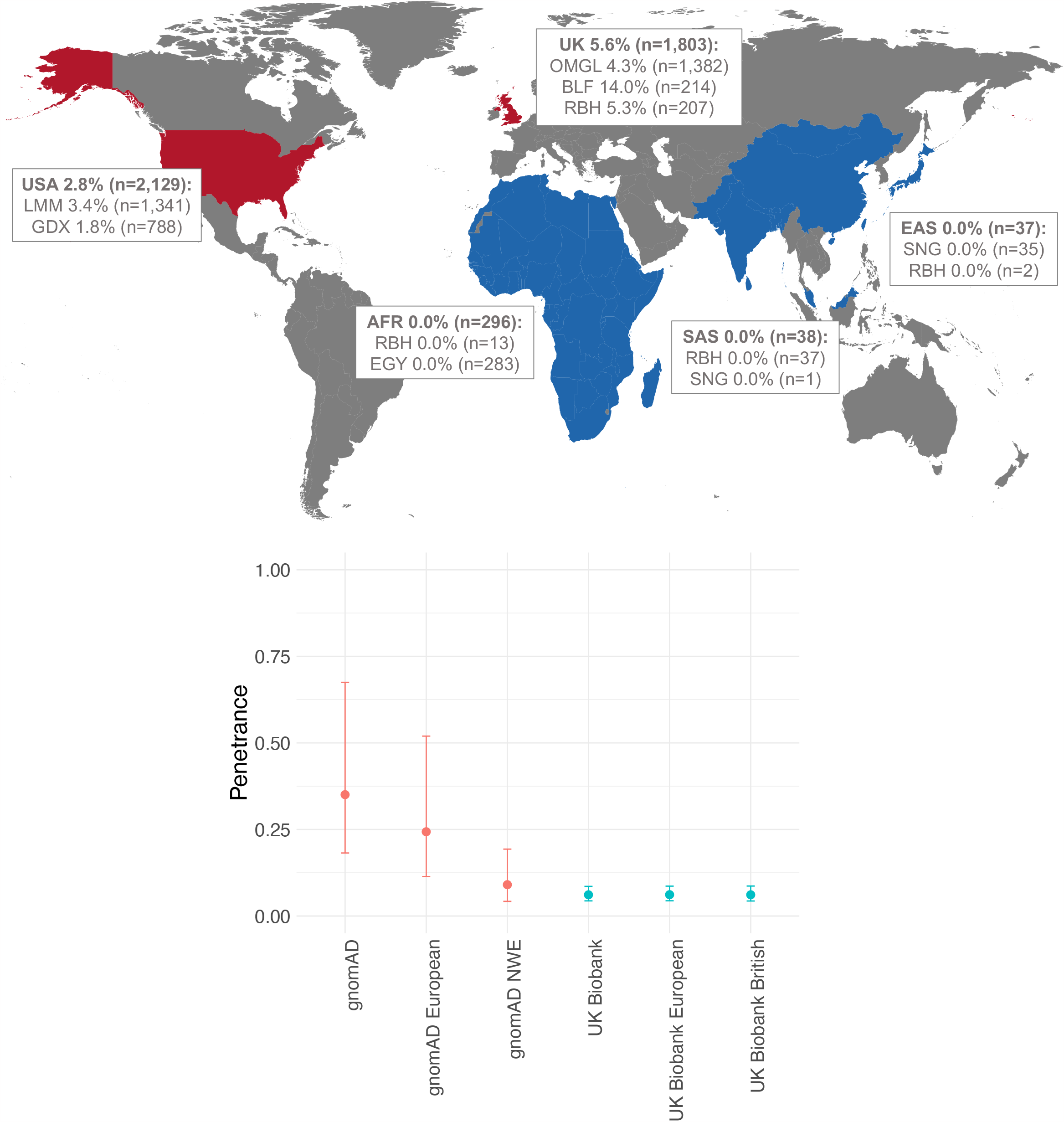

## References

1. Blout Zawatsky, C.L., Bick, D., Bier, L., Funke, B., Lebo, M., Lewis, K.L., Orlova, E., Qian, E., Ryan, L., Schwartz, M.L.B., et al. (2023). Elective genomic testing: Practice resource of the National Society of Genetic Counselors. J. Genet. Couns.

2. Miller, D.T., Lee, K., Abul-Husn, N.S., Amendola, L.M., Brothers, K., Chung, W.K., Gollob, M.H., Gordon, A.S., Harrison, S.M., Hershberger, R.E., et al. (2022). ACMG SF v3.1 list for reporting of secondary findings in clinical exome and genome sequencing: A policy statement of the American College of Medical Genetics and Genomics (ACMG). Genet. Med. 24, 1407–1414.

3. Wilde, A.A.M., Semsarian, C., Márquez, M.F., Shamloo, A.S., Ackerman, M.J., Ashley, E.A., Sternick, E.B., Barajas-Martinez, H., Behr, E.R., Bezzina, C.R., et al. (2022). European Heart Rhythm Association (EHRA)/Heart Rhythm Society (HRS)/Asia Pacific Heart Rhythm Society (APHRS)/Latin American Heart Rhythm Society (LAHRS) Expert Consensus Statement on the state of genetic testing for cardiac diseases. EP Eur. 24, 1307–1367.

4. de Wert, G., Dondorp, W., Clarke, A., Dequeker, E.M.C., Cordier, C., Deans, Z., van El, C.G., Fellmann, F., Hastings, R., Hentze, S., et al. (2021). Opportunistic genomic screening. Recommendations of the European Society of Human Genetics. Eur. J. Hum. Genet. 29, 365–377.

5. Ormondroyd, E., Mackley, M.P., Blair, E., Craft, J., Knight, J.C., Taylor, J.C., Taylor, J., and Watkins, H. (2018). “Not pathogenic until proven otherwise”: Perspectives of UK clinical genomics professionals toward secondary findings in context of a Genomic Medicine Multidisciplinary Team and the 100,000 Genomes Project. Genet. Med. 20, 320–328.

6. McGurk, K.A., Zheng, S.L., Henry, A., Josephs, K., Edwards, M., de Marvao, A., Whiffin, N., Roberts, A., Lumbers, T.R., O’Regan, D.P., et al. (2022). Correspondence on “ACMG SF v3.0 list for reporting of secondary findings in clinical exome and genome sequencing: a policy statement of the American College of Medical Genetics and Genomics. Genet. Med. 24, 744–746.

7. Wilson, J.M.G., and Jungner, G. (1968). Principles and practice of screening for disease: public health papers no. 34 (Geneva: World Health Organization).

8. Wynn, J., Martinez, J., Bulafka, J., Duong, J., Zhang, Y., Chiuzan, C., Preti, J., Cremona, M.L., Jobanputra, V., Fyer, A.J., et al. (2018). Impact of Receiving Secondary Results from Genomic Research: A 12-Month Longitudinal Study. J. Genet. Couns. 27, 709–722.

9. Hart, M.R., Biesecker, B.B., Blout, C.L., Christensen, K.D., Amendola, L.M., Bergstrom, K.L., Biswas, S., Bowling, K.M., Brothers, K.B., Conlin, L.K., et al. (2019). Secondary findings from clinical genomic sequencing: prevalence, patient perspectives, family history assessment, and health-care costs from a multisite study. Genet. Med. Off. J. Am. Coll. Med. Genet. 21, 1100–1110.

10. Thauvin-Robinet, C., Thevenon, J., Nambot, S., Delanne, J., Kuentz, P., Bruel, A.-L., Chassagne, A., Cretin, E., Pelissier, A., Peyron, C., et al. (2019). Secondary actionable findings identified by exome sequencing: expected impact on the organisation of care from the study of 700 consecutive tests. Eur. J. Hum. Genet. 27, 1197–1214.

11. Mackley, M.P., Fletcher, B., Parker, M., Watkins, H., and Ormondroyd, E. (2017). Stakeholder views on secondary findings in whole-genome and whole-exome sequencing: A systematic review of quantitative and qualitative studies. Genet. Med. 19, 283–293.

12. Ormondroyd, E., Harper, A.R., Thomson, K.L., Mackley, M.P., Martin, J., Penkett, C.J., Salatino, S., Stark, H., Stephens, J., and Watkins, H. (2020). Secondary findings in inherited heart conditions: a genotype-first feasibility study to assess phenotype, behavioural and psychosocial outcomes. Eur. J. Hum. Genet. 28, 1486–1496.

13. de Marvao, A., McGurk, K.A., Zheng, S.L., Thanaj, M., Bai, W., Duan, J., Biffi, C., Mazzarotto, F., Statton, B., Dawes, T.J.W., et al. (2021). Phenotypic Expression and Outcomes in Individuals With Rare Genetic Variants of Hypertrophic Cardiomyopathy. J. Am. Coll. Cardiol. 78, 1097–1110.

14. Pirruccello, J.P., Bick, A., Wang, M., Chaffin, M., Friedman, S., Yao, J., Guo, X., Venkatesh, B.A., Taylor, K.D., Post, W.S., et al. (2020). Analysis of cardiac magnetic resonance imaging in 36,000 individuals yields genetic insights into dilated cardiomyopathy. Nat. Commun. 11, 2254.

15. Pirruccello, J.P., Bick, A., Chaffin, M., Krishna, G., Choi, S.H., Lubitz, S.A., Carolyn, Y., Ng, K., Philippakis, A., Ellinor, P.T., et al. (2021). Titin truncating variants in adults without known congestive heart failure. J. Am. Coll. Cardiol. 75, 1239–1241.

16. Castel, S.E., Cervera, A., Mohammadi, P., Aguet, F., Reverter, F., Wolman, A., Guigo, R., Iossifov, I., Vasileva, A., and Lappalainen, T. (2018). Modified penetrance of coding variants by cis-regulatory variation contributes to disease risk. Nat. Genet. 50, 1327–1334.

17. Goodrich, J.K., Singer-Berk, M., Son, R., Sveden, A., Wood, J., England, E., Cole, J.B., Weisburd, B., Watts, N., Caulkins, L., et al. (2021). Determinants of penetrance and variable expressivity in monogenic metabolic conditions across 77,184 exomes. Nat. Commun. 12, 3505.

18. Kingdom, R., and Wright, C.F. (2022). Incomplete Penetrance and Variable ExpressivitylJ: From Clinical Studies to Population Cohorts. Front. Genet. 13, 920390.

19. Glazier, A.A., Thompson, A., and Day, S.M. (2019). Allelic imbalance and haploinsufficiency in MYBPC3-linked hypertrophic cardiomyopathy. Pflügers Arch. - Eur. J. Physiol. 471, 781–793.

20. Roberts, A.M., Ware, J.S., Herman, D.S., Schafer, S., Baksi, J., Bick, A.G., Buchan, R.J., Walsh, R., John, S., Wilkinson, S., et al. (2015). Integrated allelic, transcriptional, and phenomic dissection of the cardiac effects of titin truncations in health and disease. Sci. Transl. Med. 7, 270ra6.

21. Tadros, R., Francis, C., Xu, X., Vermeer, A.M.C., Harper, A.R., Huurman, R., Kelu Bisabu, K., Walsh, R., Hoorntje, E.T., te Rijdt, W.P., et al. (2021). Shared genetic pathways contribute to risk of hypertrophic and dilated cardiomyopathies with opposite directions of effect. Nat. Genet. 53, 128–134.

22. Harper, A.R., Goel, A., Grace, C., Thomson, K.L., Petersen, S.E., Xu, X., Waring, A., Ormondroyd, E., Kramer, C.M., Ho, C.Y., et al. (2021). Common genetic variants and modifiable risk factors underpin hypertrophic cardiomyopathy susceptibility and expressivity. Nat. Genet. 53, 135–142.

23. McGurk, K.A., and Halliday, B.P. (2022). Dilated cardiomyopathy – details make the difference. Eur. J. Heart Fail. 24, 1197–1199.

24. Gail, M.H., Pee, D., Benichou, J., and Carroll, R. (1999). Designing studies to estimate the penetrance of an identified autosomal dominant mutation: Cohort, case-control, and genotyped-proband designs. Genet. Epidemiol. 16, 15–39.

25. Lorenzini, M., Norrish, G., Field, E., Ochoa, J.P., Cicerchia, M., Akhtar, M.M., Syrris, P., Lopes, L.R., Kaski, J.P., and Elliott, P.M. (2020). Penetrance of Hypertrophic Cardiomyopathy in Sarcomere Protein Mutation Carriers. J. Am. Coll. Cardiol. 76, 550– 559.

26. Minikel, E.V., Vallabh, S.M., Lek, M., Estrada, K., Samocha, K.E., Sathirapongsasuti, J.F., McLean, C.Y., Tung, J.Y., Yu, L.P.C., Gambetti, P., et al. (2016). Quantifying prion disease penetrance using large population control cohorts. Sci. Transl. Med. 8, 322ra9.

27. Ackerman, M.J., Priori, S.G., Willems, S., Berul, C., Brugada, R., Calkins, H., Camm, A.J., Ellinor, P.T., Gollob, M., Hamilton, R., et al. (2011). HRS/EHRA Expert Consensus Statement on the State of Genetic Testing for the Channelopathies and Cardiomyopathies. EP Eur. 13, 1077–1109.

28. Bycroft, C., Freeman, C., Petkova, D., Band, G., Elliott, L.T., Sharp, K., Motyer, A., Vukcevic, D., Delaneau, O., O’Connell, J., et al. (2018). The UK Biobank resource with deep phenotyping and genomic data. Nature 562, 203–209.

29. Szustakowski, J.D., Balasubramanian, S., Kvikstad, E., Khalid, S., Bronson, P.G., Sasson, A., Wong, E., Liu, D., Wade Davis, J., Haefliger, C., et al. (2021). Advancing human genetics research and drug discovery through exome sequencing of the UK Biobank. Nat. Genet. 53, 942–948.

30. Karczewski, K.J., Francioli, L.C., Tiao, G., Cummings, B.B., Alföldi, J., Wang, Q., Collins, R.L., Laricchia, K.M., Ganna, A., Birnbaum, D.P., et al. (2020). The mutational constraint spectrum quantified from variation in 141,456 humans. Nature 581, 434–443.

31. McLaren, W., Gil, L., Hunt, S.E., Riat, H.S., Ritchie, G.R.S., Thormann, A., Flicek, P., and Cunningham, F. (2016). The Ensembl Variant Effect Predictor. Genome Biol. 17, 122.

32. Jaganathan, K., Kyriazopoulou Panagiotopoulou, S., McRae, J.F., Darbandi, S.F., Knowles, D., Li, Y.I., Kosmicki, J.A., Arbelaez, J., Cui, W., Schwartz, G.B., et al. (2019). Predicting Splicing from Primary Sequence with Deep Learning. Cell 176, 535–548.

33. Ioannidis, N.M., Rothstein, J.H., Pejaver, V., Middha, S., McDonnell, S.K., Baheti, S., Musolf, A., Li, Q., Holzinger, E., Karyadi, D., et al. (2016). REVEL: An Ensemble Method for Predicting the Pathogenicity of Rare Missense Variants. Am. J. Hum. Genet. 99, 877–885.

34. Landrum, M.J., Lee, J.M., Riley, G.R., Jang, W., Rubinstein, W.S., Church, D.M., and Maglott, D.R. (2014). ClinVar: public archive of relationships among sequence variation and human phenotype. Nucleic Acids Res. 42, D980–5.

35. Purcell, S., Neale, B., Todd-Brown, K., Thomas, L., Ferreira, M.A.R., Bender, D., Maller, J., Sklar, P., de Bakker, P.I.W., Daly, M.J., et al. (2007). PLINK: a tool set for whole-genome association and population-based linkage analyses. Am. J. Hum. Genet. 81, 559–575.

36. Ingles, J., Goldstein, J., Thaxton, C., Caleshu, C., Corty, E.W., Crowley, S.B., Dougherty, K., Harrison, S.M., McGlaughon, J., Milko, L. V., et al. (2019). Evaluating the Clinical Validity of Hypertrophic Cardiomyopathy Genes. Circ. Genomic Precis. Med. 12, 57–64.

37. Jordan, E., Peterson, L., Ai, T., Asatryan, B., Bronicki, L., Brown, E., Celeghin, R., Edwards, M., Fan, J., Ingles, J., et al. (2021). Evidence-Based Assessment of Genes in Dilated Cardiomyopathy. Circulation 144, 7–19.

38. Whiffin, N., Walsh, R., Govind, R., Edwards, M., Ahmad, M., Zhang, X., Tayal, U., Buchan, R., Midwinter, W., Wilk, A.E., et al. (2018). CardioClassifier: disease- and gene-specific computational decision support for clinical genome interpretation. Genet. Med. 20, 1246–1254.

39. Zou, Y., Song, L., Wang, Z., Ma, A., Liu, T., Gu, H., Lu, S., Wu, P., Zhang, Y., Shen, L., et al. (2004). Prevalence of idiopathic hypertrophic cardiomyopathy in China: A population-based echocardiographic analysis of 8080 adults. Am. J. Med. 116, 14–18.

40. Maron, B.J., Spirito, P., Roman, M.J., Paranicas, M., Okin, P.M., Best, L.G., Lee, E.T., and Devereux, R.B. (2004). Prevalence of hypertrophic cardiomyopathy in a population-based sample of American Indians aged 51 to 77 years (the Strong Heart Study). Am. J. Cardiol. 93, 1510–1514.

41. Maron, B.J., Gardin, J.M., Flack, J.M., Gidding, S.S., Kurosaki, T.T., and Bild, D.E. (1995). Prevalence of Hypertrophic Cardiomyopathy in a General Population of Young Adults. Circulation 92, 785–789.

42. Petersen, S.E., Aung, N., Sanghvi, M.M., Zemrak, F., Fung, K., Paiva, J.M., Francis, J.M., Khanji, M.Y., Lukaschuk, E., Lee, A.M., et al. (2017). Reference ranges for cardiac structure and function using cardiovascular magnetic resonance (CMR) in Caucasians from the UK Biobank population cohort. J. Cardiovasc. Magn. Reson. 19, 18.

43. Mestroni, L., Maisch, B., McKenna, W.J., Schwartz, K., Charron, P., Rocco, C., Tesson, F., Richter, A., Wilke, A., and Komajda, M. (1999). Guidelines for the study of familial dilated cardiomyopathies. Collaborative Research Group of the European Human and Capital Mobility Project on Familial Dilated Cardiomyopathy. Eur. Heart J. 20, 93–102.

44. McNally, E.M., and Mestroni, L. (2017). Dilated cardiomyopathy: Genetic determinants and mechanisms. Circ. Res. 121, 731–748.

45. O’Malley, K.J., Cook, K.F., Price, M.D., Wildes, K.R., Hurdle, J.F., and Ashton, C.M. (2005). Measuring diagnoses: ICD code accuracy. Health Serv. Res. 40, 1620–1639.

46. Hershberger, R.E., Hedges, D.J., and Morales, A. (2013). Dilated cardiomyopathy: The complexity of a diverse genetic architecture. Nat. Rev. Cardiol. 10, 531–547.

47. Shah, R.A., Asatryan, B., Dabbagh, G.S., Aung, N., Khanji, M.Y., Lopes, L.R., van Duijvenboden, S., Holmes, A., Muser, D., Landstrom, A.P., et al. (2022). Frequency, Penetrance, and Variable Expressivity of Dilated Cardiomyopathy-Associated Putative Pathogenic Gene Variants in UK Biobank Participants. Circulation 146, 101–124.

48. Lindeboom, R.G.H., Supek, F., and Lehner, B. (2016). The rules and impact of nonsense-mediated mRNA decay in human cancers. Nat. Genet. 48, 1112–1118.

49. Haggerty, C.M., Damrauer, S.M., Levin, M.G., Birtwell, D., Carey, D.J., Golden, A.M., Hartzel, D.N., Hu, Y., Judy, R., Kelly, M.A., et al. (2019). Genomics-First Evaluation of Heart Disease Associated With Titin-Truncating Variants. Circulation 140, 42–54.

50. Saltzman, A.J., Mancini-DiNardo, D., Li, C., Chung, W.K., Ho, C.Y., Hurst, S., Wynn, J., Care, M., Hamilton, R.M., Seidman, G.W., et al. (2010). Short communication: the cardiac myosin binding protein C Arg502Trp mutation: a common cause of hypertrophic cardiomyopathy. Circ. Res. 106, 1549–1552.

51. Whiffin, N., Minikel, E., Walsh, R., O’Donnell-Luria, A.H., Karczewski, K., Ing, A.Y., Barton, P.J.R., Funke, B., Cook, S.A., Macarthur, D., et al. (2017). Using high-resolution variant frequencies to empower clinical genome interpretation. Genet. Med. 19, 1151– 1158.

52. Fry, A., Littlejohns, T.J., Sudlow, C., Doherty, N., Adamska, L., Sprosen, T., Collins, R., and Allen, N.E. (2017). Comparison of Sociodemographic and Health-Related Characteristics of UK Biobank Participants With Those of the General Population. Am. J. Epidemiol. 186, 1026–1034.

53. Mahmood, A., Morris-Rosendahl, D., Edwards, M., Fleming, A., Homfray, T., Mason, S., Quinn, E., Ware, J., Baksi, J., Prasad, S., et al. (2022). 10 Disease penetrance in asymptomatic carriers of familial cardiomyopathy variants. Heart 108, A9 LP–A9.

54. Richards, S., Aziz, N., Bale, S., Bick, D., Das, S., Gastier-Foster, J., Grody, W.W., Hegde, M., Lyon, E., Spector, E., et al. (2015). Standards and guidelines for the interpretation of sequence variants: A joint consensus recommendation of the American College of Medical Genetics and Genomics and the Association for Molecular Pathology. Genet. Med. 17, 405–424.

55. McAfee, Q., Chen, C.Y., Yang, Y., Caporizzo, M.A., Morley, M., Babu, A., Jeong, S., Brandimarto, J., Bedi, K.C., Flam, E., et al. (2022). Truncated titin proteins in dilated cardiomyopathy. Sci. Transl. Med. 13, eabd7287.

56. Fomin, A., Gärtner, A., Cyganek, L., Tiburcy, M., Tuleta, I., Wellers, L., Folsche, L., Hobbach, A.J., von Frieling-Salewsky, M., Unger, A., et al. (2022). Truncated titin proteins and titin haploinsufficiency are targets for functional recovery in human cardiomyopathy due to TTN mutations. Sci. Transl. Med. 13, eabd3079.

