## Supplementary Information for "The penetrance of rare variants in cardiomyopathy-associated genes: a cross-sectional approach to estimate penetrance for secondary findings"

**Table of tables**

**Table of lists**

**Table of figures**

### 1 Supplementary method

#### 1.1 Overview of the estimation of penetrance and its confidence interval

In this study, we adapted the estimate of penetrance from Minikel et al. (2016)^1^

$$P(D|A)=P(D)\frac{P(A|D)}{P(A)},$$

(**Eq. S1**)

where $P(D|A)$ is the penetrance of the variant (by adulthood), *i.e.,* the probability of disease given a risk allele; $P(D)$ is the prevalence of the disease, *i.e.,* the baseline risk in the general population; $P(A|D)$ is the frequency of individuals with the disease who have the allele, *i.e.*, the allele frequency in cases; and $P(A)$ is the frequency of the allele in the general population, *i.e.*, the population allele frequency.

An alternative approach would be to estimate penetrance via a likelihood ratio test, *i.e.*, the probability of disease given a risk allele divided by a positive test. However, this requires healthy controls, *i.e*., the identification of healthy controls instead of population cohorts. This is erroneous without known cardiac status.

In the following, we indicate with $D|A$, $D$, $A|D$ and $A$, the random variables (r.v.s) for the penetrance of the variant, the prevalence of the disease, the allele frequency as a proportion in cases, and the allele frequency as a proportion in the general population, respectively. We indicate with $p_{D|A}\equiv P(D|A)$, $p_{D}\equiv P(D)$, $p_{A|D}\equiv P\left( A | D \right)$ and $p_{A}\equiv P(A)$, the probability of the corresponding events. Finally, we specify with $\pi(D|A$), $\pi(D)$, $\pi(A|D)$ and $\pi(A)$, the distribution of the corresponding (discrete or continuous) r.v.s.

To estimate the confidence interval surrounding the estimate of penetrance, we assessed several methods:

- Minikel *et al.* (2016)^1^ used binomial confidence intervals to estimate the uncertainty regarding the penetrance. The authors estimated the binomial proportion $\left( 1-\alpha\right)\%$ confidence interval for $A|D$ and independently for $A$, divided separately the lower limits (LL) and the upper limits (UL) of the confidence intervals and multiply them by estimated $p_{D}$. In this framework, the penetrance confidence interval could be outside the interval $[0,1]$ (“overshooting”^2^) and was therefore truncated in the interval $[0,1]$.
- We considered using the above estimate of uncertainty and tested other methods proposed in literature for the confidence interval of binomial proportions (*e.g.*, simple asymptotic or Wald method, Wilson score method, etc. see for instance^3,4^ and references therein) and adjusted the nominal level of significance such that the coverage probability aligns with the $(1-\alpha)\%$ nominal level^5^.
- We also wanted to fully estimate the uncertainty surrounding the penetrance estimate. To do this, we aimed to undertake a fully Bayesian approach to estimate the confidence interval for penetrance including an estimate of uncertainty regarding the prevalence of cardiomyopathy described in the literature. In our framework, this was not possible. When a joint beta-binomial model is specified for $\pi(D|A) \propto\pi(A|D)\pi(D)$, where $A|D$ follows a binomial distribution with the probability of success $\pi(D)$, and $D$ is distributed as a beta density, the marginal distribution $\pi\left( A \right)$ is given^6^. Thus, a Bayesian approach cannot be used to quantify the uncertainty of penetrance. In our cross-sectional approach, $A$ is assumed independent from $A|D$ and follows a binomial distribution, whereas from a Bayesian perspective, $\pi(A)$ is derived by marginalizing out $D$ form the joint distribution $\pi(A,D)$, *i.e.*, $\pi(A)=\int\pi(A|D)\pi(D)dD$.
  For comparison, we plotted (see **Figure S5**) the beta-binomial distribution derived from the marginalization of the joint distribution against the corresponding binomial distribution assuming $A$ and $A|D$ are independent.
- We also tested a Monte Carlo approach to overcome the problem of the fully Bayesian formulation by using an inverse logit transformation of a normal distribution as the prior density for $D$, while retaining the above specification for $\pi(A|D)$ (binomial distribution with probability of success $\pi(D)$) and $\pi(A)$ (binomial distribution) and sampled independent realisations from $(D,A|D)$ and $A$to derive the $(1-\alpha)\%$ Monte Carlo confidence interval for penetrance. To avoid overshooting, each realisation of the Monte Carlo simulation was checked and, if necessary, truncated in the interval $[0,1]$.
- Our final approach, and the approach used here, was to assume the independence of the r.v.s $D$, $A|D$, and $A$, to derive the $\left( 1-\alpha\right)\%$ confidence interval for penetrance as the product and ratio of binomial proportions. Our method of choice used the specialised version of the Central Limit Theorem, the Delta method^7^ on the log-transformed random variable $\log\left( D | A \right)=\log\left( D \right)+\log\left( A | D \right)-\log\left( A \right)$ with an improved mean approximation and adjustment for degeneracy^3^. The parameterisation of the binomial distribution $\pi(D)$ was derived from a meta-analysis of literature-based estimates of the prevalence of HCM, while UK Biobank CMR-derived estimate of $p_{D}$ was used in the penetrance equation for DCM where few published studies were available for inclusion in the meta-analysis.

#### 1.2 Estimation of penetrance

Following Minikel *et al.* (2016)^1^, penetrance is defined as the probability of developing disease given a risk allele $p_{D|A}$ and can be estimated by Bayes’ rule (**Eq. S1**). Three parameters were used to define penetrance by adulthood: i) the prevalence $p_{D}$of the disease, *i.e.*, the baseline lifetime risk in the general population, ii) the proportion $p_{A|D}$ of individuals with the disease who have the allele, *i.e.*, the allele frequency in cases, and iii) the frequency $p_{A}$ of the allele in the general population, *i.e.*, the population allele frequency. The allele frequency is used in $p_{A|D}$ and $p_{A}$ and it is estimated as the probability $p$ of success in a binomial experiment by using the allele counts $x$, *i.e.*, the binomial number of successes, and allele number $n$, *i.e.*, the binomial number of trials. We estimate the penetrance of an allele under a dominant genetic model^8^ as

$$p_{D|A}=p_{D}\frac{p_{A|D}}{p_{A}}=\frac{x_{D}x_{A|D}}{x_{A}}\frac{n_{A}}{n_{D}n_{A|D}}.$$

(**Eq. S2**)

The penetrance $p(D|A)$ of an allele is estimated using three parameters: $p_{D}$, the fixed probability of disease calculated by meta-analysis of reported prevalence of disease from literature (with $x_{D}$, allele count, and $n_{D}$, the allele measure, both estimated, see below), $p_{A|D}$, the probability of the allele given disease, estimated from allele frequency in cases (with $x_{A|D}$ and $n_{D|A}$ observed), and $p_{A}$, the probability of the allele, estimated from the allele frequency in population cohorts (with $x_{A}$ and $n_{A}$ observed).

#### 1.3 Probability of the disease: cardiomyopathy prevalence estimates

The prevalence of cardiomyopathy has been previously estimated and reported as the most simplified ratio of 1 in 500 for hypertrophic cardiomyopathy (HCM) and 1 in 250 for dilated cardiomyopathy (DCM)^9^. To identify the true confidence with our current knowledge of the prevalence of cardiomyopathy, a literature review was undertaken to identify population-based prevalence estimates of cardiomyopathy (**Table S1**).

For the prevalence of DCM, 12 cohorts were identified from literature^10–19^ (**Figure S1**). We have previously found the use of cardiac imaging to have higher sensitivity in estimating cardiomyopathy prevalence than ICD codes^20^. Only one article used imaging in identifying DCM prevalence^17^. We therefore assessed the prevalence of clinical DCM (LVEDV > 232ml in males and > 175ml in females, plus LVEF < 50%, in the absence of a record of CAD or HCM) in the imaging tranche of the UK Biobank^21^. 177 DCM cases were identified from cardiac imaging of 39,003 participants ($p_{D}$ = 0.45% (binomial 95% $\mathrm{CI}_{D}=$ 0.39%-0.53%) or 1 in 220)^22^. As a meta-analysis cannot be undertaken with only two cohorts, we were restricted to using the UK Biobank estimate only, which is similar to the expected DCM prevalence of 1 in 250^9^.

For the prevalence of HCM, 22 cohorts were identified in literature^12,13,28–36,14,15,20,23–27^ (**Figure S2, Table S2**). As expected, a combined meta-analysis of all 22 cohorts identified from literature showed high heterogeneity (*P*-value = 0, heterogeneity index *I*^2^ = 100%). Four articles used cardiac imaging in identifying HCM prevalence^20,25,26,37^. A meta-analysis of the binomial proportions was undertaken using the *meta*^38^ and *metafor*^39^ R packages. This resulted in an estimated $p_{D}$ of 0.18% (95% $\mathrm{CI}_{D}=$ 0.15%-0.23%) (**Figure S3**).

From the meta-analysis estimate of $p_{D}$ and its confidence interval, we derived the values for $x_{D}$ and $n_{D}$ (solving two unknown values in two equations, one describing the estimation of $p_{D}$ and the other, its confidence interval). However, since several ways to estimate the confidence interval for binomial proportions have been proposed in literature^3,4^, different values of $x_{D}$ and $n_{D}$ can also be obtained. We assessed four popular methods: the Wald method, based on a simple asymptotic normal approximation (**Eq. S4**), the Arcsine method, based on the Delta method for variance stabilization using the $\sin^{-1} \sqrt{p_{D}}$ transformation (**Eq. S5**), the Agresti-Coull method^40^, which relies on the asymptotic normal approximation centred in

$$\tilde{p}_{D}=\frac{\hat{p}_{D}+\frac{z_{1-\alpha/2}^{2}}{2n_{D}}}{1+\frac{z_{1-\alpha/2}^{2}}{n_{D}}},$$

(**Eq. S3**)

where $\hat{p}_{D}$ is the meta-analysis estimate of $p_{D}$ and $z_{1-\alpha/2}$ is the $1-\alpha/2$ quantile of the standard normal distribution (**Eq. S6**), and the Clopper-Pearson method, an exact method for the confidence interval of a binomial proportion (**Eq. S7**).

The best method chosen was the one that minimizes the Euclidian distance between the meta-analysis estimates $(\hat{p}_{D},{\hat{\mathrm{LL}}}_{D}, {\hat{\mathrm{UL}}}_{D})$, where ${\hat{\mathrm{LL}}}_{D}$ and ${\hat{\mathrm{UL}}}_{D}$ are the lower and upper limit of the $(1-\alpha$)% confidence interval for the prevalence of the disease, and $({\hat{\hat{p}}}_{D},{\hat{\hat{\mathrm{LL}}}}_{D}, {\hat{\hat{\mathrm{UL}}}}_{D})$, i.e., the same quantities estimated by each method after the corresponding estimates of $x_{D}$ and $n_{D}$ are obtained. At $\alpha=0.05$, given the results of the meta-analysis, the Agresti-Coull method performed the best with the lowest L_2_ norm and with low relative errors, defined as the relative difference between the meta-analysis values and their estimated values calculated by each method (**Table S3**). See **List S1** for details. This derived $x_{D}$= 97 and $n_{D}=$52,660 for HCM.

Using the same methods and included studies, we derived estimates for male- and female-specific CM prevalence. For DCM, population prevalence was estimated as ~1 in 340 females ($p_{D}=$0.30% (95% $\mathrm{CI}_{D}=$0.23%-0.38%); $x_{D}$= 60 in 20,316) and ~1 in 160 males ($p_{D}=$ 0.63% (95% $\mathrm{CI}_{D}=$0.52%-0.75%);$x_{D}$= 117 in $n_{D}=$18,687). For HCM, population prevalence was estimated as ~1 in 1,300 females ($p_{D}=$ 0.08% (95% $\mathrm{CI}_{D}=$0.04%-0.12%); $x_{D}$= 15 in $n_{D}=$19,646) and ~1 in 360 males ($p_{D}=$ 0.28% (95% $\mathrm{CI}_{D}=$0.22%-0.35%); $x_{D}$= 68 in $n_{D}=$24,411). To estimate the penetrance of variants by age, the prevalence of disease was adjusted for the proportion of total cases that were measured by each decade and it was assumed that the population allele frequency is fixed.

#### 1.4 Probability of the allele given disease: allele frequency in the case cohort

The allele frequency of variants in the case cohort was used in the penetrance calculation for $p_{D|A}$. The allele count and allele number were used for $x_{A|D}$ and $n_{A|D}$, respectively. See **Section 1.10** for further information.

#### 1.5 Probability of the allele: allele frequency in the population reference datasets

The allele frequency of variants in the combined population cohort of UK Biobank and gnomAD was used in the penetrance calculation for $p_{A}$. The allele count and allele number were used for $x_{A}$ and $n_{A}$, respectively. It is assumed that the population datasets include individuals who will later die of cardiac disease, thus enabling direct use of the gnomAD and UK Biobank allele frequencies combined as $p_{A}$. See **Section 1.9** for further information.

#### 1.6 Confidence intervals

Since it is not possible to undertake a fully Bayesian analysis to estimate the confidence interval for penetrance, we used a different approach; the specialised version of the Central Limit Theorem, the Delta method^7^, on the log-transformed random variable $\log\left( D | A \right)=\log(D)+\log\left( A | D \right)-log(A)$ (**Eq. S1**), assuming the independence between the binomial random variables $A|D$, $A$ and $D$, with an improved mean approximation and adjustment for degeneracy^3^. The Delta method concerns the approximate distribution of a function of random variables which is asymptotically normal where the mean and variance are obtained by a first-order Taylor approximation expanded around the means. The improved mean involves a better approximation of the first moment of the asymptotic normal distribution by using a second-order Taylor expansion. To address the problem of degeneracy, *i.e.*, the confidence interval’s width is 0 when the probability of success is 0, we added the constant $d=d_{x}=d_{n}=0.5$ to all $x$ and $n$, respectively^41,42^, as the allele frequencies $p_{A}$ and $p_{A|D}$ of rare variants will always tend towards zero.

We compared this approach with seven other methods for deriving the confidence intervals of penetrance (**List S2**). In the first group of methods (G1), we derived the confidence interval for penetrance as the $(1-\alpha$)% confidence interval of the ratio of binomial proportions $A|D$ and $A$, similar to the derivation of confidence intervals for the relative risk^43^, and multiplied it by the estimated value of $p_{D}$. In the second group (G2), we considered $D$as a random variable subject to uncertain quantification. We assessed the methods in groups G1 and G2 using an example variant with the following parameters: 97/52,660 for $x_{D}/n_{D}$ obtained from the HCM meta-analysis, 10/20,000 for $x_{A|D}/n_{A|D}$ and 3/600,000 for $x_{A}/n_{A}$. For all methods, we tested with degeneracy adjustment or by adding a continuity correction^3^. Our method of choice (part F in **List S2**) fully encompasses the uncertainty regarding $p_{D}$ (**Figure S4**). An example of a Bayesian approach where the prior and posterior have little overlap is depicted (**Figure S5**).

#### 1.7 Statistical power simulations

To assess how the model responds in different test cases based on allele frequency for $p_{A}$ and $p_{A|D}$, four sets of simulations were undertaken. Firstly, an assessment of the sample size required for $p_{A|D}$ and $p_{A}$ was undertaken to define the penetrance estimate (**Figure S6**). Example variants that had ~10%, ~20%, ~50%, and ~75% estimated penetrance. In each example, smaller $n_{A}$ and $n_{A|D}$ had larger CIs, however the difference was negligible in the ranges of, and greater than, the $n_{A}$ and $n_{A|D}$ used in this project. Thus, the $n_{A}$ from current publicly available data (gnomAD (n = 126,000) and UKB (n = 200,000)) and the $n_{A|D}$ in our HCM case series of 10,000 participants, provides enough precision to estimate penetrance of variants.

Secondly, increasing case samples through international effort provides negligible gain in confidence surrounding the penetrance estimates (**Figure S7**). Increases in publicly available population datasets provides a substantial gain in confidence (**Figure S8, Figure S9**). Thirdly, as the penetrance equation becomes unbalanced (through increased $p_{A|D}$ and decreased $p_{A}$ ratio) and a variant is more penetrant, the confidence intervals increase.

Fourthly, the model was used to assess the expected penetrance results in the range of $p_{A}$ and $p_{A|D}$ observed in this study, using the minimum, median, and maximum allele frequencies (**Figure S11**). Observations were excluded where $x_{A|D}$or $x_{A}$ were <= 1. For the maximum $p_{A|D}$, identification of a highly penetrant variant (*i.e.*, $p_{A|D}$ = 0.008, $p_{A}$<2×10^6^) resulted in a penetrance and 95% CI > 1. At the median and minimum $p_{A|D}$ (0.0003 and 0.0001, respectively), all variants had a penetrance of <25%, with the 95% CI increasing with increase in$p_{A}$. For all $p_{A|D}$, variants with $p_{A}$ > 0.00001 have a penetrance estimate that tends to 0, similar to the concept of filtering allele frequency^44^.

#### 1.8 Estimation of penetrance and confidence intervals as an R language script

```
penetrance <- function(x_D, n_D, x_AgD, n_AgD, x_A, n_A)
{
  set.seed(28061971)
  digits <- 6
  alpha <- 0.05
  p_D <- x_D / n_D
  p_AgD <- x_AgD / n_AgD
  p_A <- x_A / n_A
  d <- 0.5
  p_D_wod <- (x_D + d) / (n_D + d)
  p_AgD_wod <- (x_AgD + d) / (n_AgD + d)
  p_A_wod <- (x_A + d) / (n_A + d)
  log_AR <- log(p_D_wod * p_AgD_wod / p_A_wod) +
            1/2 * ((1 / p_A_wod) * (1 - p_A_wod) / (n_A + d) -
            (1 / p_D_wod) * (1 - p_D_wod) / (n_D + d) -
            (1 / p_AgD_wod) * (1 - p_AgD_wod) / (n_AgD + d))
  Var_log_AR <- (1 / p_D_wod) * (1 - p_D_wod) / (n_D + d) +
                (1 / p_AgD_wod) * (1 - p_AgD_wod) / (n_AgD + d)
                +(1 / p_A_wod) * (1 - p_A_wod) / (n_A + d)
  log_LCI <- log_AR - qnorm(1 - alpha / 2) * sqrt(Var_log_AR)
  log_UCI <- log_AR + qnorm(1 - alpha / 2) * sqrt(Var_log_AR)
  penetrance <- pmin(1, pmax(0,exp(log_AR)))
  log_lCI <- pmin(1, pmax(0,exp(log_LCI)))
  log_uCI <- pmin(1, pmax(0,exp(log_UCI)))
  my_list <- list("penetrance" = penetrance, "lci" = log_lCI,
                  "uci" = log_uCI)
  return(my_list)
}
```

See also https://github.com/ImperialCardioGenetics/variantfx/tree/main/PenetrancePaper

#### 1.9 Population reference cohort summary information

The UK Biobank (UKBB) recruited 500,000 participants aged 40–69 years across the United Kingdom between 2006 and 2010 (National Research Ethics Service - 11/NW/0382)^45^. This study was conducted under terms of access approval number 47602. Written informed consent was provided. UKBB participants underwent whole exome sequencing (WES) as previously described^46^. The WES data is in GrCh38 and left-aligned. Participants that had withdrawn were excluded from the analysis. The maximal subset of unrelated participants was used, identified by those included in the UKBB PCA analysis (S3.3.2^47^; QCed). Two sets of data were created, a dataset representing the whole QCed cohort and a dataset representing genetically white British individuals only (NWE). 167,478 participants remained, of which 137,998 were genetically white British, mean age of 56 years old at recruitment, 75,727 were male, and 91,751 were female.

The Genome Aggregation Database (gnomAD) is the result of a coalition effort to aggregate and harmonize exome sequencing data from a variety of large-scale sequencing projects^48^. The version 2.1 short variant dataset spans 125,748 exomes from unrelated individuals sequenced as part of various disease-specific and population genetic studies and lifted over to GrCh38. 57,787 were female and 67,961 were male. Ancestry is provided for global super-populations: *i.e.*, African/African American (AFR), American Admixed/Latino (AMR), East Asian (EAS), Non-Finnish European (NFE), and South Asian (SAS), and some sub populations such as Northwestern Europeans (NWE). Individuals known to be affected by severe paediatric disease have been removed, as well as their first-degree relatives, however, some individuals with severe disease may still be included in the data sets, albeit likely at a frequency equivalent to or lower than that seen in the general population. The data released by gnomAD are available free of restrictions under the Creative Commons Zero Public Domain Dedication. The aggregation and release of summary data from the exomes collected by the Genome Aggregation Database has been approved by the Partners IRB (protocol 2013P001339, "Large-scale aggregation of human genomic data"). The gnomAD dataset was incorporated into the analysis through the Ensembl Variant Effect Predictor^49^ plugin.

#### 1.10 Cardiomyopathy case cohort summary information

Datasets created in closely collaborating centres (described below – RBHT, NHCS, AHCE) of which access has been granted for sequencing BAM files, are denoted “internal datasets”. Datasets summarised and aggregated by external sequencing centres (described below – OMGL, LMM, BRGL, GDx) of which only summary counts were provided, are denoted “external datasets”.

*Internal datasets*

Royal Brompton and Harefield NHS Foundation Trust, London, UK (RBHT) provided panel sequencing on HCM and DCM diagnosed patients, as previously published^50–52^. The patients were identified by consecutive referrals to the imaging unit from the dedicated cardiomyopathy service and a network of 30 regional hospitals, forming the National Institute for Health Research Biobank. Patients were referred for diagnostic evaluation, family screening, or assessment of CM severity. All patients were prospectively enrolled for research purposes and underwent cardiac phenotyping with either cardiovascular magnetic resonance (CMR) or trans-thoracic echocardiography, with CM diagnosed according to standard criteria^50^. Further information regarding the inclusion criteria of the patients, targeted sequencing protocol, and data quality control, can be found in previously published articles^50^. All participants gave written informed consent, and the study was approved by the relevant regional research ethics committees. Samples were sequenced on the NextSeq 500, the MiSeq and the HiSeq Illumina platforms using the TruSight Cardio Sequencing Kit from Illumina (which includes 174 genes associated with inherited cardiac conditions (ICCs)). Additional samples were sequenced on the 5500xl SOLiD platform (SLD) from Life Technologies using a custom Agilent SureSelect panel of genes associated with ICCs.

National Heart Centre Singapore (NHCS), Singapore, provided panel sequencing on HCM and DCM patients via the NHCS Biobank, as previously published^50,51,53^. Patients were sequenced using the Illumina TruSight Cardio targeted panel. All patients were prospectively enrolled for research purposes and underwent cardiac phenotyping with either cardiovascular magnetic resonance (CMR) or transthoracic echocardiography, with cardiomyopathy diagnosed according to standard criteria.

Aswan Heart Centre, Egypt (AHCE) provided panel sequencing on HCM and DCM patients^54,55^. A series of Egyptian patients with CM were assessed at Aswan Heart Centre (AHC) by echocardiography and/or magnetic resonance imaging. Patients were sequenced using the Illumina TruSight Cardio targeted panel on the Illumina MiSeq or NextSeq platforms.

All samples included in the internal datasets were consolidated and joint-genotyped using GATK v4.1.9 GenomicsDBImport and GenotypeGVCFs. Variant calls were hard filtered using GATK Best Practises guidelines for germline short variant discovery. Particularly, variants with quality-by-depth (QD)<3 and read depth <10x were not included in our counts due to the high likelihood of being false positives. All variants were converted to biallelic using bcftools v1.10.2 (htslib 1.10.2) and variants with AC=0 and star (*) alternative alleles were discarded.

*External datasets*

Laboratory of Molecular Medicine, Partners HealthCare, Massachusetts, US (LMM) provided aggregated summary sequencing information on patients with reported cardiomyopathy and consecutive diagnostic referrals for clinical genetic testing, *i.e.*, HCM and DCM (no phenotypic confirmation), as previously published^50,51,56–58^. The LMM HCM cohort comprised unrelated probands referred for HCM clinical genetic testing^59^. Any individuals with an unclear clinical diagnosis of HCM, or with left ventricular hypertrophy due to an identified syndrome such as Fabry or Danon disease, or unaffected individuals with a family history of HCM were excluded. The LMM DCM cohort comprised individual probands referred for DCM clinical genetic testing. According to the published report, all patients had DCM or clinical features consistent with DCM based on the medical and family history information provided by ordering providers. Additionally, any cases with confirmed diagnoses of other cardiomyopathies, structural heart disease, congenital heart disease or syndromic or environmental causes were not included in the study. Only rare variants were included in the aggregated data. Briefly, various sequencing technologies were used across time (Sanger; targeted next-generation sequencing) but with complete coverage (Sanger used to fill gaps in NGS). The LMM2 dataset is a small subset of the LMM cohort that contains ancestry information for the reported variants.

Oxford Molecular Genetics Laboratory, Oxford University Hospitals NHS Foundation Trust, Oxford, UK (OMGL), provided aggregated summary sequencing data on HCM and DCM apparently unrelated patients that were referred from Clinical Genetics centers across the UK for clinical genetic testing with initial clinical diagnosis of HCM or DCM made by a consultant cardiologist. The data included in this analysis is previously published^51,58^. All samples received for diagnostic genetic testing of HCM or DCM genes were eligible and analysis was undertaken in a routine clinical setting using clinical consent.

Belfast Regional Genetics Laboratory, Belfast, UK (BRGL), provided aggregated summary sequencing data on HCM diagnosed patients that had been referred for a Sanger screen. They provided information on four genes, including *TNNI3* of which only information on exons 7 and 8.

GeneDx, Maryland, US (GDx), provided aggregated summary sequencing data on HCM diagnosed patients using panel data between 2016-2017. The data included information on referrals for full panel sequencing. To our knowledge, GDx do not perform further analysis to rule out unrecognised relatedness.

See summary information for the number of participants analysed for each gene of interest (**Table S4**). Actual numbers of samples included in the case cohort varies by gene. The number reported represents the maximum number of samples sequenced across for any gene. Institutional review board–approved protocols were used in this study and all included patients provided written, informed consent for their data to be included in research.

*Ancestry, age at scan and sex*

For the internal cohorts of RBHT, NHCS, AHCE and the LMM2 cohort, ancestry was determined via self-report at sample recruitment (**Table S5**). Local ancestry codes were assigned to one of the eight population codes used in gnomAD to allow ancestry matching across all cohorts. Age at scan was recorded and used in all age-based analyses for the case cohorts. Sex was self-reported at recruitment for all internal datasets, except NHCS. GDx provided age at scan and sex information only for the variants that were reported.

#### 1.11 Data merging

*Technical differences and curation of aggregated datasets*

The datasets included in this study have intra sequencing technology differences, e.g., Illumina and SOLiD technology have separate filtering, inter sequencing technology differences, e.g., NextSeq has higher resolution and depth than HiSeq and MiSeq, and intra panel differences, e.g., WES or target panel which vary in depth (i.e., WES has lower depth) and coverage. The NHCS provided data that was pre-filtered on bam level to a conservative quality of reads which reduced the number of reads.

The external data was shared in multiple different formats (e.g., excel, text, tab- or comma-separated values) with different variant identifiers (HGVSc or genomic position). All variants were confirmed and harmonised to variant call format (vcf) genomic coordinates using VEP v104, and bcftools v1.10.2 (htslib 1.10.2) was used to normalize variants (left align and parsimonious). Quality control or pre-filtering to the reported variants of the external datasets prior to this was subjective to the genetic centres.

#### 1.12 Variant curation

All data (case cohort aggregated data, gnomAD, and UKB) was analysed in GrCh38. The aggregated data of the case cohorts was lifted over from GrCh37 using Picard Tools (version 2.23.1). The resulting vcf file was annotated using Ensembl Variant Effect Predictor (VEP; version 105)^49^ with plugins and additional data for ClinVar (version 20220115)^60^, gnomAD (version r2.1)^48^, SpliceAI (1.3.1)^61^, REVEL^62^, and LOFTEE^48^. The VEP output was analysed using R (version 4.1.2) and Rstudio. The UKBB WES data was incorporated into the analysis using the --frq and --frq counts file formats from PLINK (version 1.9)^63^. Variants identified in the gnomAD data as AC0 (AC=0) were set as missing in the analyses and therefore could only be assessed using the UKBB WES data. The aggregate frequency and count data from gnomAD and UKB were summarised in an additive manner.

Variants identified in the case cohorts were analysed. MANE, protein altering variants of genes of interest that had a MAF of < 0.1% in gnomAD and UKBB were identified. Protein altering variants were included if specified as high or moderate impact by Sequence Ontology^64^ and ENSEMBL^65^, with the addition of splice region variants for further curation. The genes of interest represent a list of 8 sarcomere-encoding genes with definitive evidence of an association with HCM (*MYBPC3, MYH7, MYL2, MYL3, TNNI3, TNNT2, TPM1, ACTC1*)^66^ and 11 genes with definitive or strong evidence of an association with DCM (*BAG3, DES, LMNA, MYH7, PLN, RBM20, SCN5A, TNNC1, TNNT2, TTN, DSP)*^67^. *FLNC* was not included in this study as it was not present on the clinical panels analysed in the case cohort. Analysis was restricted to robustly disease-associated variant classes for each gene: all PAVs of *MYBPC3*; non-truncating variants (non-tvs; inframe indels, missense variants, start/stop lost variants, and nonsense-mediated decay incompetent premature termination codons (NMDi-PTCs)) for the other 7 HCM-associated genes^20^ (*MYH7, MYL2, MYL3, TNNI3, TNNT2, TPM1, ACTC1*); all PAVs for *BAG3, LMNA, PLN, RBM20, SCN5A,* and *DSP*; TTNtvs (cardiac PSI >90%^52^); non-tvs in *DES* and *MYH7*; and missense variants only in *TNNC1* and *TNNT2*.

Splice region variants (in the region of the canonical splice donor and acceptor sites, within 1-3 bases of the exon or 3-8 bases of the intron) with a non-protein altering flag (*i.e.,* synonymous and intron variants) that would otherwise be excluded were assessed in a number of ways; via ClinVar report: those found pathogenic or likely pathogenic with at least 2 star evidence for HCM and DCM in ClinVar and reported functional evidence for splicing were termed “splice confirmed” or if the functional evidence was unclear for splicing were termed “splice likely”; via prediction threshold: the remaining variants were included in the analysis met a recommended SpliceAI threshold for “high precision” of > 0.8. For *TTN*, splice region, missense variants were analysed by Splice AI to identify those variants predicted to cause splicing that would otherwise be excluded.

LOFTEE was incorporated in the analysis to exclude loss of function (LoF) variants that were flagged as “low confidence” (LC) such as “NAGNAG site” requiring reannotation to non-LoF variant status and removal of 5’UTR and 3’UTR splice variants. Essential splice variant LoF occurs in the UTR of the transcript. Additional positional annotation included nonsense-mediated decay (NMD), to identify variants that introduce protein-truncating variants (PTCs) that are insensitive to NMD. The NMD plugin in VEP was used to identify stop-gained variants that are predicted to be NMD incompetent. Additionally, LoF variants found i) 55bp into the penultimate exon^68^ or the whole penultimate exon if shorter than 55bps, ii) in intronless transcripts (a single exon gene, *i.e.,* *PLN*), and iii) frameshift variant where the PTC is predicted to fall 55bp of the penultimate exon or in the final exon. Furthermore, variants flagged “coding sequence variant” or “protein altering variant” were manually curated, as were “stop_lost” and “start_lost” which were examined via ENSEMBL sequence and UCSC Genome Browser^69^ to identify in-frame rescues nearby. Where there was no obvious rescue to assess, the variant was denoted as “inframe insertion”.

Variants were classified as pathogenic/likely pathogenic (P/LP) if reported as P/LP for the correct CM multiple times in ClinVar and confirmed by manual review, or if annotated as P/LP according to ACMG criteria, using the semi-automated CardioClassifier decision support tool^70^ (similar curation previously published^20^). We note the duplication of definitive evidence for *MYH7* and *TNNT2* for both HCM and DCM, variants in these genes were treated as having a role in either HCM or DCM.

Additional allele frequency filtering was used to adjust for potential pre-filtering undertaken for the external datasets: the HCM cohorts (case and population) were filtered to include variants that have a MAF less than the maximum population AF (gnomAD and UKBB) of the external datasets (of which GDx and OMGL had the most filtering, and lowest maximum population allele frequency, for HCM and DCM, respectively). This was a MAF <0.00036598 in gnomAD and MAF <0.0007344 in UKBB for HCM (via GDx) and a MAF <0.000552987 in gnomAD and MAF <0.0006031 in UKBB for DCM (via OMGL). This dataset made up the total variants depicted in this study (**Table S8, Table S9**). To estimate penetrance, only variants that were observed more than once in both the case cohort and population reference dataset were included in the analysis (**Table S10, Table S11**).

For aggregate penetrance estimates of all rare cardiomyopathy variants by subgroup, the UKBB WES data underwent the same variant curation pipeline and filtering thresholds.

### 2 Supplementary results

*Aggregate penetrance by sex and age*

Estimates of the penetrance of rare variants in CM-associated genes by sex and age were undertaken in a subgroup of the HCM and DCM case cohorts where data on reported sex or age was available. Thus, it is not directly comparable to the aggregate penetrance analyses of all samples, although not statistically different. For example, the aggregate penetrance of rare variants in HCM- and DCM-associated genes when calculated in the subgroup of cases with reported sex information available, and using the UK Biobank as reference population, was 27.3% (20.9%-35.5%) for HCM pathogenic variants and 21.6% (10.7%-43.9%) for DCM pathogenic variants. The same for the subgroup of cases with age information available, was 28.3% (21.7%-36.8%) for HCM pathogenic variants and 21.6% (10.4%-45.1%) for DCM pathogenic variants.

*Aggregate penetrance of variant consequences*

The penetrance estimates for specific variant consequences had many notable findings: i) NMDi PTCs (nonsense-mediated decay incompetent premature termination codons or predicted loss of function or truncating variants), NMDc PTCs (NMD competent), and variants expected to lead to splicing in *MYBPC3, BAG3*, *DSP*, and *LMNA*, were most penetrant, ii) pathogenic *TNNT2* inframe deletions, found in abundance in CM cases but absent from reference cohorts, drove an increased penetrance signal for both HCM and DCM, and iii) TTNtvs had estimated penetrance of <20%. The specific inframe deletions in *TNNT2* that caused the “other protein altering variant” subgroup of *TNNT2* variants to have high penetrance for disease were: the variant TNNT2:c.659_661del, identified in 28 DCM cases (1% total cases; 3% G+ cases; 89% have EUR ancestry) and the variant TNNT2:c.517_519del, identified in 15 HCM cases (0.1% total cases; 0.4% G+ cases; 100% EUR ancestry). REVEL software (threshold of 0.75) predicted significantly different penetrance between missense variants in *MYH7, MYL3, TNNI3, TPM1, LMNA*, and *RBM20*, and SpliceAI software (threshold of 0.80) predicted significantly different penetrance between splice variants in *MYBPC3* and *DSP*.

*Simulations*

The simulations showed that penetrance estimates for highly penetrant variants (e.g., >50% penetrance) have large confidence intervals**.** However, if a variant has *at least* 10% population penetrance (via the lower bound of the confidence interval), it is unlikely that the carrier will be released from future clinical follow up. For variants with a more modest estimated penetrance (e.g., <50%), we show that we are now able to estimate penetrance more confidently for variants likely to be identified as secondary findings.

The rate of change of the “error” to the limit confirmed that the gain in confidence from increasing case samples is negligible (the plot plateaus) but increases in future population participants would provide a substantial gain in confidence surrounding the penetrance estimates.

We assessed the size of the confidence interval when varying population allele frequency and case allele frequency. As described by the penetrance equation through the ratio of $P(A|D)/P(A)$ and observed in the simulations, the rarer the variant is in the population (e.g., observed twice in 300,000 participants) and the more common the variant is in the case cohort, the larger the confidence interval. The penetrance equation promotes the increase of the confidence interval in such cases when the penetrance is high due to the unbalanced allele frequency between the smaller case cohort and very large population cohort. In addition, through assessment of simulations within the allele frequency ranges of the variants observed in this study, variants with a very high penetrance and can have an estimated penetrance of >100%. While theoretically this could be the case, we did not observe any real variants in our dataset that had a combination of case and population allele frequencies that resulted in an estimated penetrance of >100% (maximum penetrance was 66.8% for HCM, 78.6% for DCM). Such variants are unlikely to be observed several times in the population reference cohort.

### 3 Supplementary tables

Table S1 Articles assessed in literature review of the prevalence of DCM.

See excel file.

Table S2 Articles assessed in literature review of the prevalence of HCM.

See excel file.

Table S3 Selection of the Agresti-Coull method and comparison of binomial proportion methods for deriving parameters from the meta-analysis results.

${\hat{\boldsymbol{x}}}_{\boldsymbol{D}}$ and ${\hat{\boldsymbol{n}}}_{\boldsymbol{D}}$ were derived from the meta-analysis results ${\hat{\boldsymbol{p}}}_{\boldsymbol{D}}$, $\hat{\mathbf{LL}}$and $\hat{\mathbf{UL}}$. ${\hat{\hat{p}}}_{D}=\hat{x}_{D}/\hat{n}_{D}$, $\hat{\hat{\mathrm{LL}}}$ and $\hat{\hat{\mathrm{UL}}}$ are the estimated lower and upper 95% confidence interval obtained by each method, using the corresponding estimated values of $\hat{x}_{D}$ and $\hat{n}_{D}$. Relative error (%) with sign associated with each method is defined as the difference between the meta-analysis value minus the corresponding estimated one divided by the meta-analysis value.

|  | **Wald** | **Arcsine** | **Agresti-Coull** | **Clopper-Pearson** |
| --- | --- | --- | --- | --- |
| $\hat{p}_{D}$ | 1.841×10^-3^ | | | |
| $\hat{x}_{D}$ | 97 | 97 | 97 | 101 |
| $\hat{n}_{D}$ | 52,554 | 52,328 | 52,660 | 55,146 |
| ${\hat{\hat{p}}}_{D}$ | 1.846×10^-3^ | 1.854×10^-3^ | 1.842×10^-3^ | 1.832×10^-3^ |
| ${\hat{\hat{p}}}_{D}$ relative error (%) | −0.26 | −0.69 | −0.06 | 0.52 |
| $\hat{\mathrm{LL}}$ | 1.492×10^-3^ | | | |
| $\hat{\hat{\mathrm{LL}}}$ | 1.479×10^-3^ | 1.503×10^-3^ | 1.492×10^-3^ | 1.492×10^-3^ |
| $\hat{\hat{\mathrm{LL}}}$ relative error (%) | 0.89 | −0.75 | 0.00 | -0.002 |
| $\hat{\mathrm{UL}}$ | 2.225×10^-3^ | | | |
| $\hat{\hat{\mathrm{UL}}}$ | 2.213×10^-3^ | 2.241×10^-3^ | 2.228×10^-3^ | 2.225×10^-3^ |
| $\hat{\hat{\mathrm{UL}}}$ relative error (%) | 0.55 | −0.72 | −0.14 | 0.00 |

Table S4 Genes analysed in this study and the allele number sequenced in each disease cohort.

Allele number is twice the cases included in the study. *, TNNI3 exons 7 and 8 only; °, truncating variants in TTN only; -, not measured.

|  | **LMM** | **LMM2** | **OMGL** | **BRGL** | **GDx** | **RBHT** | **SLD** | **NHCS** | **AHCE** | **max AN** |
| --- | --- | --- | --- | --- | --- | --- | --- | --- | --- | --- |
| **HCM** |  |  |  |  |  |  |  |  |  |  |
| *ACTC1* | 5300 | - | 3070 | - | 4740 | 836 | 510 | 182 | 914 | **15342** |
| *MYBPC3* | 5824 | - | 6534 | 1260 | 4740 | 836 | 510 | 182 | 914 | **20590** |
| *MYH7* | 5824 | - | 6400 | 1260 | 4740 | 836 | 510 | 182 | 914 | **20456** |
| *MYL2* | 5300 | - | 3070 | - | 4740 | 836 | 510 | 182 | 914 | **15342** |
| *MYL3* | 5300 | - | 3070 | - | 4740 | 836 | 510 | 182 | 914 | **15342** |
| *TNNI3* | 5824 | - | 6270 | 1260* | 4740 | 836 | 510 | 182 | 914 | **20326** |
| *TNNT2* | 5824 | - | 6382 | 1260 | 4740 | 836 | 510 | 182 | 914 | **20438** |
| *TPM1* | 5824 | - | 3070 | - | 4740 | 836 | 510 | 182 | 914 | **15866** |
| **max AN** | **5824** | - | **6534** | **1260** | **4740** | **836** | **510** | **182** | **914** | **20800** |
| **DCM** |  |  |  |  |  |  |  |  |  |  |
| *BAG3* | - | 366 | - | - | - | 1758 | - | 214 | 160 | **2498** |
| *DES* | 1180 | 366 | 608 | - | - | 1758 | - | 214 | 160 | **4286** |
| *TTN* | 312**°** | 366 | 608**°** | - | - | 1758 | - | 214 | 160 | **3418** |
| *MYH7* | 1512 | 366 | 1118 | - | - | 1758 | - | 214 | 160 | **5128** |
| *TNNC1* | 312 | 366 | - | - | - | 1758 | - | 214 | 160 | **2810** |
| *TNNT2* | 1512 | 366 | 996 | - | - | 1758 | - | 214 | 160 | **5006** |
| *LMNA* | 1480 | 366 | 608 | - | - | 1758 | - | 214 | 160 | **4586** |
| *FLNC* | - | - | - | - | - | - | - | - | - | - |
| *PLN* | 1480 | 366 | 710 | - | - | 1758 | - | 214 | 160 | **4688** |
| *SCN5A* | - | 366 | 608 | - | - | 1758 | - | 214 | 160 | **3106** |
| *RBM20* | 312 | 366 | - | - | - | 1758 | - | 214 | 160 | **2810** |
| *DSP* | 246 | 366 | 608 | - | - | 1758 | - | 214 | 160 | **3352** |
| **Max AN** | **1512** | **366** | **1118** | - | - | **1758** | - | **214** | **160** | **5128** |

Table S5 Ancestry, age, and sex; case cohort participant summary information.

See excel file.

Table S6 Variant counts in the aggregated dataset per gene and variant consequence for HCM.

See excel file.

Table S7 Variant counts in the aggregated dataset per gene and variant consequence for DCM.

See excel file.

Table S8 Summary information of 1,340 rare variants in HCM-associated genes.

See excel file.

Table S9 Summary information of 665 rare variants in DCM-associated genes.

See excel file.

Table S10 Penetrance estimates for 257 rare variants in HCM-associated genes.

See excel file.

Table S11 Penetrance estimates for 59 rare variants in DCM-associated genes.

See excel file.

**Table S12** Estimated penetrance of eleven variants more common in non-EUR ancestry.

See excel file.

**Table S13** Variant counts in the aggregated UKBB dataset per gene and variant consequence for HCM.

See excel file.

**Table S14** Variant counts in the aggregated UKBB dataset per gene and variant consequence for DCM.

See excel file.

**Table S15** Aggregated penetrance by curation.

See excel file.

**Table S16** Aggregated penetrance by rarity.

See excel file.

**Table S17** Aggregated penetrance by age.

See excel file.

**Table S18** Aggregated penetrance by gene for HCM.

See excel file.

**Table S19** Aggregated penetrance by gene for DCM.

See excel file.

**Table S20** Aggregated penetrance by sex.

See excel file.

### 4 Supplementary lists

List S1 Selection of the Agresti-Coull method and other methods assessed to estimate the number of cases and the population size for the disease prevalence.

For each method considered, the estimated values of $x_{D}$ and $n_{D}$ are derived as shown below. The best method was selected by assessment of the Euclidean distance between $(\hat{p}_{D},LL(\hat{p}_{D}),UL(\hat{p}_{D})$), the estimated value, and the lower and upper limits of the 95% confidence interval of the prevalence obtained from the meta-analysis, and $({\hat{\hat{p}}}_{D},LL({\hat{\hat{p}}}_{D}),UL({\hat{\hat{p}}}_{D})$) obtained by each method, using the corresponding estimated values of $\hat{x}_{D}$ and $\hat{n}_{D}$. For simplicity of notation, we omit the subscript $D$ and set $\hat{\mathrm{LL}}=\mathrm{LL}\left( \hat{p} \right)$, $\hat{\mathrm{UL}}=UL(\hat{p})$, $\hat{\hat{p}}=\hat{x}/\hat{n}$, $\hat{\hat{\mathrm{LL}}}=LL(\hat{\hat{p}})$ and $\hat{\hat{\mathrm{UL}}}=UL(\hat{\hat{p}})$.

A) *Wald method*

$$\hat{\sigma}=\left( \frac{\hat{\mathrm{UL}}-\hat{\mathrm{LL}}}{2z_{1-\alpha/2}} \right)^{2},$$

$$\hat{n}=\left\lceil\frac{\hat{p}\hat{q}}{\hat{\sigma}} \right\rceil,$$

$$\hat{x}=\left\lceil\hat{n}\hat{p} \right\rceil,$$

(**Eq. S4**)

where $\hat{\sigma}$ is the estimated standard error of $\hat{p}$ obtained from the meta-analysis, $z_{1-\alpha/2}$ is the $1-\alpha/2$ quantile of the standard normal distribution, $\hat{q}=1-\hat{p}$ and $\left\lceil y \right\rceil$ indicates the smallest integers not less than $y$.

B) *Arcsine method*

$$\hat{n}=\left\lceil\left[ \frac{z_{1-\alpha/2}}{2\left( \sin^{-1} \sqrt{\hat{\mathrm{LL}}}-\sin^{-1} \sqrt{\hat{p}} \right)} \right]^{2} \right\rceil,$$

$$\hat{x}=\left\lceil\hat{n}\hat{p} \right\rceil.$$

(**Eq. S5**)

C) *Agresti-Coull method*

$$\hat{n}=\left\lceil\frac{\tilde{p}\tilde{q}}{\left( \frac{\tilde{p}-\hat{\mathrm{LL}}}{z_{1-\alpha/2}^{2}} \right)^{2}}-z_{1-\alpha/2}^{2} \right\rceil,$$

$$\hat{x}=\left\lceil\left( \hat{n}+z_{1-\alpha/2}^{2} \right)\tilde{p}-\frac{z_{1-\alpha/2}^{2}}{2} \right\rceil,$$

(**Eq. S6**)

where $\tilde{p}$ is defined (**Eq. S2**) and $\tilde{q}=1-\tilde{p}$. However, the quantity $\tilde{p}$ is not available from the meta-analysis and depends on unknown value $n$. For this reason, the solution (**Eq. S6**) is obtained numerically. Using the fact that $\tilde{p}>\hat{p}$ if $\hat{p}<0.5$, the solution is attained by finding the positive constant $c_{0}$ such that $\tilde{p}=\hat{p}+c_{0}$ for which $\hat{x}/\hat{n}=\hat{p}$ under the constraint that $\hat{p}<0.5$, *i.e.*, $\hat{x}<\left\lceil\hat{n}/2 \right\rceil$.

D) *Clopper-Pearson method*

$$LL=F_{\mathrm{Beta}}^{-1}\left( \alpha/2;x,n-x+1 \right),$$

$$UL=F_{\mathrm{Beta}}^{-1}\left( 1-\alpha/2;x+1,n-x \right),$$

(**Eq. S7**)

where $\mathrm{LL}$ and $\mathrm{UL}$ are the theoretical exact lower and upper limits of the 95% confidence interval and $F_{\mathrm{Beta}}^{-1}(\cdot)$ is the inverse of the cumulative density function of the Beta density. The solution for $x$ and $n$, with $x\leq n$, is obtained numerically as the values that minimise the Euclidean distance between ($LL,UL$) defined (**Eq. S7**) and $(\hat{\mathrm{LL}},\hat{\mathrm{UL}}$), the lower and upper limits of the $1-\alpha$ confidence interval of the prevalence obtained from the meta-analysis, respectively. To reduce the computational cost of the exhaustive search, we also assume $\hat{x}<\left\lceil\hat{n}/2 \right\rceil$ as in the Agresti-Coull method.

List S2 Methods assessed to derive the confidence intervals of penetrance.

We consider two groups of methods to assess the confidence interval of the penetrance. In the first group (G1), the confidence interval for the ratio of the random variables $A|D$ and $A$ (**Eq. S1**) is obtained similarly to the derivation of the confidence interval for relative risk^5^. Most of the methods are readily available in the R package *ratesci*^71^ unless stated otherwise. The upper and lower limit of the $\left( 1-\alpha\right)\%$ confidence interval is then multiplied by the estimated value of the prevalence $p_{D}$.

The second group (G2) of methods consider the prevalence $D$ as a random variable and the confidence interval is derived assuming the independence of all the quantities involved. The second group of methods rely on the Delta method applied on the log-transformed random variable $D|A$ or directly on $D|A$ (**Eq. S1**) with/without improved mean approximation.

To address the problem of degeneracy, *i.e.*, the confidence interval’s width is 0 when the probability of success is either 0 or 1, we added the constant $d_{x}=0.5$ and $d_{n}=0.5$ to $x$ and $n$, respectively, in the binomial random variables $A|D$, $A$ and $D$^41,42^ or add a continuity correction^3^ to the confidence interval. To avoid “overshooting”^2^, *i.e.*, the confidence interval of penetrance could be outside the interval $[0,1]$ and the results are truncated in the interval $[0,1]$. The $\mathrm{CI}\left( p_{D|A} \right)$ is obtained as follows:

A) $T$he product of $p_{D}$ and the $\left( 1-\alpha\right)\%$ confidence interval of the ratio of binomial proportions using the with Delta method on $\log(A|D / A)=\log\left( A | D \right)-\log(A)$^41,42,72^ without degeneracy

$$\mathrm{CI}\left( p_{D|A} \right)=p_{D}exp\left\{ \log\frac{p_{A|D}^{d}}{p_{A}^{d}}\pm z_{1-\alpha/2}\sqrt{\frac{1-p_{A|D}^{d}}{p_{A|D}^{d}n_{A|D}^{d}}+\frac{1-p_{A}^{d}}{p_{A}^{d}n_{A}^{d}}} \right\},$$

(**Eq. S8**)

where $n_{A}^{d}=(n_{A}+d_{n})$, $p_{A}^{d}=(x_{A}+d_{x})/(n_{A}+d_{n})$ and similarly for $n_{A|D}^{d}$ and $p_{A|D}^{d}$.

B) $T$he product of $p_{D}$ and the $\left( 1-\alpha\right)\%$ confidence interval of the ratio of binomial proportions using the “method of variance estimates recovery” (MOVER)^2,73,74^ method with continuity correction implemented in the function moverci included in the R package *ratesci*.
C) $T$he product of $p_{D}$ and the $\left( 1-\alpha\right)\%$ confidence interval of the ratio of binomial proportions using the approximate Bayesian “method of variance estimates recovery” (MOVER-B)^2,73–75^ with beta priors and continuity correction implemented in the function moverbci included in the R package *ratesci*.
D) The product of $p_{D}$ and the $\left( 1-\alpha\right)\%$ confidence interval of the ratio of binomial proportions with “skewness-corrected asymptotic score” (SCAS)^43,76,77^ with continuity correction implemented in the function scasci included in the R package *ratesci*.
G2.A) $\left( 1-\alpha\right)\%$ confidence interval of the product and ratio of binomial proportions using the with Delta method on $\log\left( D | A \right)=\log(D\times A|D/A)=\log\left( D \right)+\log\left( A | D \right)-\log(A)$ without degeneracy

$$\mathrm{CI}\left( p_{D|A} \right)=\exp\left\{ \log\frac{{p_{D}^{d}p}_{A|D}^{d}}{p_{A}^{d}}\pm z_{1-\alpha/2}\sqrt{\frac{1-p_{D}^{d}}{p_{D}^{d}n_{D}^{d}}+\frac{1-p_{A|D}^{d}}{p_{A|D}^{d}n_{A|D}^{d}}+\frac{1-p_{A}^{d}}{p_{A}^{d}n_{A}^{d}}} \right\}.$$

(**Eq. S9**)

G2.B) $\left( 1-\alpha\right)\%$ confidence interval of the product and ratio of binomial proportions using the with Delta method on $\log\left( D | A \right)=\log(D\times A|D/A)=\log\left( D \right)+\log\left( A | D \right)-\log(A)$ with improved mean approximation and without degeneracy

$$\mathrm{CI}\left( p_{D|A} \right)=\exp\left\{ \log\frac{{p_{D}^{d}p}_{A|D}^{d}}{p_{A}^{d}}-\frac{1}{2}\left( \frac{1-p_{D}^{d}}{p_{D}^{d}n_{D}^{d}}+\frac{1-p_{A|D}^{d}}{p_{A|D}^{d}n_{A|D}^{d}}-\frac{1-p_{A}^{d}}{p_{A}^{d}n_{A}^{d}} \right)\pm z_{1-\alpha/2}\sqrt{\frac{1-p_{D}^{d}}{p_{D}^{d}n_{D}^{d}}+\frac{1-p_{A|D}^{d}}{p_{A|D}^{d}n_{A|D}^{d}}+\frac{1-p_{A}^{d}}{p_{A}^{d}n_{A}^{d}}} \right\}.$$

(**Eq. S10**)

G2.C) $\left( 1-\alpha\right)\%$ confidence interval of the product and ratio of binomial proportions using the with Delta method on $D\left| A=D\times A \right|D/A$ without degeneracy

$$\mathrm{CI}\left( p_{D|A} \right)=\frac{{p_{D}^{d}p}_{A|D}^{d}}{p_{A}^{d}}\left( 1\pm z_{1-\alpha/2}\sqrt{\frac{1-p_{D}^{d}}{p_{D}^{d}n_{D}^{d}}+\frac{1-p_{A|D}^{d}}{p_{A|D}^{d}n_{A|D}^{d}}+\frac{1-p_{A}^{d}}{p_{A}^{d}n_{A}^{d}}} \right).$$

(**Eq. S11**)

G2.D) $\left( 1-\alpha\right)\%$ confidence interval of the product and ratio of binomial proportions using the Delta method on $D\left| A=D\times A \right|D/A$ with improved mean approximation and without degeneracy

$$\mathrm{CI}\left( p_{D|A} \right)=\frac{{p_{D}^{d}p}_{A|D}^{d}}{p_{A}^{d}}\left( 1+\frac{1-p_{A}^{d}}{p_{A}^{d}n_{A}^{d}}\pm z_{1-\alpha/2}\sqrt{\frac{1-p_{D}^{d}}{p_{D}^{d}n_{D}^{d}}+\frac{1-p_{A|D}^{d}}{p_{A|D}^{d}n_{A|D}^{d}}+\frac{1-p_{A}^{d}}{p_{A}^{d}n_{A}^{d}}} \right).$$

(**Eq. S12**)

### 5 Supplementary figures

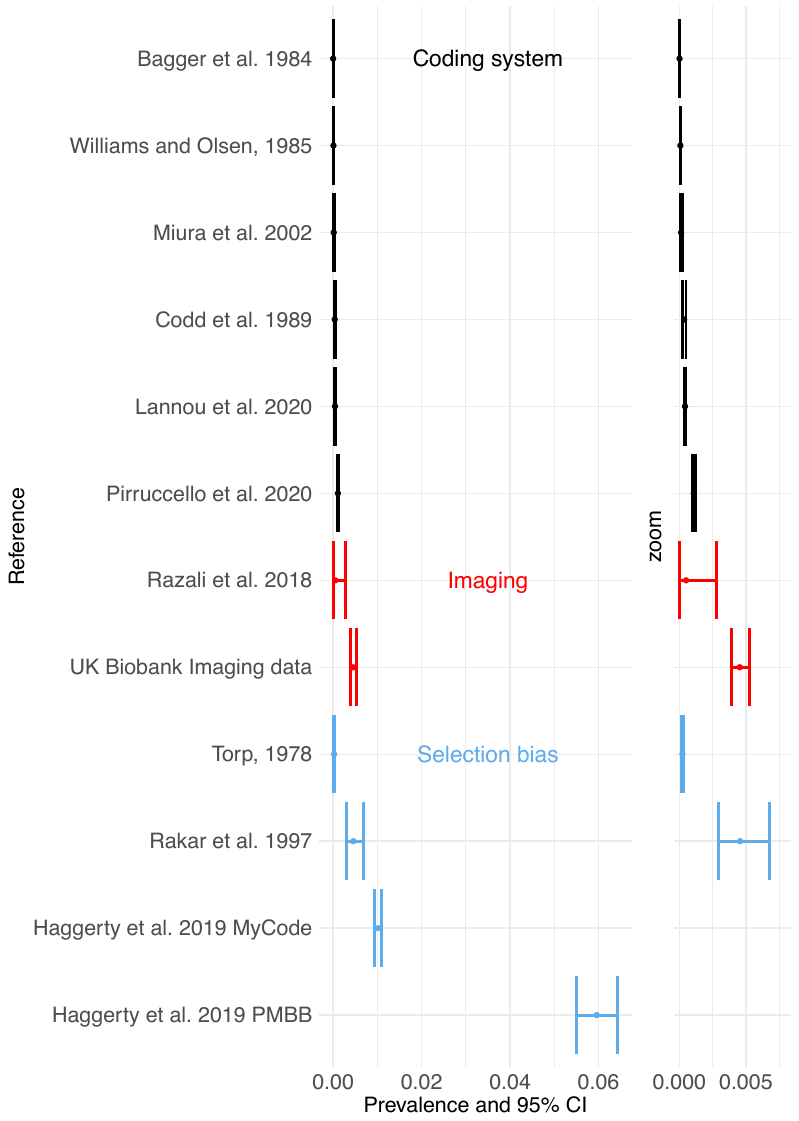

Figure S1 Meta-analysis of population prevalence estimated for DCM in literature.

(Left panel) Forest plot depicting the prevalence and associated binomial confidence interval for each literature reference. (Right panel, zoom) The same forest plot with the x-axis shortened to between 0 and 0.008. Coding system, prevalence estimates that were derived using large population datasets with International Classification of Diseases (ICD) or other coding systems and have decreased prevalence estimates; Imaging, prevalence estimates that were derived using imaging data such as cardiac MRI or echocardiography and provide estimates that better reflect the true DCM prevalence; Selection bias, patients referred for imaging measures based on previous symptoms and have increased prevalence, or, participants are active, selected for being young or athletic and have decreased prevalence, or, participants are elderly and the prevalence estimate is substantially increased.

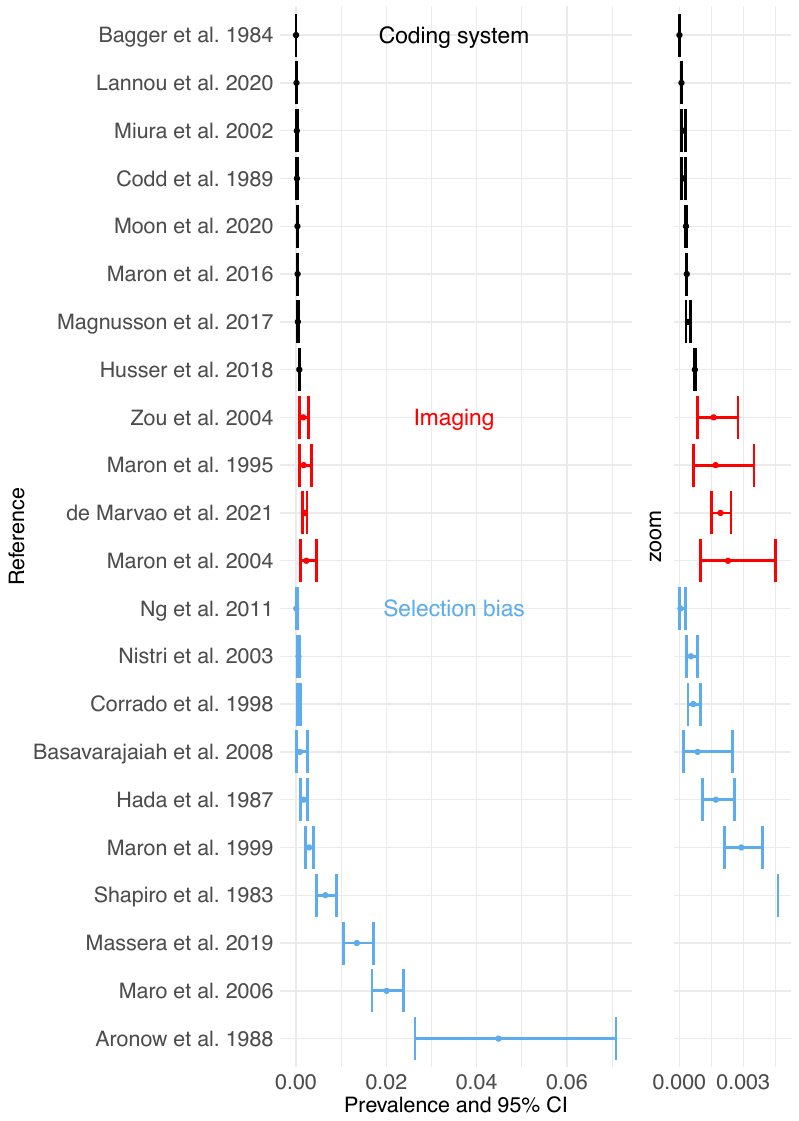

Figure S2 Meta-analysis of population prevalence estimated for HCM in literature.

(Left panel) Forest plot depicting the prevalence and associated binomial confidence interval for each literature reference. (Right panel, zoom) The same forest plot with the x-axis shortened to between 0 and 0.005. Coding system, prevalence estimates that were derived using large population datasets with International Classification of Diseases (ICD) or other coding systems and have decreased prevalence estimates; Imaging, prevalence estimates that were derived using imaging data such as cardiac MRI or echocardiography and provide estimates that better reflect the true HCM prevalence; Selection bias, patients referred for imaging measures based on previous symptoms and have increased prevalence, or, participants are active, selected for being young or athletic and have decreased prevalence, or, participants are elderly and the prevalence estimate is substantially increased. References^12,13,28–37,14,78,79,15,20,23–27^.

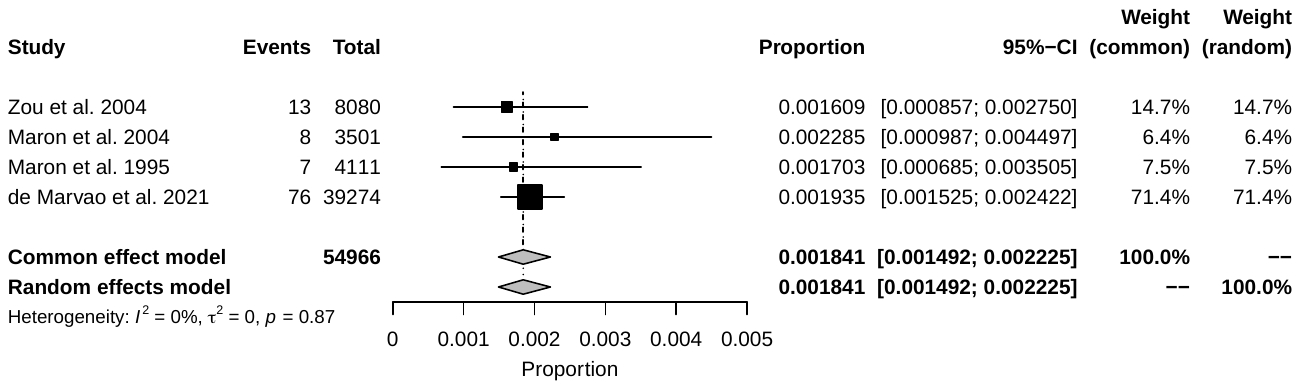

**All**

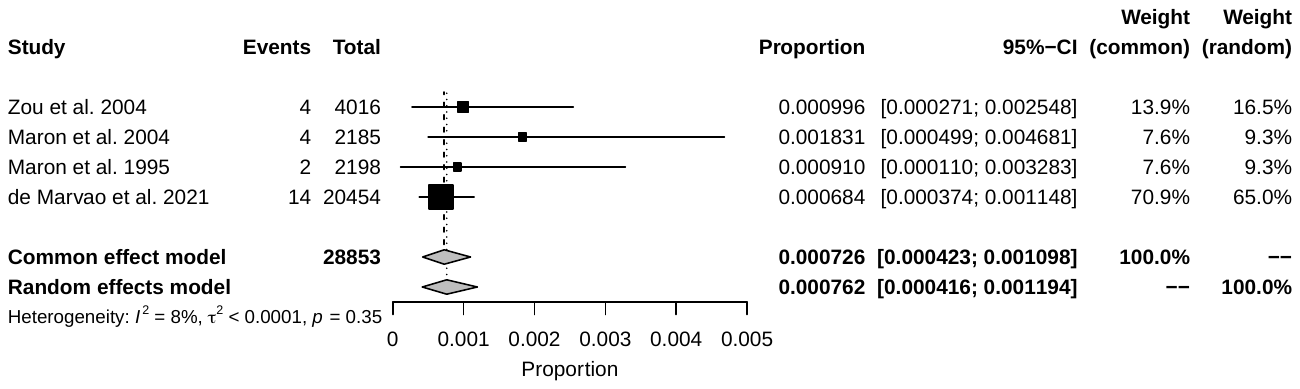

**Women**

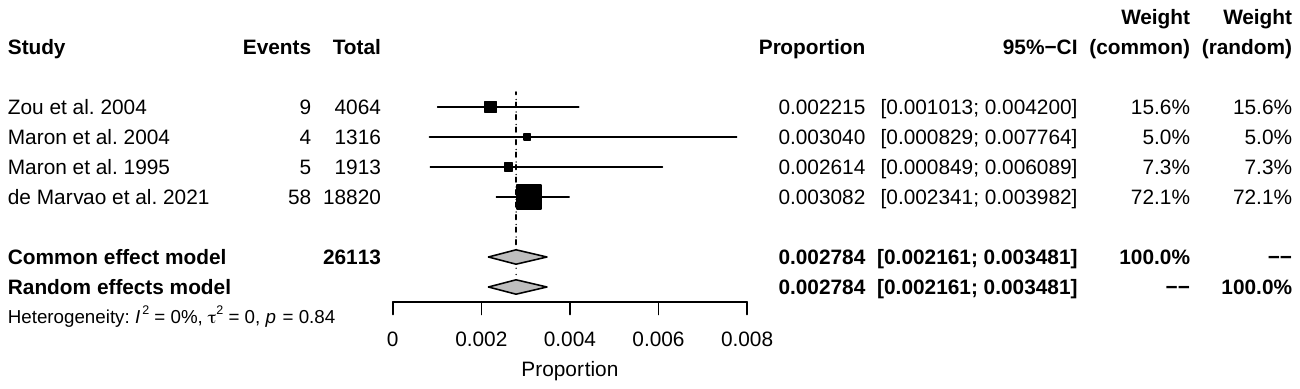

**Men**

Figure S3 Meta-analysis for binomial proportions of four population prevalence estimates of hypertrophic cardiomyopathy.

Four studies were included that had assessed for the prevalence of HCM using imaging for population screening. The heterogeneity indexes are not significant (P>0.05).

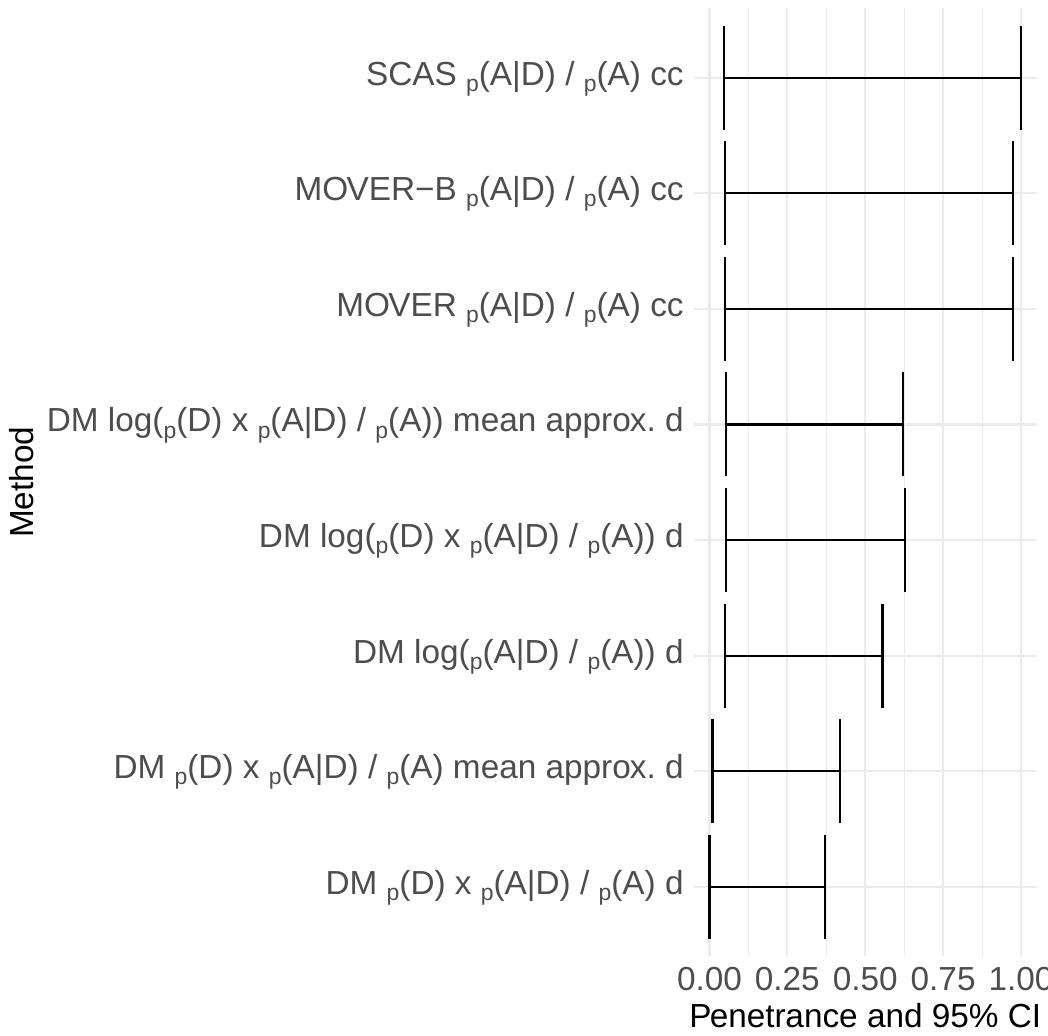

Figure S4 Assessment of nine methods to estimate the 95% confidence interval of penetrance.

The method of choice is DM on $p(D)\times p\left( A | D \right)/ p(A)$ mean approx. d. SCAS, skewness-corrected asymptotic score; DM, Delta method; mean approx., improved mean approximation; cc, continuity correction; d, adjustment for degeneracy; $p(D)$, probability of disease; $p(A|D)$, probability of disease given the allele; $p(A)$, probability of the allele in the population.

A)	B)

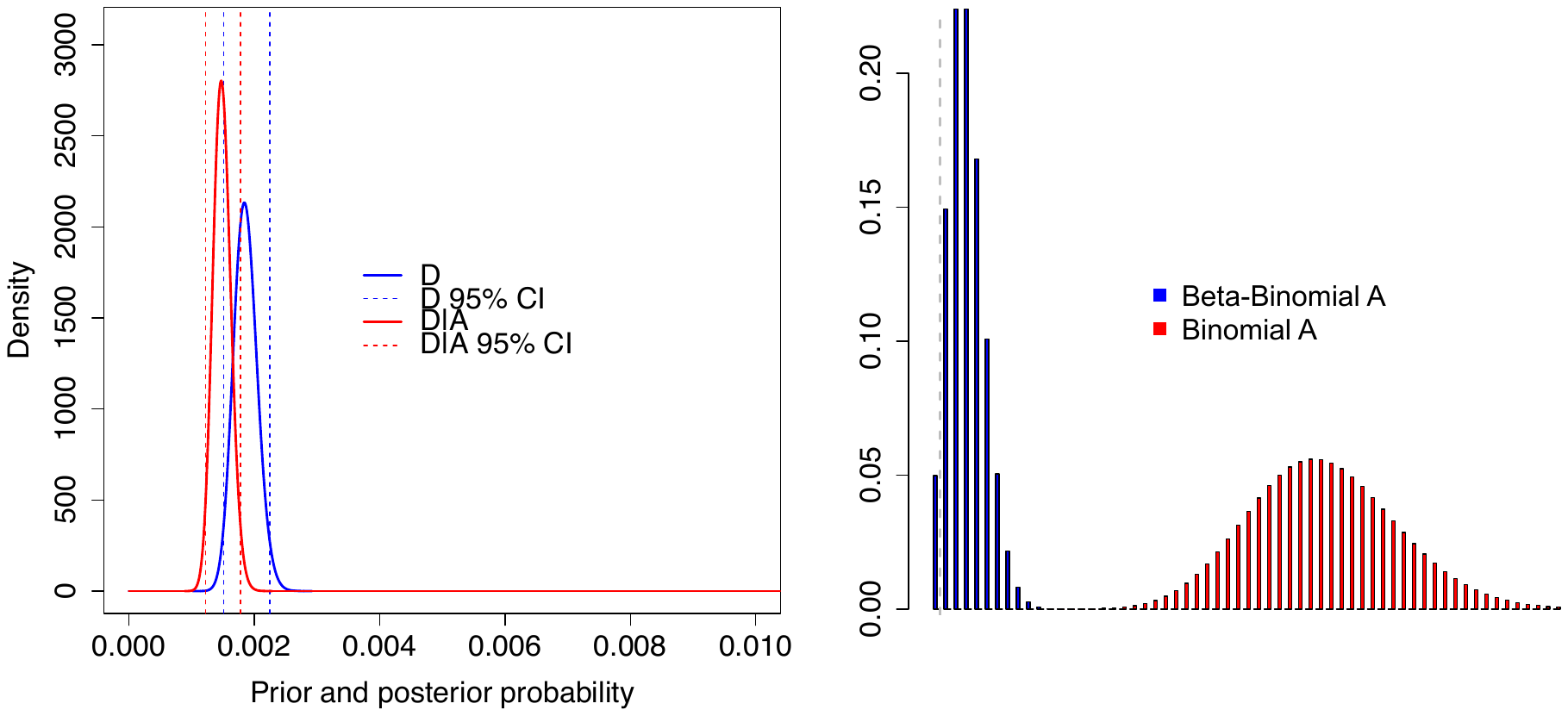

Figure S5 A fully Bayesian approach is not suitable for estimating penetrance in this manner.

A) Based on real data parameter specifications, the Beta distribution of the prevalence $D$ and the posterior Beta distribution of the penetrance $D|A$ have marginal overlap. B) The divergence between the known distribution of $A$ (Beta-Binomial) once $D$ and $A|D$ are specified (beta and Binomial densities, respectively) and the Binomial distribution of $A$ (independent from $D$ and $A|D$) are very different.

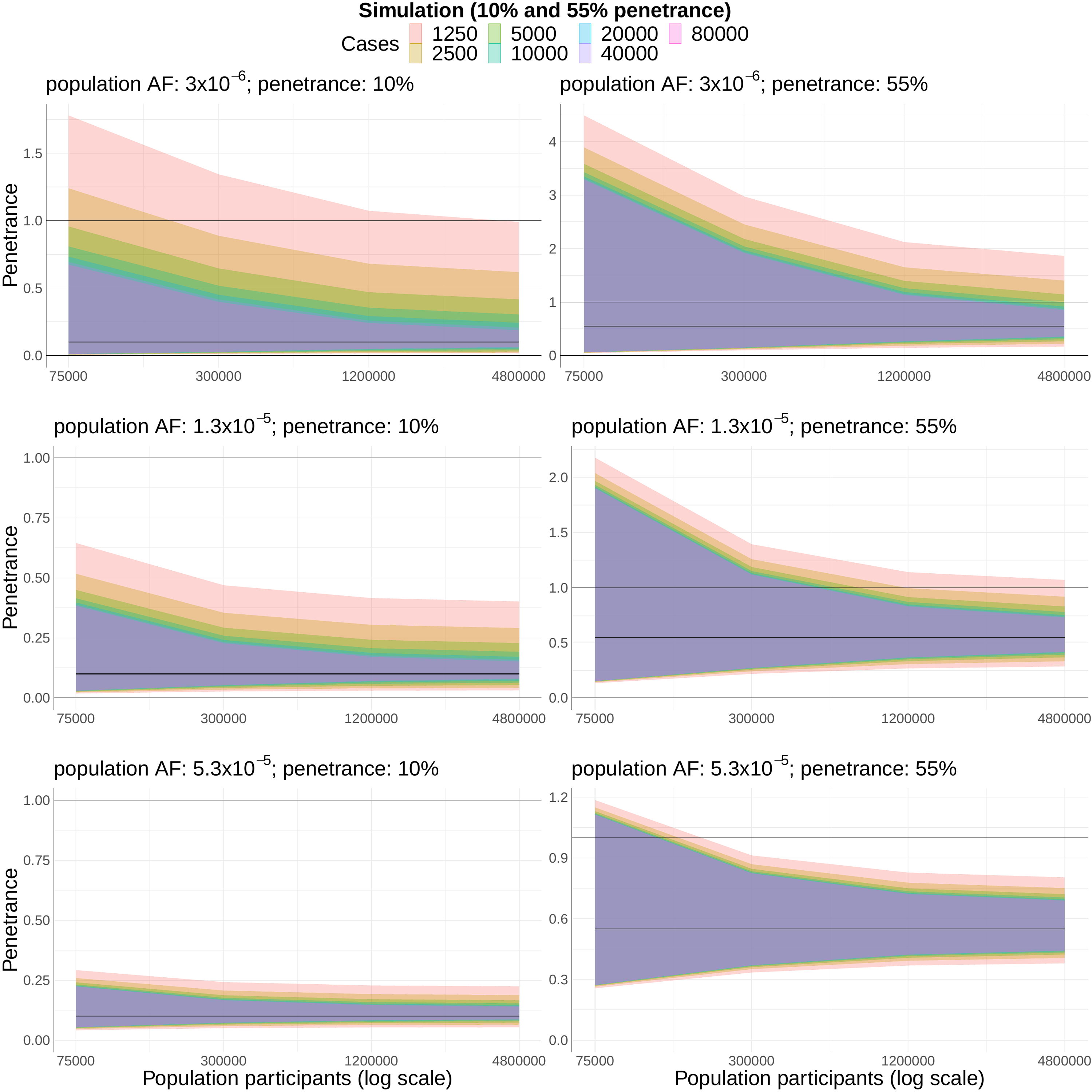

Figure S6 With 10,000 cases, increasing population participants aids penetrance estimates.

Efforts to increase reference population sample size will provide additional confidence (i.e., narrower confidence intervals) than further case aggregation after 10,000 cases is reached (with the caveat that more variants will be identified). The graph denotes the results of a simulation of a variant with 10% estimated penetrance and 55% estimated penetrance. The x-axis varies population reference cohort size, and the legend varies case cohort size. Black line, 100% penetrance; pink line, penetrance estimate.

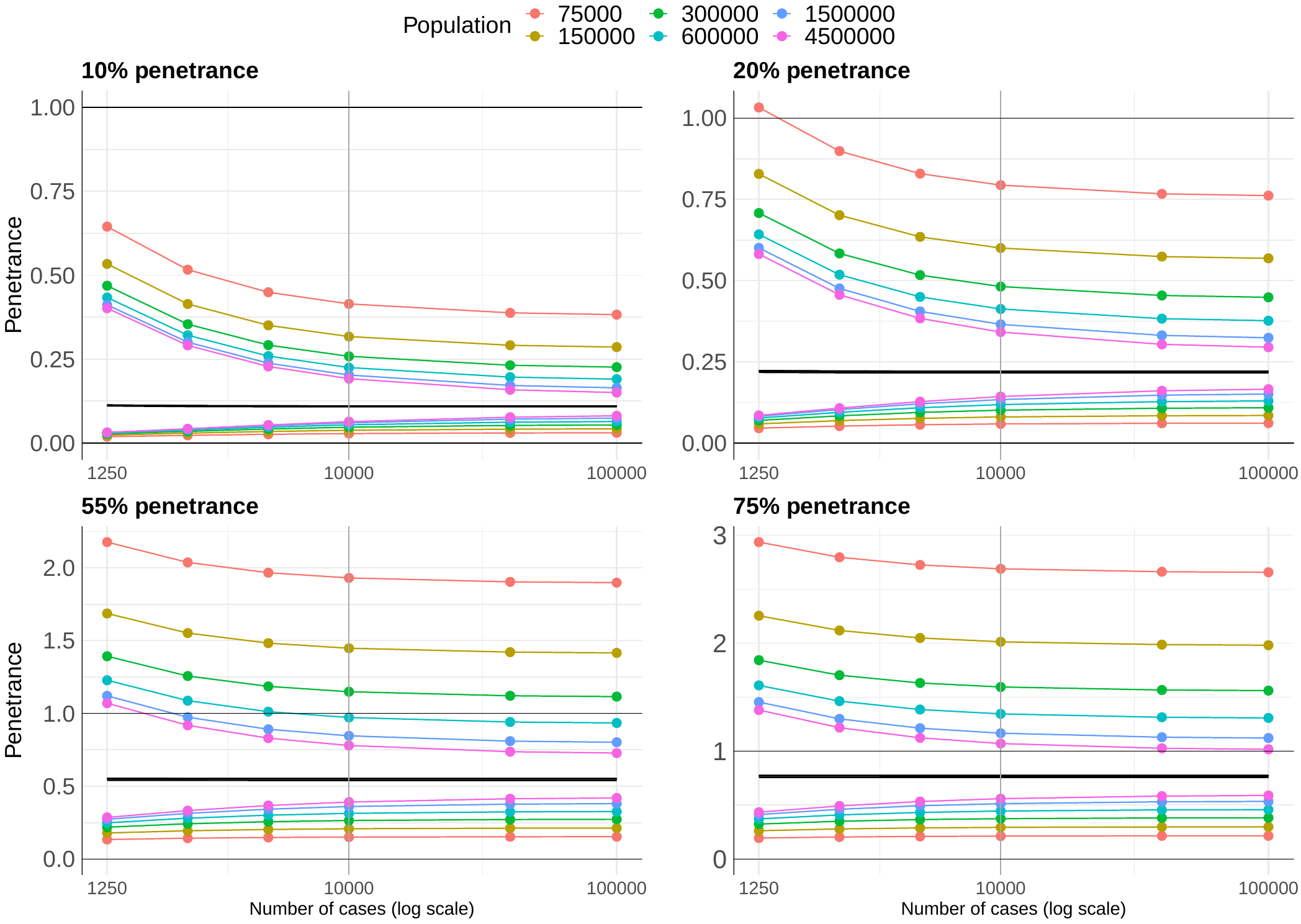

Figure S7 Negligible gains in confidence will be provided by increasing case sample size, while substantial gains will be observed by incorporation of future large-scale population datasets.

Example variants had a penetrance of ~10%, ~20%, ~55%, and ~75% (popAF=0.000013, caseAF=0.0008, 0.0016, 0.004, 0.0056, respectively). The penetrance estimate is shown as a black line, the UCI are coloured above the penetrance estimate and the LCI coloured below. The grey horizontal line denotes depicts a penetrance of 1.0 or 100% for assessment of the UCI. The grey vertical line denotes the sample size used in this study. The sizes of population reference cohorts are depicted as coloured points. The x-axis describes case cohort samples, and the legend describes the number of gnomAD and UKB participants.

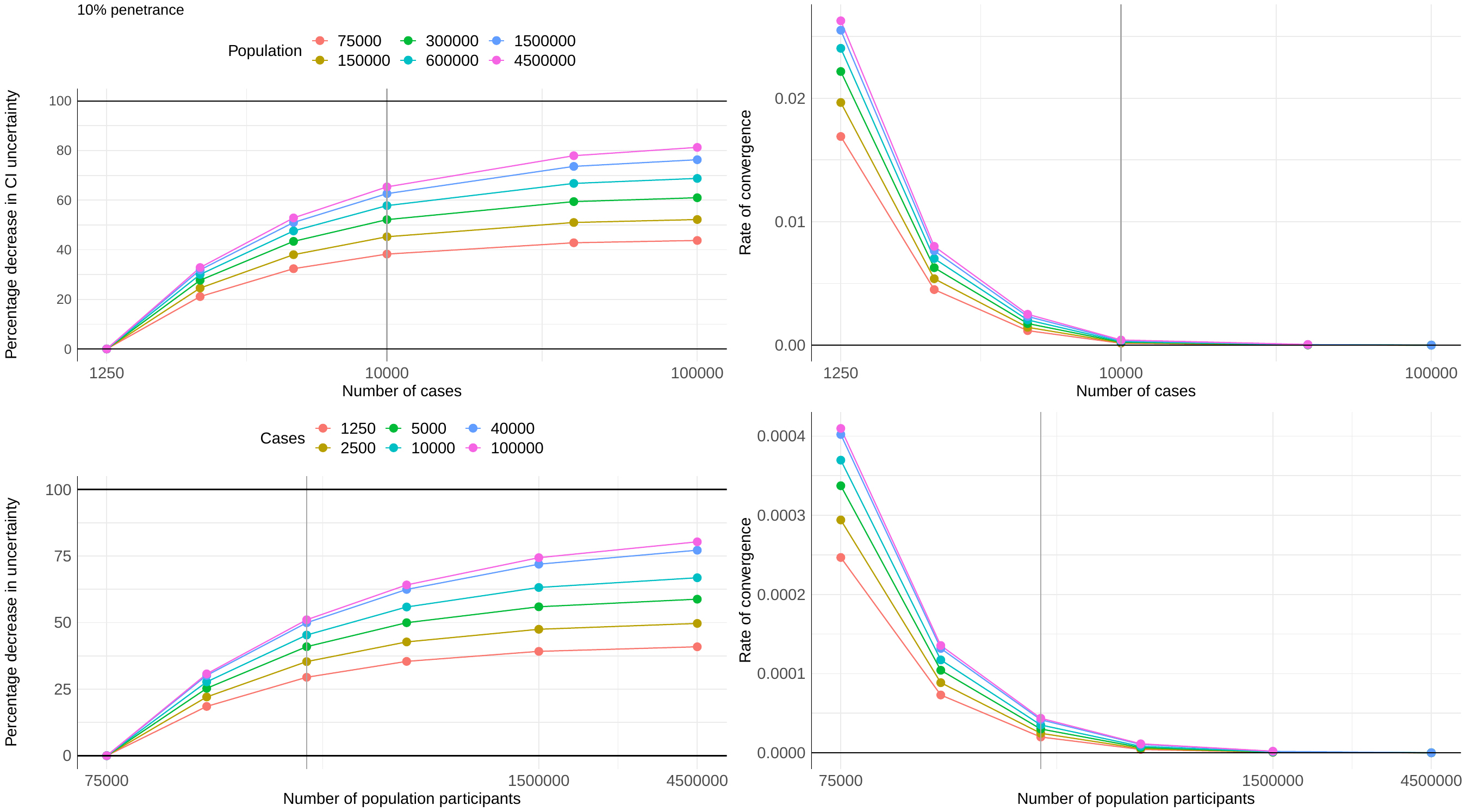

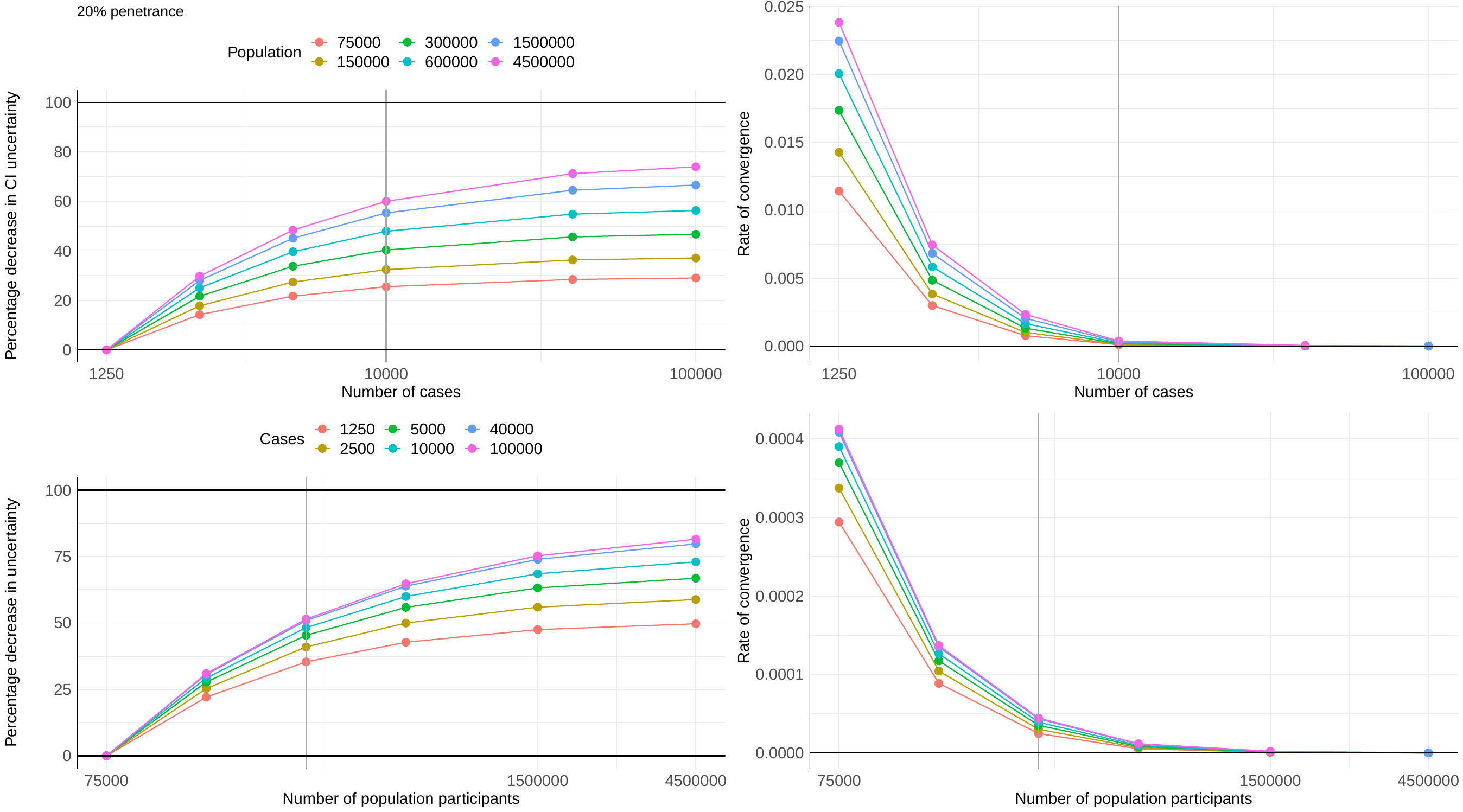

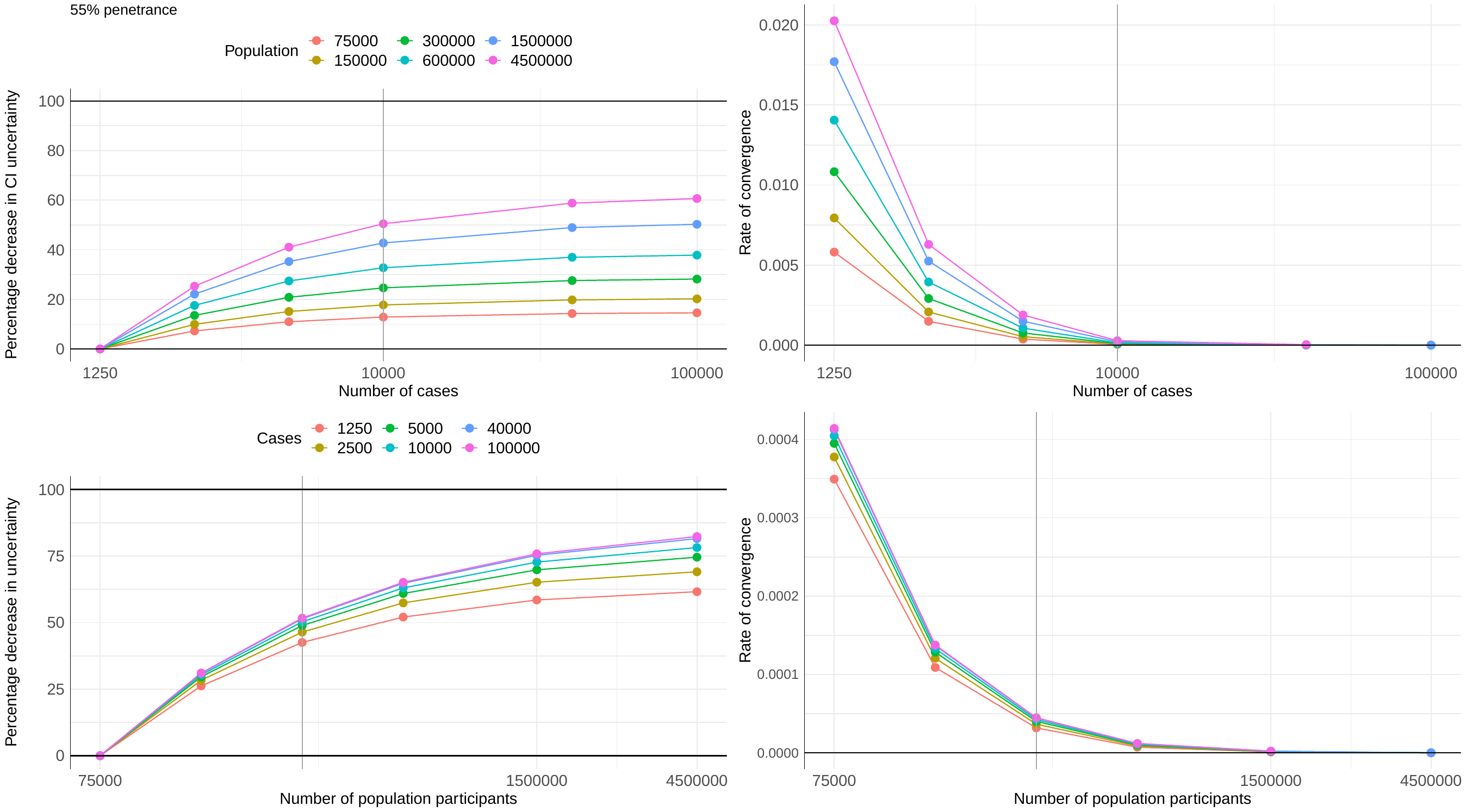

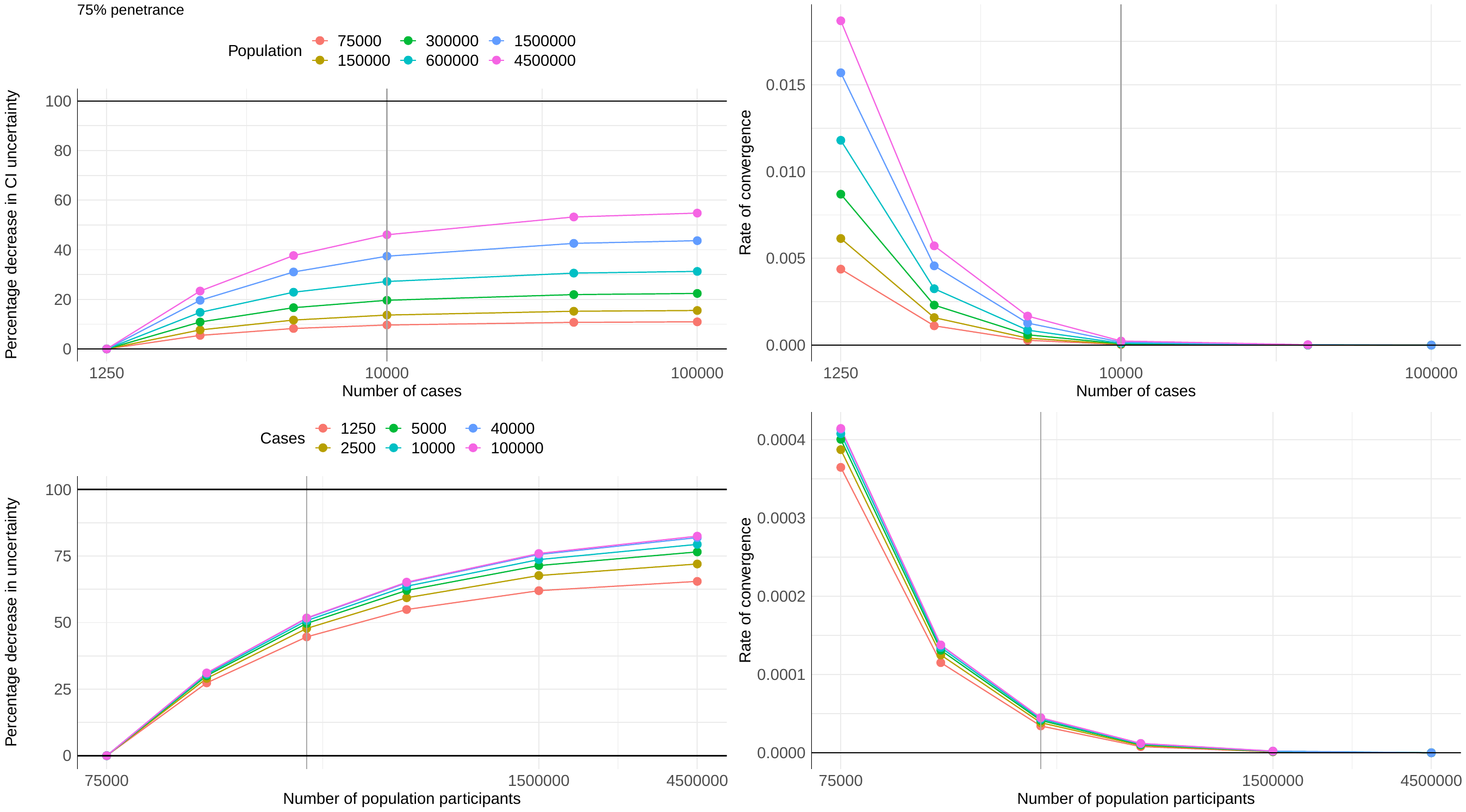

Figure S8 Simulation of the gain in confidence of the penetrance estimate with increasing sample size.

Example variants had a penetrance of ~10%, ~20%, ~55%, and ~75% (popAF=0.000013, caseAF=0.0008, 0.0016, 0.004, 0.0056, respectively). Estimates of the percentage decrease in uncertainty (or gain in certainty/error) with increasing sample size are shown on the left. Estimates of the rate of convergence of the error are shown on the right. The grey vertical line denotes the sample size used in this study. The size of population or case cohort are depicted as coloured points and indicated by the legend. The x-axis describes cohort size.

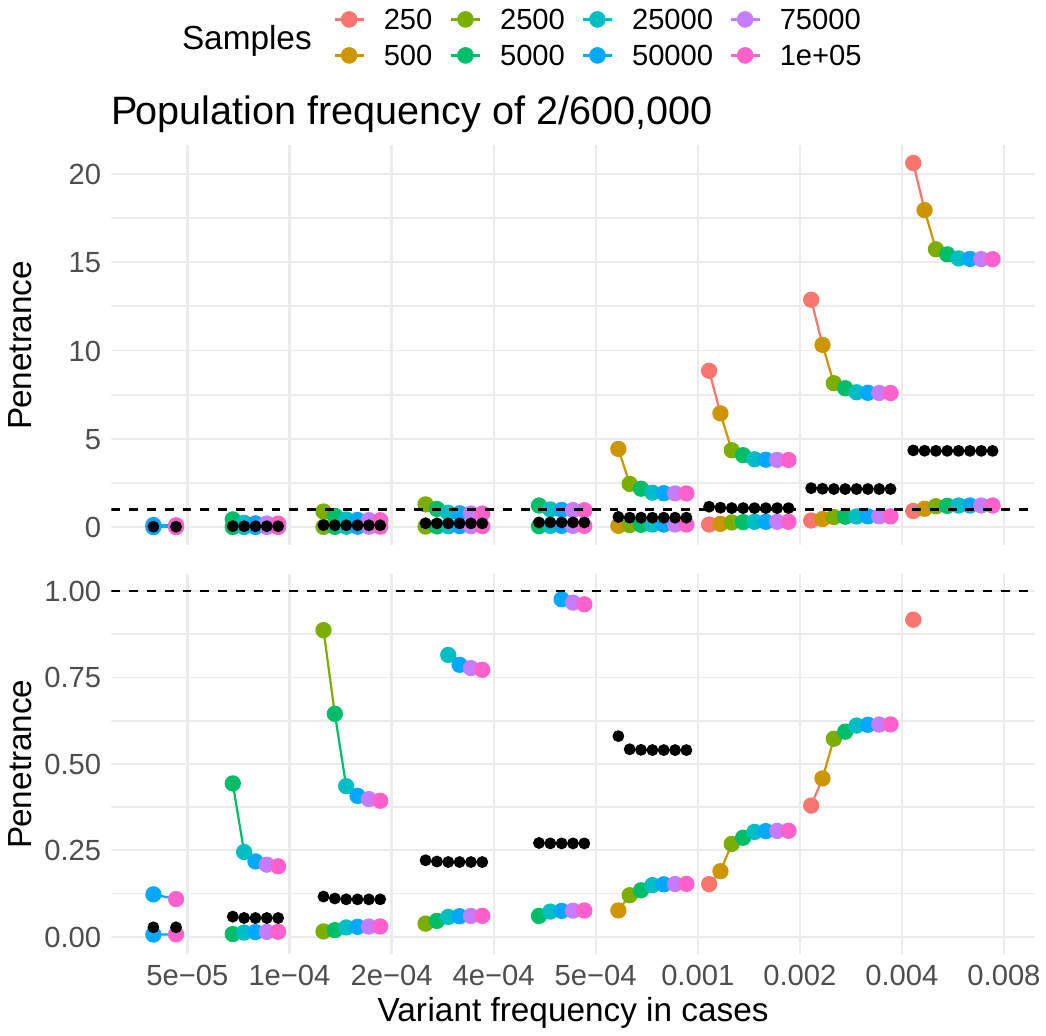

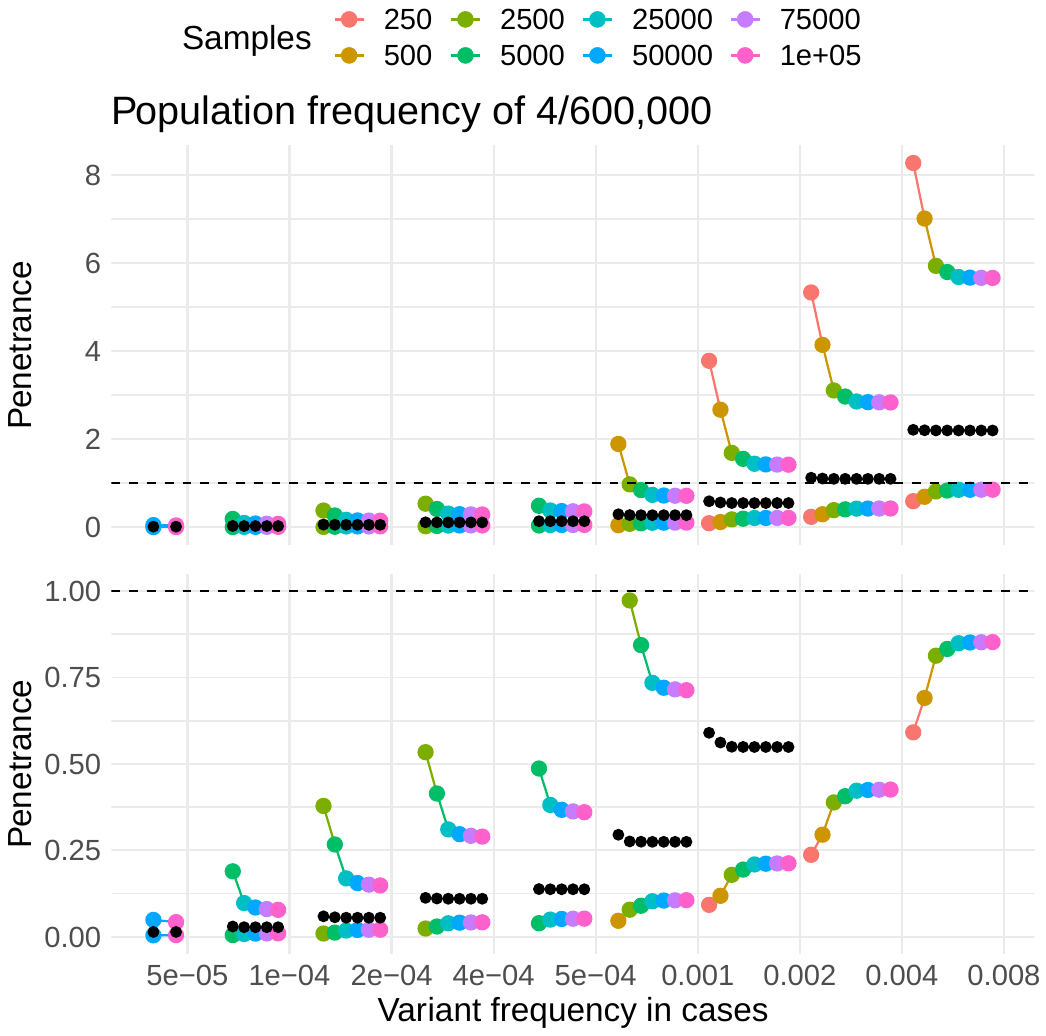

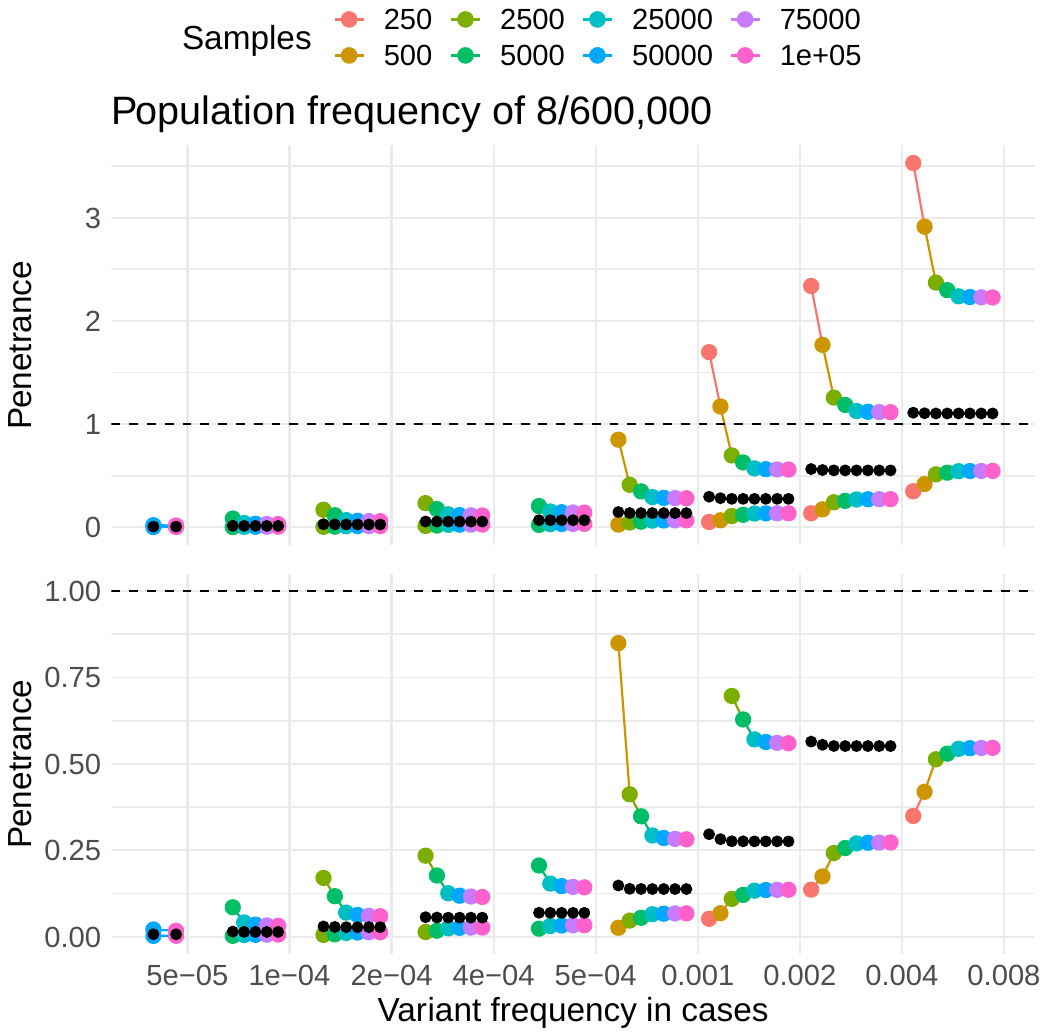

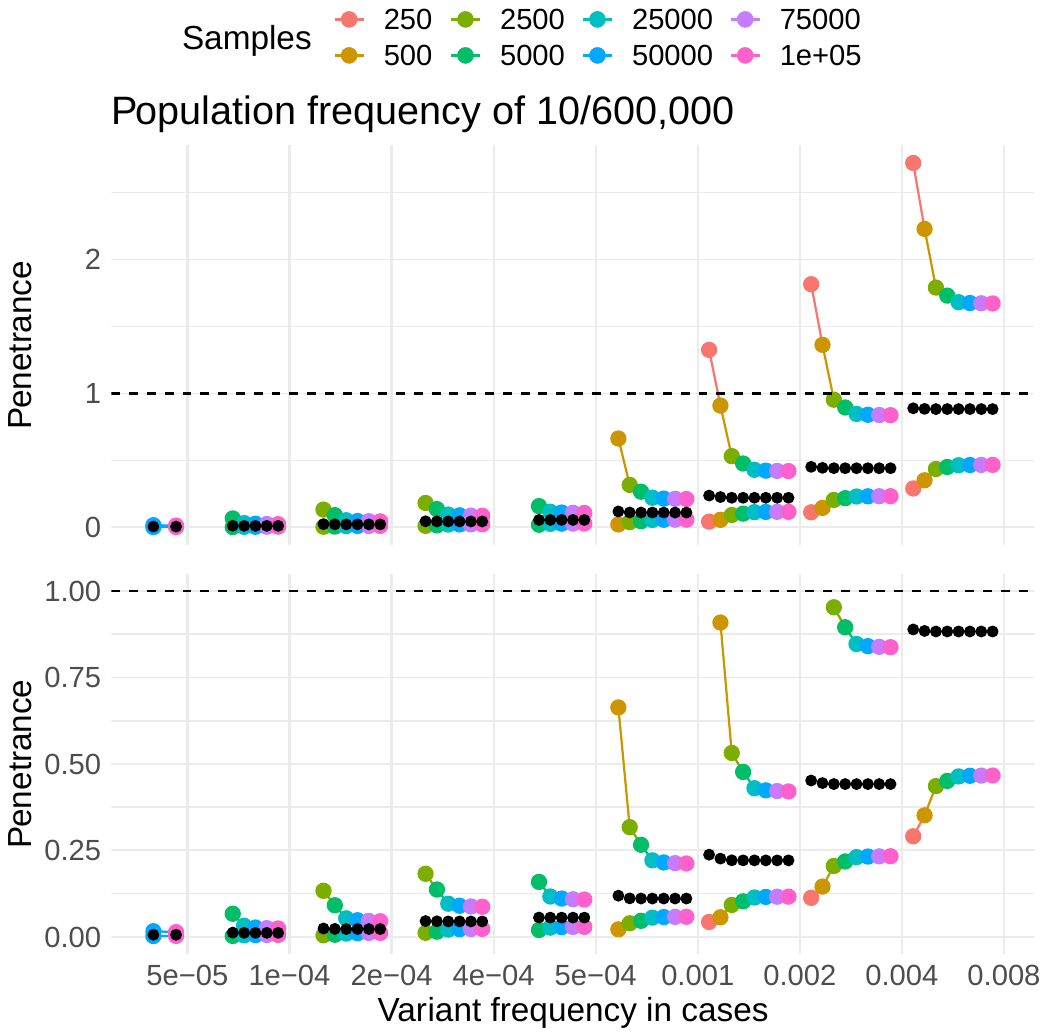

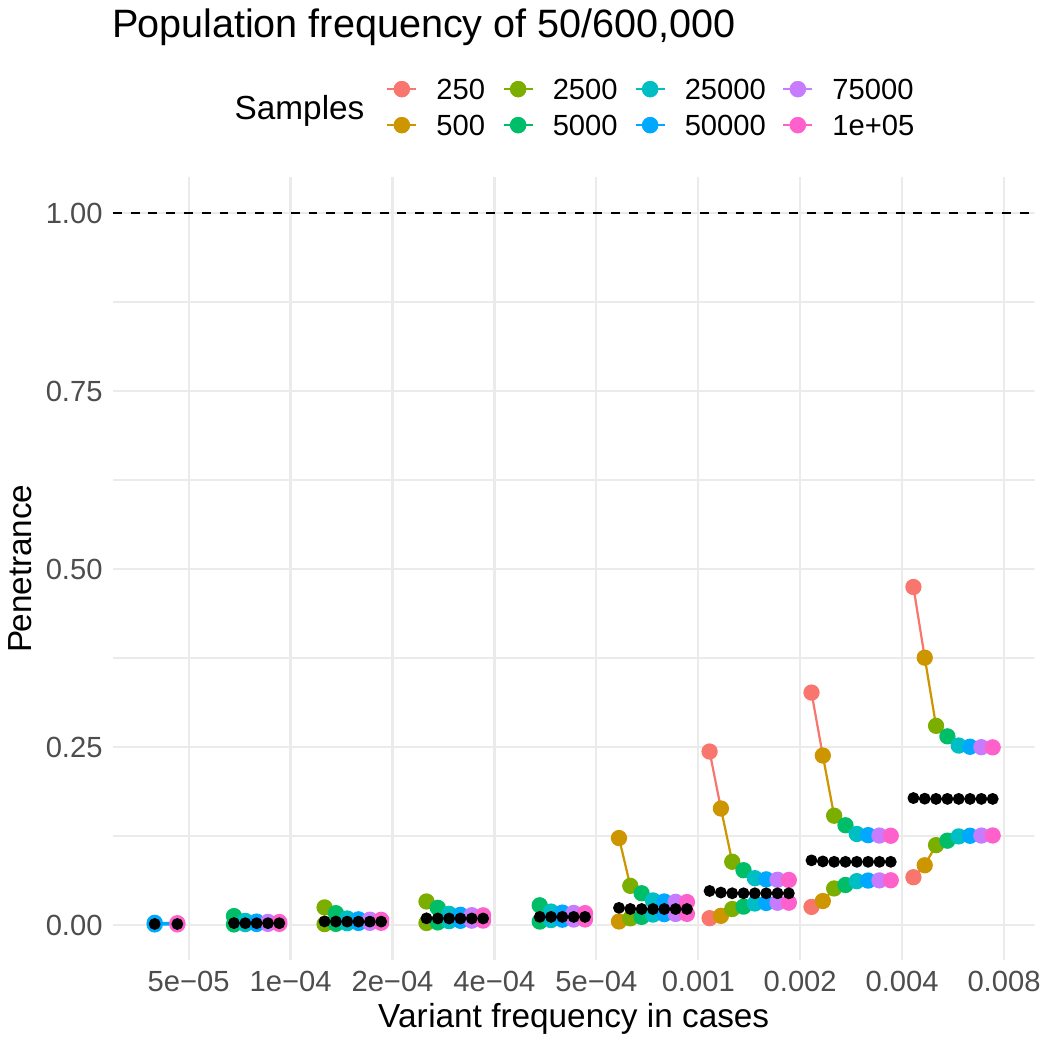

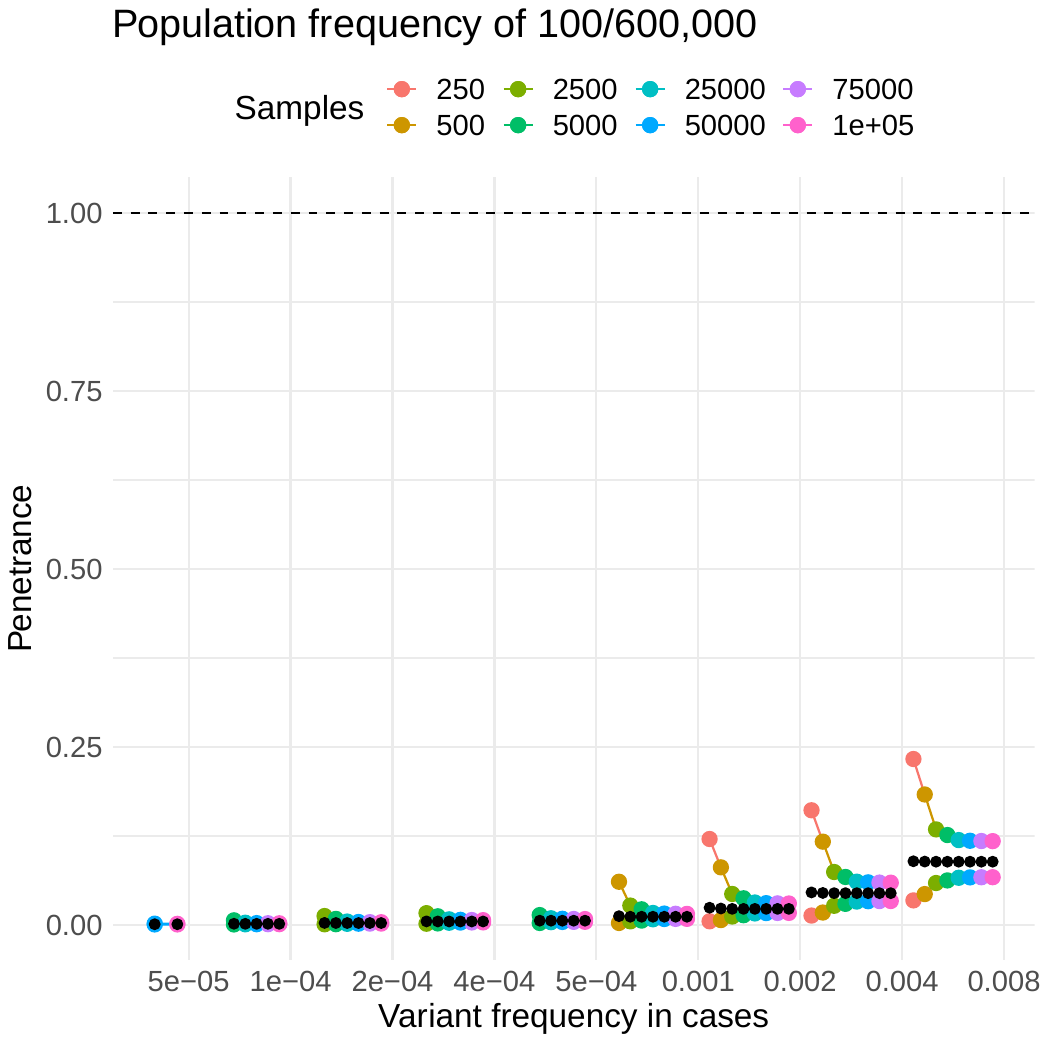

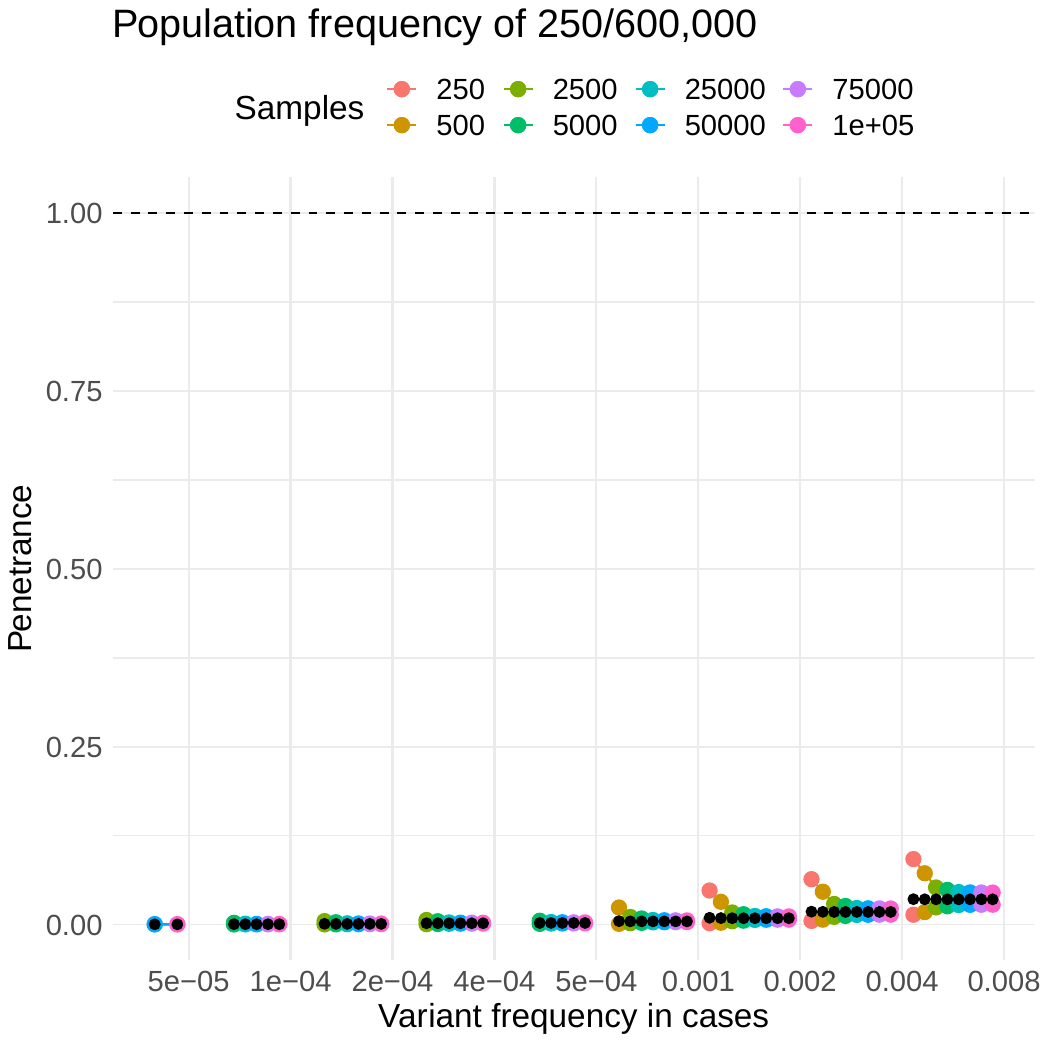

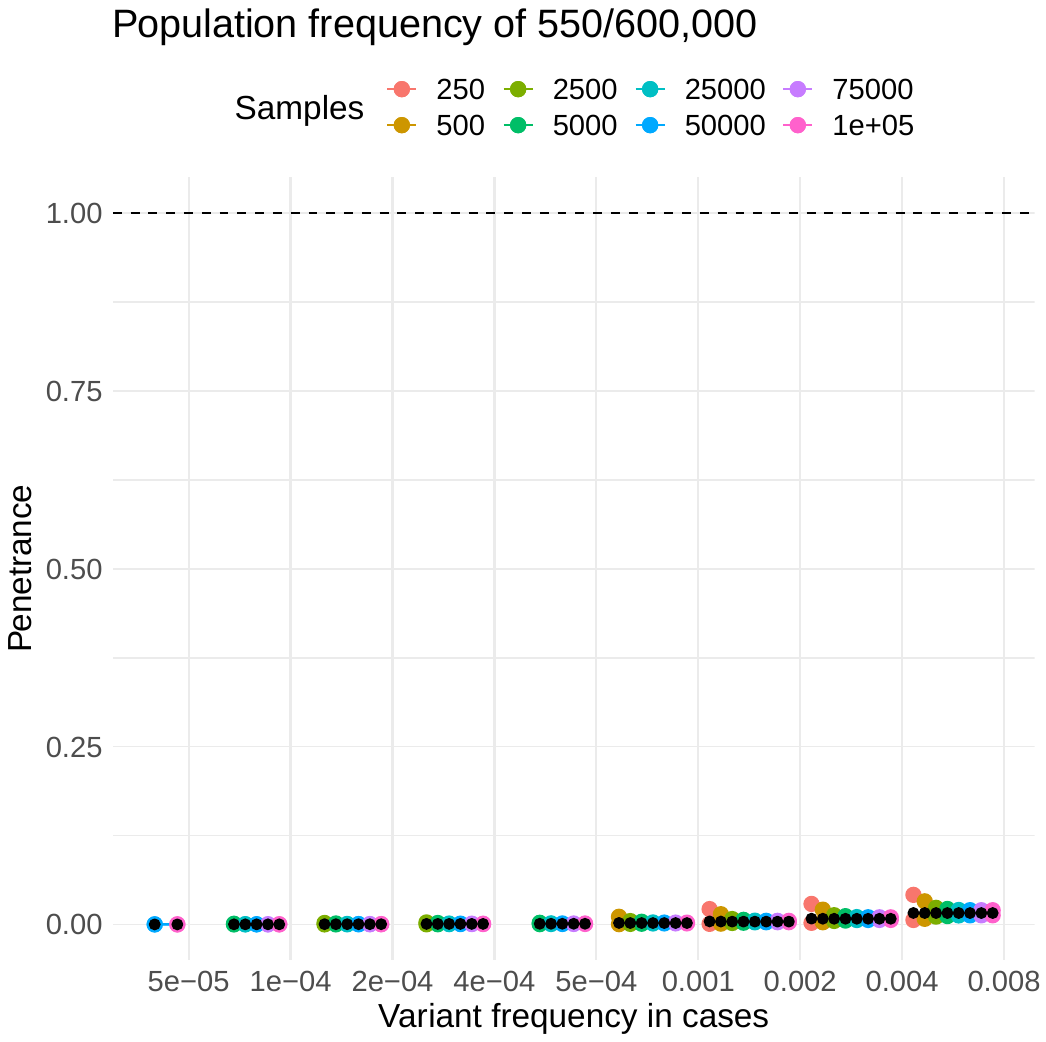

Figure S9 As the probability of the allele increases, precision increases, and the estimate of penetrance decreases.

Example variants had a population AC/AN (depicted as title of each plot) where popAF=0.000003-0.0009. Y-axis, estimates of penetrance; x-axis, caseAF=0.0001–0.008. The grey horizontal dashed line denotes a penetrance of 1.0 or 100%. The size of cases cohort is depicted as coloured points and indicated by the legend. Probability of the allele ($\boldsymbol{p}_{\boldsymbol{A}}$).

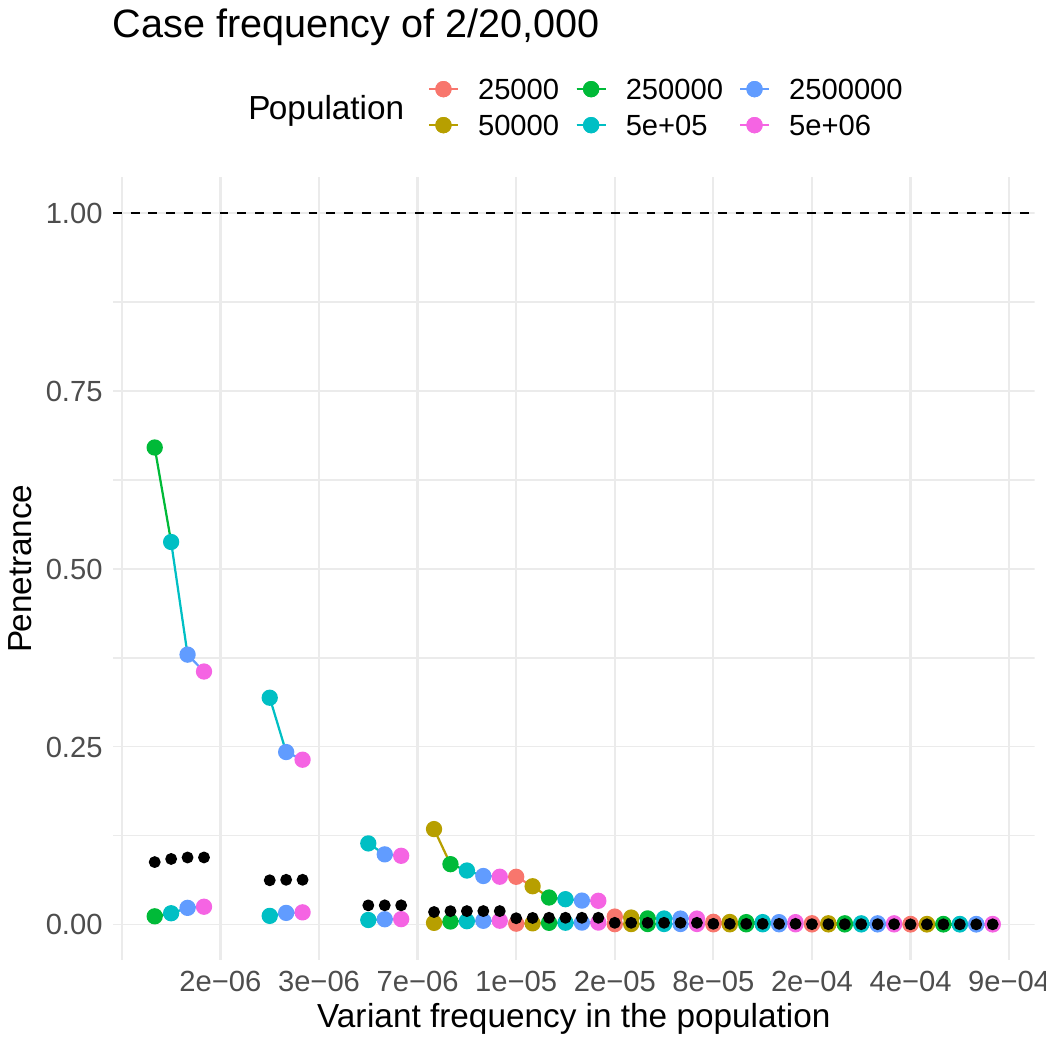

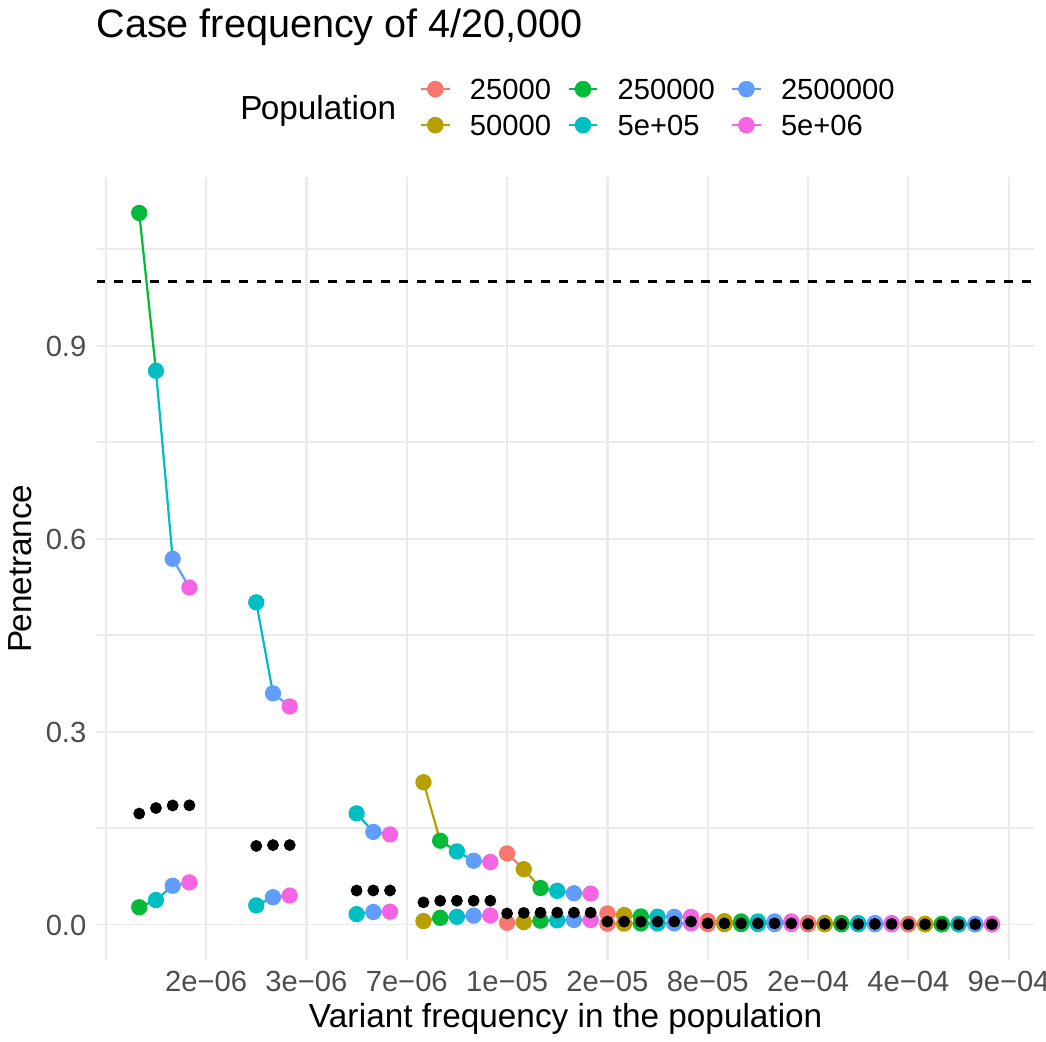

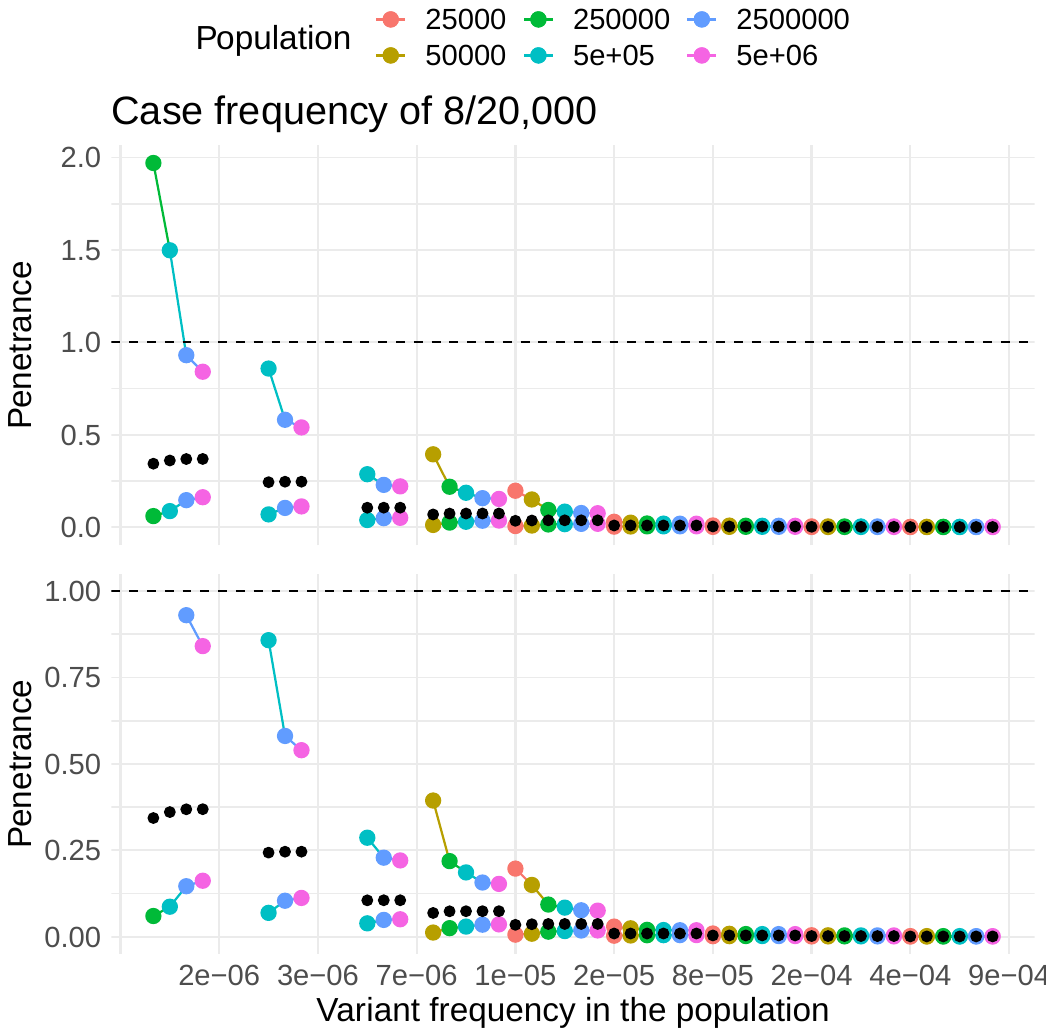

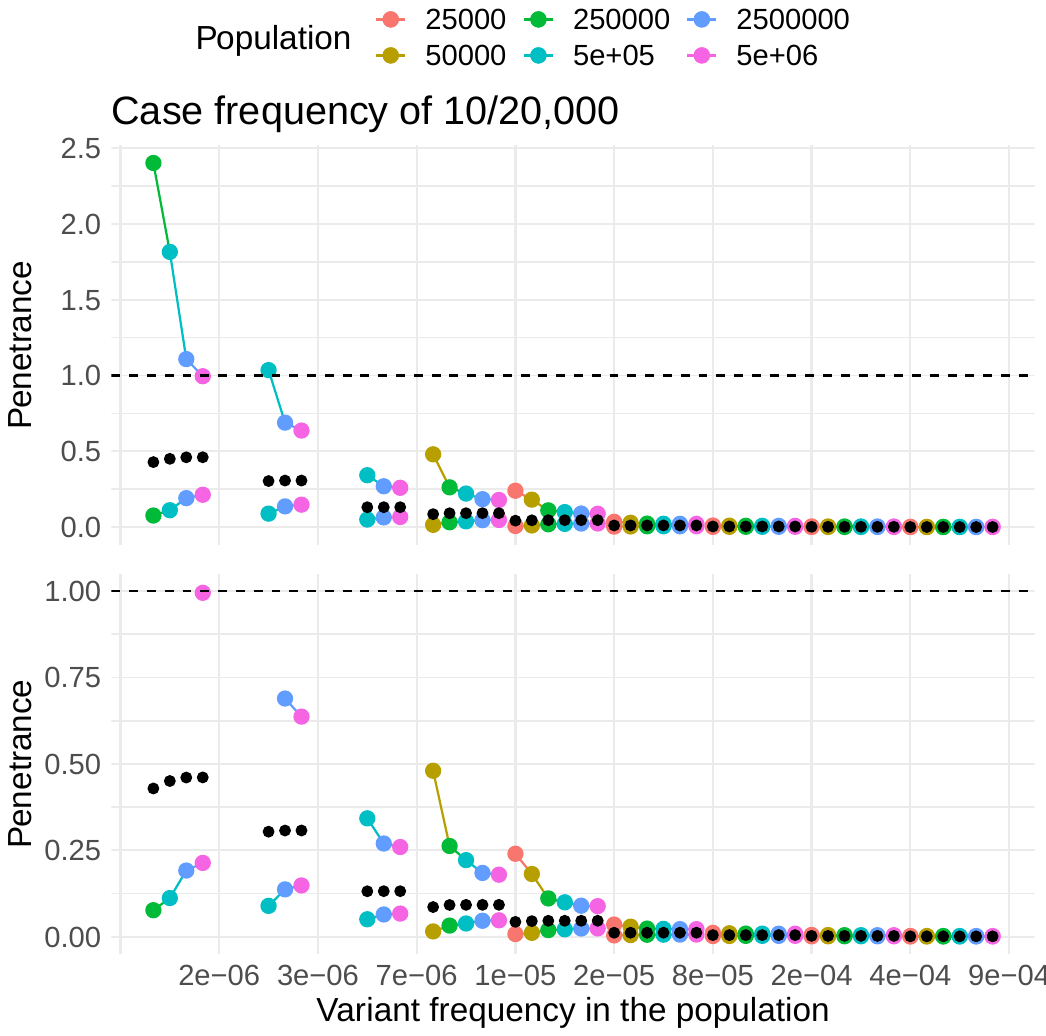

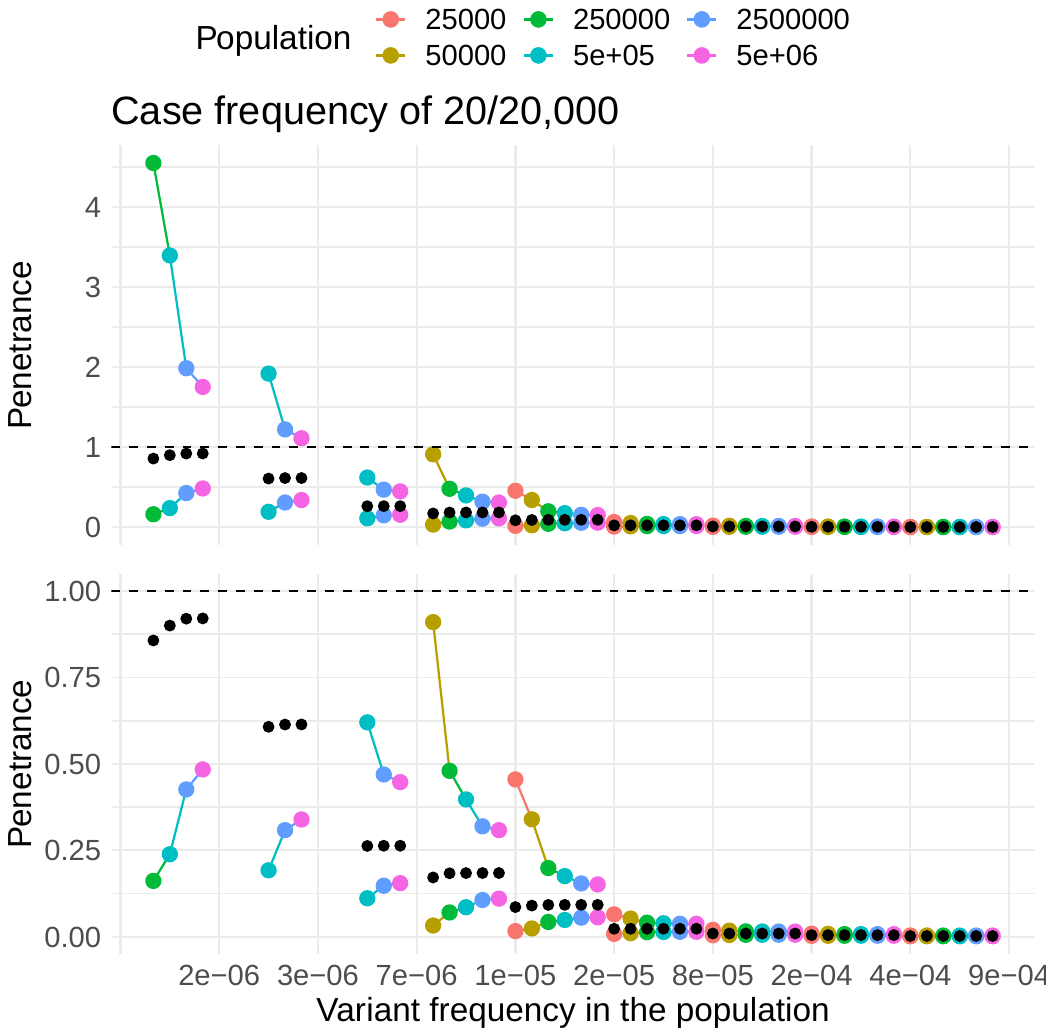

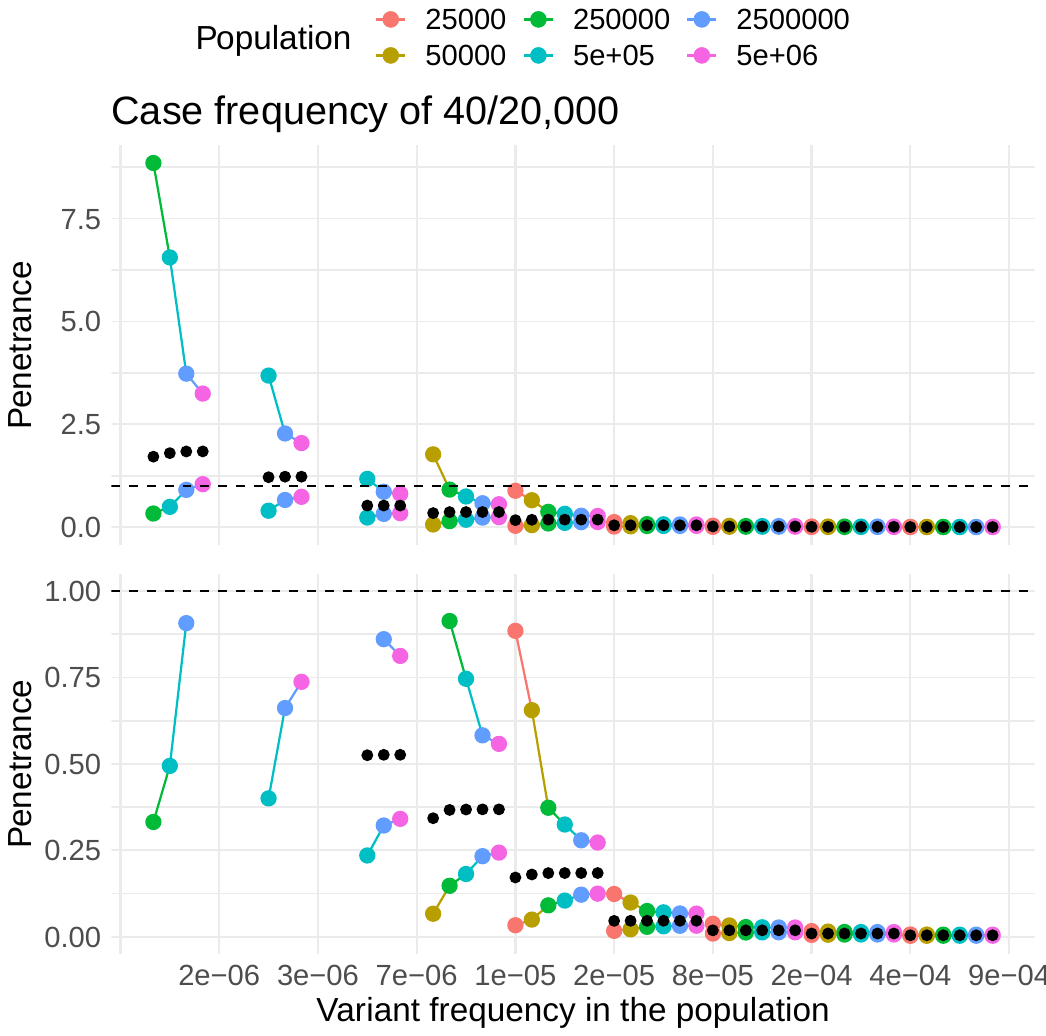

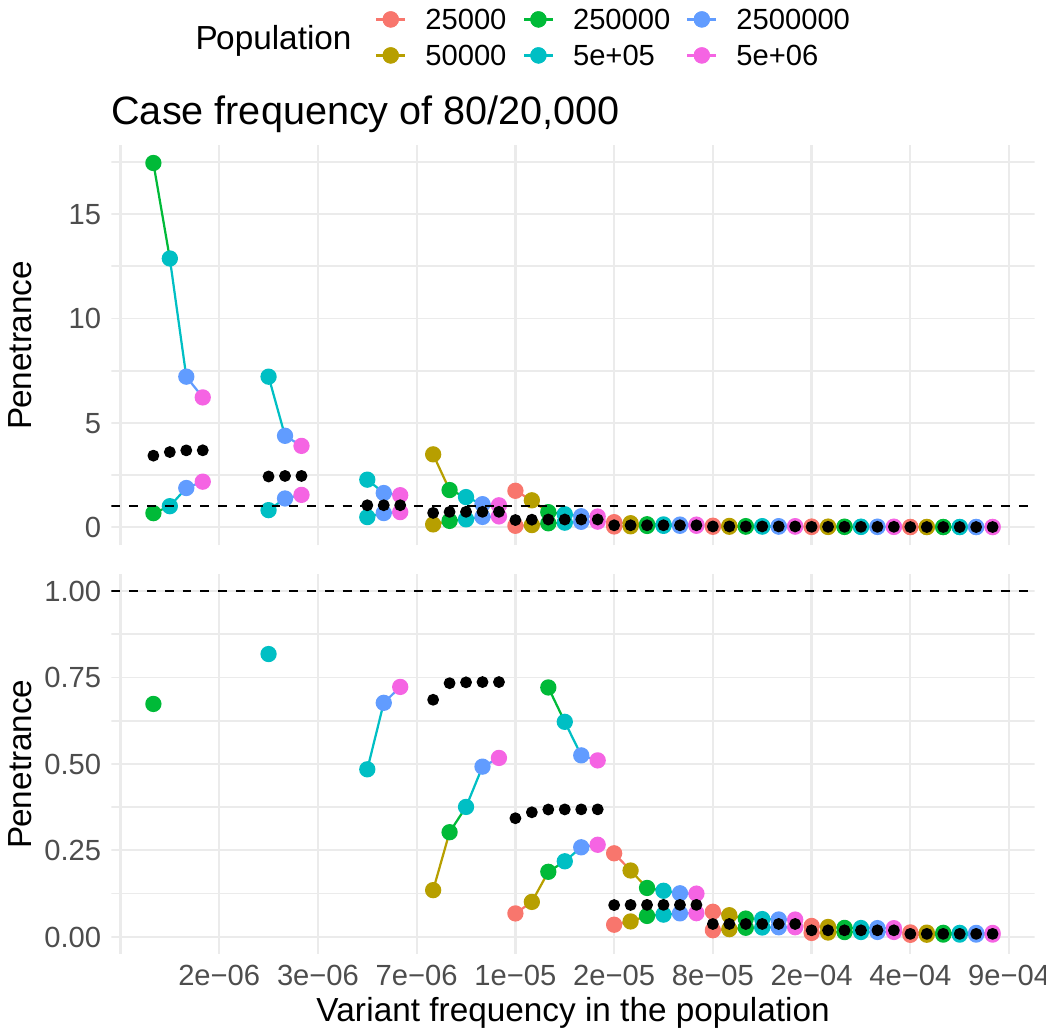

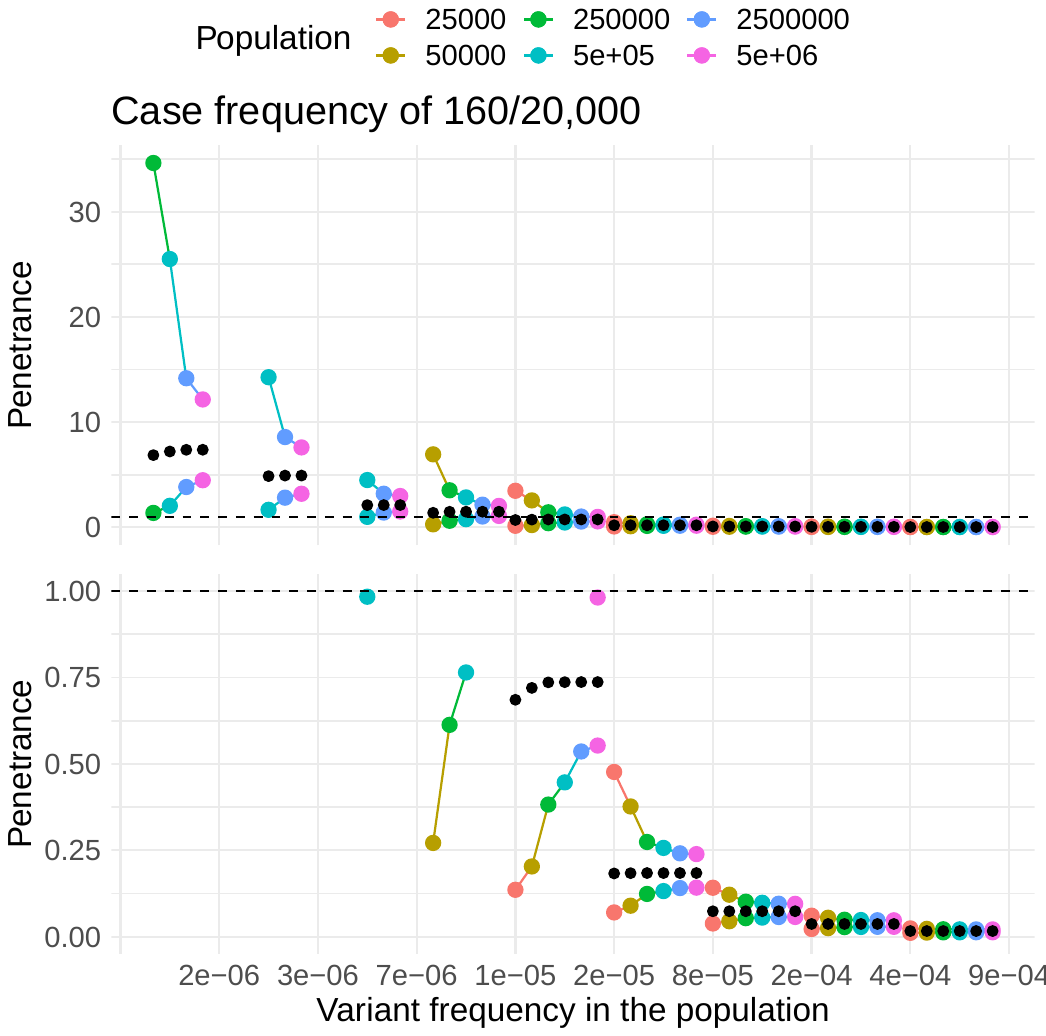

Figure S10 As the probability of the allele given disease increases, penetrance increases, and the precision of the estimate of penetrance has less confidence.

Example variants had a case AC/AN (depicted as title of each plot) where caseAF=0.0001–0.008. Y-axis, estimates of penetrance; x-axis, minor allele frequency in case cohorts ranging from 0.000003–0.0009. The grey horizontal dashed line denotes a penetrance of 1.0 or 100%. The size of population cohort is depicted as coloured points and indicated by the legend. Probability of the allele given disease ($\boldsymbol{p}_{\boldsymbol{A|D}}$).

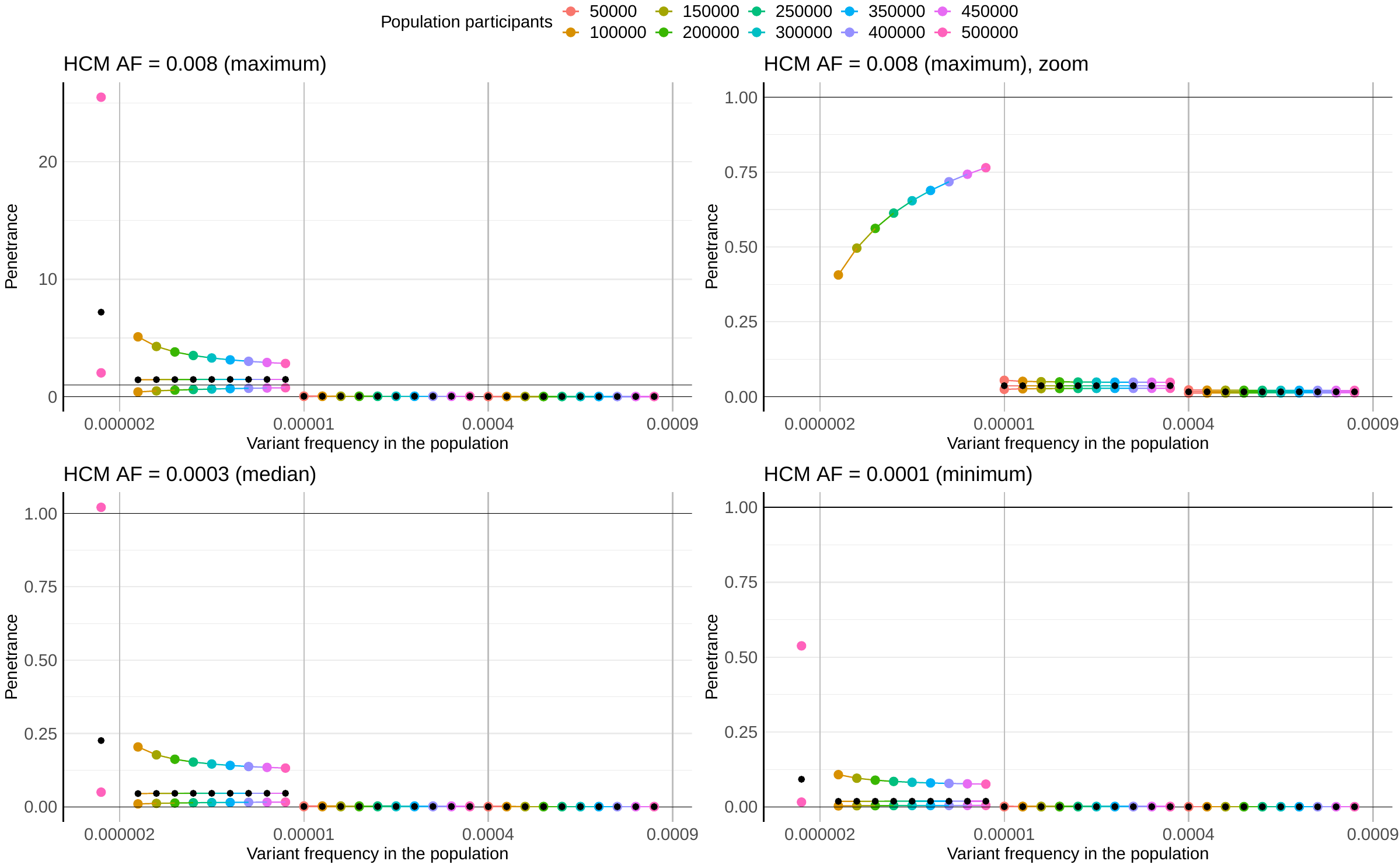

Figure S11 Simulations of the expected penetrance estimates in the range of the probability of the allele and the probability of the allele given disease, observed in this study.

The maximum $\boldsymbol{p}_{\boldsymbol{A|D}}$ observed was 0.008, the median was 0.0003, and the minimum was 0.0001 for the variants included in this study. Y-axis, estimates of penetrance; X-axis, four $\boldsymbol{p}_{\boldsymbol{A}}$ tested. The coloured points represented population reference sample size. The UCI is above the estimate of penetrance (black points) and the LCI below. The grey horizontal lines depict an estimated penetrance of 1.0 or 100%. While theoretically shown here, variants with a very high penetrance can have an estimated penetrance of >100, we did not observe any real variants in our dataset that had a combination of case and population allele frequencies that resulted in an estimated penetrance of > 100% (maximum penetrance was 66.8% for HCM, 78.6% for DCM).

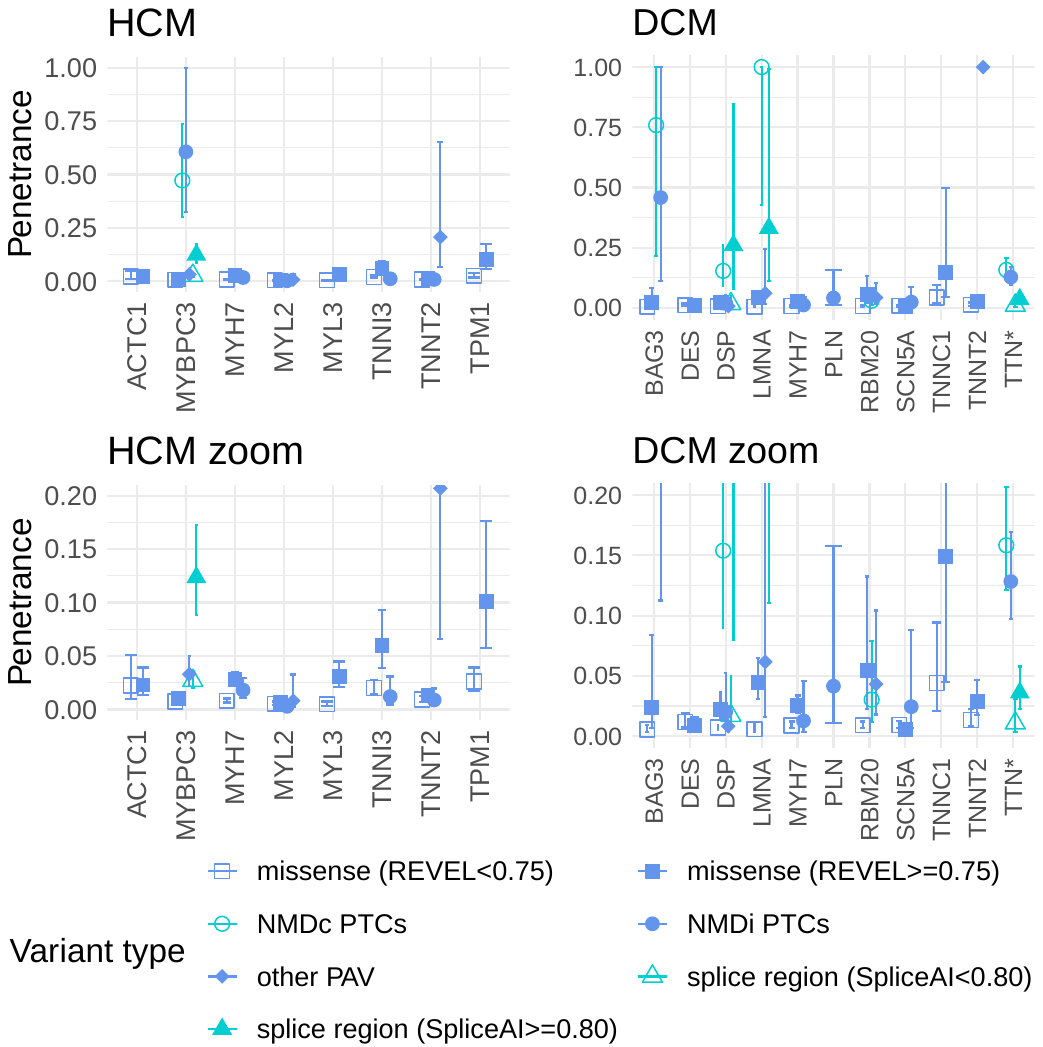

Figure S12 Aggregated penetrance of loss of function variants is highest (REVEL and SpliceAI softwares assessed).

The plot depicts estimated penetrance of rare variants in HCM-associated (left) and DCM-associated (right) genes. LoF and non-LoF variant groups are plotted in green and blue, respectively. LoF, predicted loss of function variants. This plot provides additional stratification for missense variants predicted as deleterious (using REVEL) and splice region variants predicted to be splice disrupting (using SpliceAI).

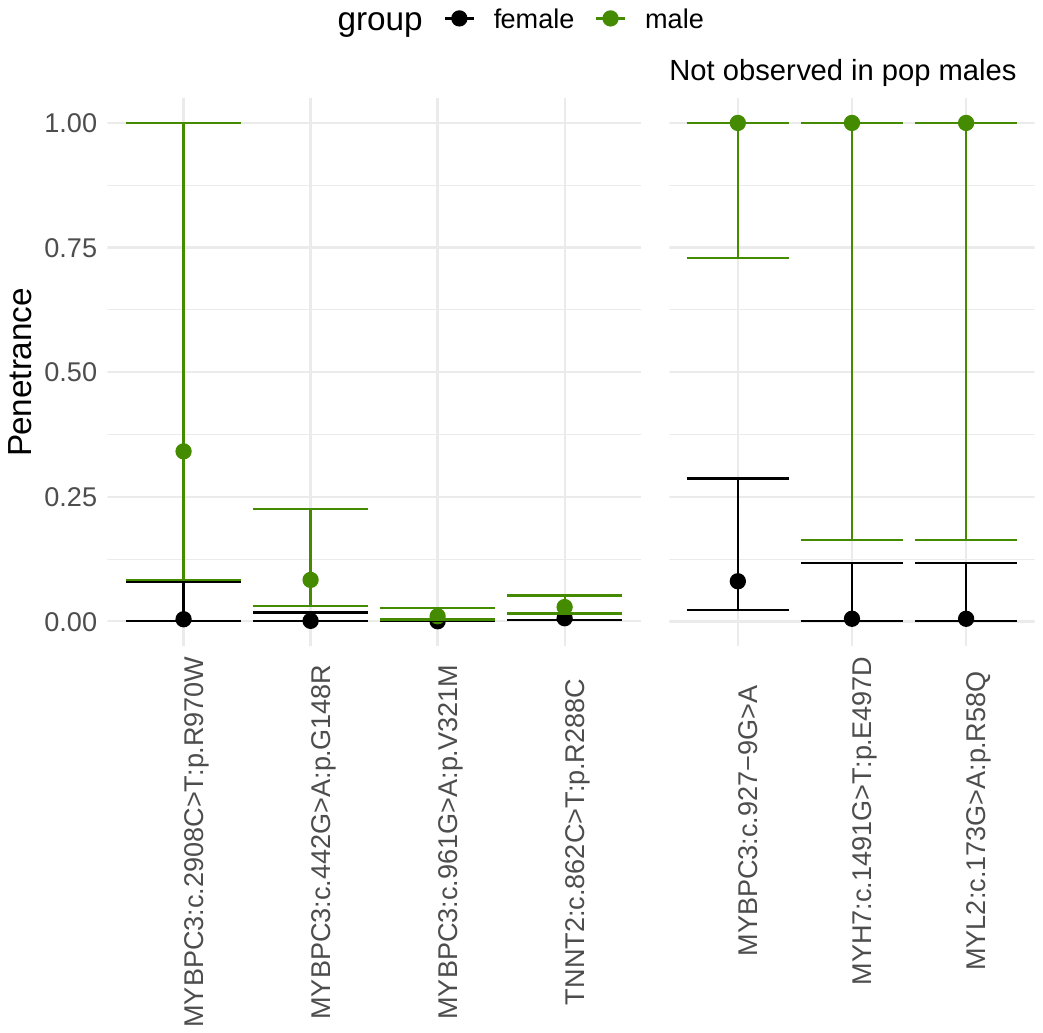

Figure S13 Variants with significantly decreased penetrance in females compared to males from Group 2.

The plot depicts the sex-specific estimates of penetrance of seven rare variants in HCM-associated genes with decreased penetrance in females. The variants are more common in females in our data. The variants on the right side of the plot were variants observed in male cases but not in males of the population reference datasets. Overlapping confidence intervals was observed for the sex-specific penetrance estimates of all other variants.

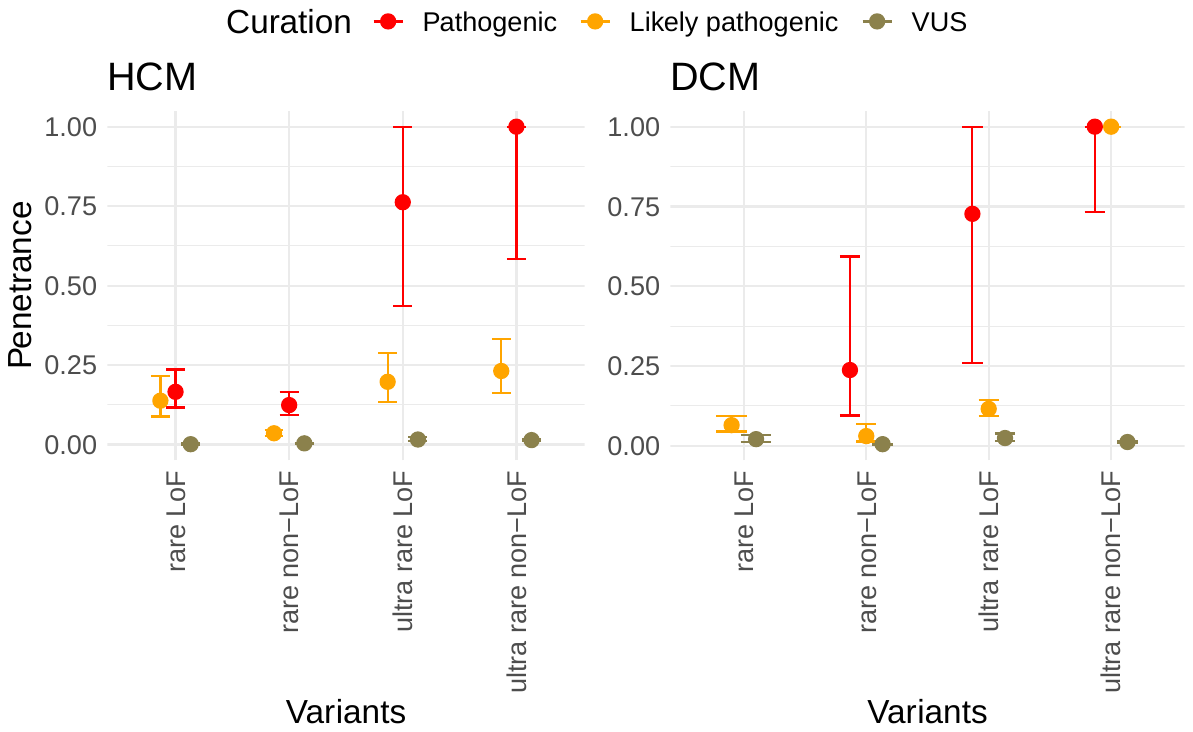

Figure S14 Aggregate penetrance of variants in CM-associated genes grouped by rarity and consequence.

This figure was used to inform the flow chart of the graphical abstract. The figure depicts ClinVar curation for four subgroups of variants based on rarity (ultra-rare, gnomAD AC = 0; rare, gnomAD AC > 0) and whether the variants are predicted to be loss of function (LoF, frameshift, stop gained, essential splice; non-LoF, missense, indels, start and stop lost). There was only one observation of one rare pathogenic LoF variant for DCM (PLN:c.116T>G:p.L39*; DCM AC = 1; gnomAD AC = 4), thus penetrance could not be estimated for this subgroup. 50% of the LP ultra-rare non-LoF group for DCM consisted of variants in *LMNA*.

### 6 References

1.	Minikel EV, Vallabh SM, Lek M, Estrada K, Samocha KE, Sathirapongsasuti JF, McLean CY, Tung JY, Yu LPC, Gambetti P, Blevins J, Zhang S, Cohen Y, Chen W, Yamada M, Hamaguchi T, Sanjo N, Mizusawa H, Nakamura Y, Kitamoto T, Collins SJ, Boyd A, Will RG, Knight R, Ponto C, Zerr I, Kraus TFJ, Eigenbrod S, Giese A, Calero M, et al. Quantifying prion disease penetrance using large population control cohorts. *Sci Transl Med* 2016;**8**:322ra9.

2.	Newcombe RG. Interval estimation for the difference between independent proportions: comparison of eleven methods. *Stat Med* England; 1998;**17**:873–890.

3.	Newcombe RG. Two-sided confidence intervals for the single proportion: Comparison of seven methods. *Stat Med* 1998;**17**:857–872.

4.	Brown LD, Cai TT, DasGupta A. Interval Estimation for a Binomial Proportion. *Stat Sci* Institute of Mathematical Statistics; 2001;**16**:101–117.

5.	Fagerland MW, Newcombe RG. Confidence intervals for odds ratio and relative risk based on the inverse hyperbolic sine transformation. *Stat Med* John Wiley & Sons; 2013;**32**:2823–2836.

6.	Denison DGT, Christopher CH, Bani KM, Adrian FMS. Bayesian Methods for Nonlinear Classification and Regression. John Wiley & Sons; 2002.

7.	Stuart A, Ord JK, Arnold SF. Kendalls’ Advanced Theory of Statistics, Vol. 1: Distribution Theory, 6th Edition. John Wiley & Sons; 2010.

8.	Lewis CM. Genetic association studies: design, analysis and interpretation. Brief. Bioinform. Oxford Academic; 2002. p. 146–153.

9.	Hershberger RE, Hedges DJ, Morales A. Dilated cardiomyopathy: The complexity of a diverse genetic architecture. *Nat Rev Cardiol* 2013;**10**:531–547.

10.	Haggerty CM, Damrauer SM, Levin MG, Birtwell D, Carey DJ, Golden AM, Hartzel DN, Hu Y, Judy R, Kelly MA, Kember RL, Lester Kirchner H, Leader JB, Liang L, Mcdermott-Roe C, Babu A, Morley M, Nealy Z, Person TN, Pulenthiran A, Small A, Smelser DT, Stahl RC, Sturm AC, Williams H, Baras A, Margulies KB, Cappola TP, Dewey FE, Verma A, et al. Genomics-First Evaluation of Heart Disease Associated With Titin-Truncating Variants. *Circulation* 2019;**140**:42–54.

11.	Torp A. Incidence of congestive cardiomyopathy. *Postgrad Med J* 1978;**54**:435–437.

12.	Codd MB, Sugrue DD, Gersh BJ, Melton LJ. Epidemiology of idiopathic dilated and hypertrophic cardiomyopathy: A population-based study in Olmsted County, Minnesota, 1975-1984. *Circulation* 1989;**80**:564–572.

13.	Bagger JP, Baandrup U, Rasmussen K, Møller M, Vesterlund T. Cardiomyopathy in western Denmark. *Br Heart J* 1984;**52**:327–331.

14.	Miura K, Nakagawa H, Morikawa Y, Sasayama S, Matsumori A, Hasegawa K, Ohno Y, Tamakoshi A, Kawamura T, Inaba Y. Epidemiology of idiopathic cardiomyopathy in Japan: Results from a nationwide survey. *Heart* 2002;**87**:126–130.

15.	Lannou S, Mansencal N, Couchoud C, Lassalle M, Dubourg O, Stengel B, Jacquelinet C, Charron P. The Public Health Burden of Cardiomyopathies: Insights from a Nationwide Inpatient Study. *J Clin Med* 2020;**9**:920.

16.	Williams DG, Olsen EGJ. Prevalence of overt dilated cardiomyopathy in two regions of England. *Br Heart J* 1985;**54**:153–155.

17.	Razali R, Danuri N, Ibrahim ZO, Peng HB, Yusoff K. A4530 Prevalence of dilated cardiomyopathy in Malaysia population by echocardiographic screening. *J Hypertens* 2018;**36**:e163–e164.

18.	Rakar S, Sinagra G, Lenarda A Di, Poletti A, Bussani R, Silvestri F, Camerini F, Alberti E, Lardieri G, Mestroni L, Morgera T, Pinamonte B, Salvi A, D’Ambrosio A, Gregori D, Perkan A, Zecchin M. Epidemiology of dilated cardiomyopathy. A prospective post-mortem study of 5252 necropsies. *Eur Heart J* 1997;**18**:117–123.

19.	Pirruccello JP, Bick A, Wang M, Chaffin M, Friedman S, Yao J, Guo X, Venkatesh BA, Taylor KD, Post WS, Rich S, Lima JAC, Rotter JI, Philippakis A, Lubitz SA, Ellinor PT, Khera A V., Kathiresan S, Aragam KG. Analysis of cardiac magnetic resonance imaging in 36,000 individuals yields genetic insights into dilated cardiomyopathy. *Nat Commun* 2020;**11**:2254.

20.	Marvao A de, McGurk KA, Zheng SL, Thanaj M, Bai W, Duan J, Biffi C, Mazzarotto F, Statton B, Dawes TJW, Savioli N, Halliday BP, Xu X, Buchan RJ, Baksi AJ, Quinlan M, Tokarczuk P, Tayal U, Francis C, Whiffin N, Theotokis PI, Zhang X, Jang M, Berry A, Pantazis A, Barton PJR, Rueckert D, Prasad SK, Walsh R, Ho CY, et al. Phenotypic Expression and Outcomes in Individuals With Rare Genetic Variants of Hypertrophic Cardiomyopathy. *J Am Coll Cardiol* 2021;**78**:1097–1110.

21.	Petersen SE, Aung N, Sanghvi MM, Zemrak F, Fung K, Paiva JM, Francis JM, Khanji MY, Lukaschuk E, Lee AM, Carapella V, Kim YJ, Leeson P, Piechnik SK, Neubauer S. Reference ranges for cardiac structure and function using cardiovascular magnetic resonance (CMR) in Caucasians from the UK Biobank population cohort. *J Cardiovasc Magn Reson* 2017;**19**:18.

22.	Bai W, Suzuki H, Huang J, Francis C, Wang S, Tarroni G, Guitton F, Aung N, Fung K, Petersen SE, Piechnik SK, Neubauer S, Evangelou E, Dehghan A, O’Regan DP, Wilkins MR, Guo Y, Matthews PM, Rueckert D. A population-based phenome-wide association study of cardiac and aortic structure and function. *Nat Med* 2020;**26**:1654–1662.

23.	Husser D, Ueberham L, Jacob J, Heuer D, Riedel-Heller S, Walker J, Hindricks G, Bollmann A. Prevalence of clinically apparent hypertrophic cardiomyopathy in Germany—An analysis of over 5 million patients. *PLoS One* 2018;**13**:1–8.

24.	Moon I, Lee SY, Kim HK, Han K Do, Kwak S, Kim M, Lee HJ, Hwang IC, Lee H, Park JB, Yoon YE, Kim YJ, Cho GY. Trends of the prevalence and incidence of hypertrophic cardiomyopathy in Korea: A nationwide population-based cohort study. *PLoS One* 2020;**15**:1–10.

25.	Zou Y, Song L, Wang Z, Ma A, Liu T, Gu H, Lu S, Wu P, Zhang Y, Shen L, Cai Y, Zhen Y, Liu Y, Hui R. Prevalence of idiopathic hypertrophic cardiomyopathy in China: A population-based echocardiographic analysis of 8080 adults. *Am J Med* 2004;**116**:14–18.

26.	Maron BJ, Gardin JM, Flack JM, Gidding SS, Kurosaki TT, Bild DE. Prevalence of Hypertrophic Cardiomyopathy in a General Population of Young Adults. *Circulation* 1995;**92**:785–789.

27.	Massera D, McClelland RL, Ambale-Venkatesh B, Gomes AS, Hundley WG, Kawel-Boehm N, Yoneyama K, Owens DS, Garcia MJ, Sherrid M V., Kizer JR, Lima JAC, Bluemke DA. Prevalence of Unexplained Left Ventricular Hypertrophy by Cardiac Magnetic Resonance Imaging in MESA. *J Am Heart Assoc* 2019;**8**.

28.	Magnusson P, Palm A, Branden E, Mörner S. Misclassification of hypertrophic cardiomyopathy: Validation of diagnostic codes. *Clin Epidemiol* 2017;**9**:403–410.

29.	Maron BJ, Mathenge R, Casey SA, Poliac LC, Longe TF. Clinical profile of hypertrophic cardiomyopathy identified de novo in rural communities. *J Am Coll Cardiol* Elsevier Masson SAS; 1999;**33**:1590–1595.

30.	Hada Y, Sakamoto T, Amano K, Yamaguchi T, Takenaka K, Takahashi H, Takikawa R, Hasegawa I, Takahashi T, Suzuki JI, Sugimoto T, Saito KI. Prevalence of hypertrophic cardiomyopathy in a population of adult japanese workers as detected by echocardiographic screening. *Am J Cardiol* 1987;**59**:183–184.

31.	Maro EE, Janabi M, Kaushik R. Clinical and echocardiographic study of hypertrophic cardiomyopathy in Tanzania. *Trop Doct* 2006;**36**:225–227.

32.	Nistri S, Thiene G, Basso C, Corrado D, Vitolo A, Maron BJ. Screening for hypertrophic cardiomyopathy in a young male military population. *Am J Cardiol* 2003;**91**:1021–1023.

33.	Basavarajaiah S, Wilson M, Whyte G, Shah A, McKenna W, Sharma S. Prevalence of Hypertrophic Cardiomyopathy in Highly Trained Athletes. Relevance to Pre-Participation Screening. *J Am Coll Cardiol* 2008;**51**:1033–1039.

34.	Aronow WS, Kronzon I. Prevalence of hypertrophic cardiomyopathy and its association with mitral anular calcium in elderly patients. *Chest* 1988;**94**:1295–1296.

35.	Shapiro LM, Zezulka A. Hypertrophic cardiomyopathy: A common disease with a good prognosis. Five year experience of a district general hospital. *Heart* 1983;**50**:530–533.

36.	Maron MS, Hellawell JL, Lucove JC, Farzaneh-Far R, Olivotto I. Occurrence of clinically diagnosed hypertrophic cardiomyopathy in the United States. *Am J Cardiol* 2016;**117**:1651–1654.

37.	Maron BJ, Spirito P, Roman MJ, Paranicas M, Okin PM, Best LG, Lee ET, Devereux RB. Prevalence of hypertrophic cardiomyopathy in a population-based sample of American Indians aged 51 to 77 years (the Strong Heart Study). *Am J Cardiol* 2004;**93**:1510–1514.

38.	Balduzzi S, Rücker G, Schwarzer G. How to perform a meta-analysis with R: A practical tutorial. *Evid Based Ment Health* 2019;**22**:153–160.

39.	Viechtbauer W. Conducting Meta-Analyses in R with the metafor Package. *J Stat Softw* 2010;**36**:1–48.

40.	Agresti A, Coull BA. Approximate Is Better than ‘Exact’ for Interval Estimation of Binomial Proportions. *Am Stat* JSTOR; 1998;**52**:119.

41.	Katz D, Baptista J, Azen SP, Pike MC. Obtaining Confidence Intervals for the Risk Ratio in Cohort Studies. *Biometrics* [Wiley, International Biometric Society]; 1978;**34**:469–474.

42.	Pettigrew HM, Gart JJ, Thomas DG. The Bias and Higher Cumulants of the Logarithm of a Binomial Variate. *Biometrika* [Oxford University Press, Biometrika Trust]; 1986;**73**:425–435.

43.	Gart JJ, Nam JM. Approximate interval estimation of the difference in binomial parameters: correction for skewness and extension to multiple tables. *Biometrics* United States; 1990;**46**:637–643.

44.	Whiffin N, Minikel E, Walsh R, O’Donnell-Luria AH, Karczewski K, Ing AY, Barton PJR, Funke B, Cook SA, Macarthur D, Ware JS. Using high-resolution variant frequencies to empower clinical genome interpretation. *Genet Med* 2017;**19**:1151–1158.

45.	Sudlow C, Gallacher J, Allen N, Beral V, Burton P, Danesh J, Downey P, Elliott P, Green J, Landray M, Liu B, Matthews P, Ong G, Pell J, Silman A, Young A, Sprosen T, Peakman T, Collins R. UK Biobank: An Open Access Resource for Identifying the Causes of a Wide Range of Complex Diseases of Middle and Old Age. *PLOS Med* Public Library of Science; 2015;**12**:e1001779.

46.	Hout CV Van, Tachmazidou I, Backman JD, Hoffman JX, Ye B, Pandey AK, Gonzaga-Jauregui C, Khalid S, Liu D, Banerjee N, Li AH, Colm O, Marcketta A, Staples J, Schurmann C, Hawes A, Maxwell E, Barnard L, Lopez A, Penn J, Habegger L, Blumenfeld AL, Yadav A, Praveen K, Jones M, Salerno WJ, Chung WK, Surakka I, Willer CJ, Hveem K, et al. Whole exome sequencing and characterization of 49,960 individuals in the UK Biobank. *Nature* 2020;**586**:749–756.

47.	Bycroft C, Freeman C, Petkova D, Band G, Elliott LT, Sharp K, Motyer A, Vukcevic D, Delaneau O, O’Connell J, Cortes A, Welsh S, Young A, Effingham M, McVean G, Leslie S, Allen N, Donnelly P, Marchini J. The UK Biobank resource with deep phenotyping and genomic data. *Nature* 2018;**562**:203–209.

48.	Karczewski KJ, Francioli LC, Tiao G, Cummings BB, Alföldi J, Wang Q, Collins RL, Laricchia KM, Ganna A, Birnbaum DP, Gauthier LD, Brand H, Solomonson M, Watts NA, Rhodes D, Singer-Berk M, England EM, Seaby EG, Kosmicki JA, Walters RK, Tashman K, Farjoun Y, Banks E, Poterba T, Wang A, Seed C, Whiffin N, Chong JX, Samocha KE, Pierce-Hoffman E, et al. The mutational constraint spectrum quantified from variation in 141,456 humans. *Nature* 2020;**581**:434–443.

49.	McLaren W, Gil L, Hunt SE, Riat HS, Ritchie GRS, Thormann A, Flicek P, Cunningham F. The Ensembl Variant Effect Predictor. *Genome Biol* 2016;**17**:122.

50.	Mazzarotto F, Tayal U, Buchan RJ, Midwinter W, Wilk A, Whiffin N, Govind R, Mazaika E, Marvao A de, Dawes TJW, Felkin LE, Ahmad M, Theotokis PI, Edwards E, Ing AY, Thomson KL, Chan LLH, Sim D, Baksi AJ, Pantazis A, Roberts AM, Watkins H, Funke B, O’Regan DP, Olivotto I, Barton PJR, Prasad SK, Cook SA, Ware JS, Walsh R. Reevaluating the Genetic Contribution of Monogenic Dilated Cardiomyopathy. *Circulation* American Heart Association; 2020;**141**:387–398.

51.	Walsh R, Thomson KL, Ware JS, Funke BH, Woodley J, McGuire KJ, Mazzarotto F, Blair E, Seller A, Taylor JC, Minikel E V, MacArthur DG, Farrall M, Cook SA, Watkins H, Consortium EA. Reassessment of Mendelian gene pathogenicity using 7,855 cardiomyopathy cases and 60,706 reference samples. *Genet Med* 2017;**19**:192–203.

52.	Roberts AM, Ware JS, Herman DS, Schafer S, Baksi J, Bick AG, Buchan RJ, Walsh R, John S, Wilkinson S, Mazzarotto F, Felkin LE, Gong S, Macarthur JAL, Cunningham F, Flannick J, Gabriel SB, Altshuler DM, MacDonald PS, Heinig M, Keogh AM, Hayward CS, Banner NR, Pennell DJ, O’Regan DP, San TR, Marvao A De, Dawes TJW, Gulati A, Birks EJ, et al. Integrated allelic, transcriptional, and phenomic dissection of the cardiac effects of titin truncations in health and disease. *Sci Transl Med* 2015;**7**:270ra6.

53.	Pua CJ, Tham N, Chin CWL, Walsh R, Khor CC, Toepfer CN, Repetti GG, Garfinkel AC, Ewoldt JF, Cloonan P, Chen CS, Lim SQ, Cai J, Loo LY, Kong SC, Chiang CWK, Whiffin N, Marvao A de, Lio PM, Hii AA, Yang CX, Le TT, Bylstra Y, Lim WK, Teo JX, Padilha K, Silva G V, Pan B, Govind R, Buchan RJ, et al. Genetic Studies of Hypertrophic Cardiomyopathy in Singaporeans Identify Variants in TNNI3 and TNNT2 That Are Common in Chinese Patients. *Circ Genomic Precis Med* United States; 2020;**13**:424–434.

54.	Aguib Y, Allouba M, Afify A, Halawa S, El-Khatib M, Sous M, Galal A, Abdelrahman E, Shehata N, Sawy A El, Elmaghawry M, Anwer S, Kamel O, Mozy W El, Khedr H, Kharabish A, Thabet N, Theotokis PI, Buchan R, Govind R, Whiffin N, Walsh R, Aguib H, Elguindy A, O’Regan DP, Cook SA, Barton PJ, Ware JS, Yacoub M. The Egyptian Collaborative Cardiac Genomics (ECCO-GEN) Project: defining a healthy volunteer cohort. *npj Genomic Med* 2020;**5**:46.

55.	Aguib Y, Allouba M, Walsh R, Ibrahim AM, Halawa S, Afify A, Hosny M, Theotokis PI, Galal A, Elshorbagy S, Roshdy M, Kassem HS, Ellithy A, Buchan R, Whiffin N, Anwer S, Cook SA, Moustafa A, ElGuindy A, Ware JS, Barton PJR, Yacoub M. New Variant With a Previously Unrecognized Mechanism of Pathogenicity in Hypertrophic Cardiomyopathy. *Circulation* American Heart Association; 2021;**144**:754–757.

56.	Alfares AA, Kelly MA, McDermott G, Funke BH, Lebo MS, Baxter SB, Shen J, McLaughlin HM, Clark EH, Babb LJ, Cox SW, Depalma SR, Ho CY, Seidman JG, Seidman CE, Rehm HL. Results of clinical genetic testing of 2,912 probands with hypertrophic cardiomyopathy: Expanded panels offer limited additional sensitivity. *Genet Med* 2015;**17**:880–888.

57.	Pugh TJ, Kelly MA, Gowrisankar S, Hynes E, Seidman MA, Baxter SM, Bowser M, Harrison B, Aaron D, Mahanta LM, Lakdawala NK, McDermott G, White ET, Rehm HL, Lebo M, Funke BH. The landscape of genetic variation in dilated cardiomyopathy as surveyed by clinical DNA sequencing. *Genet Med* 2014;**16**:601–608.

58.	Walsh R, Mazzarotto F, Whiffin N, Buchan R, Midwinter W, Wilk A, Li N, Felkin L, Ingold N, Govind R, Ahmad M, Mazaika E, Allouba M, Zhang X, Marvao A De, Day SM, Ashley E, Colan SD, Michels M, Pereira AC, Jacoby D, Ho CY, Thomson KL, Watkins H, Barton PJR, Olivotto I, Cook SA, Ware JS. Quantitative approaches to variant classification increase the yield and precision of genetic testing in Mendelian diseases: The case of hypertrophic cardiomyopathy. *Genome Med* Genome Medicine; 2019;**11**:1–18.

59.	Alfares AA, Kelly MA, McDermott G, Funke BH, Lebo MS, Baxter SB, Shen J, McLaughlin HM, Clark EH, Babb LJ, Cox SW, DePalma SR, Ho CY, Seidman JG, Seidman CE, Rehm HL. Results of clinical genetic testing of 2,912 probands with hypertrophic cardiomyopathy: expanded panels offer limited additional sensitivity. *Genet Med* 2015;**17**:880–888.

60.	Landrum MJ, Lee JM, Riley GR, Jang W, Rubinstein WS, Church DM, Maglott DR. ClinVar: public archive of relationships among sequence variation and human phenotype. *Nucleic Acids Res* 2014;**42**:D980-5.

61.	Jaganathan K, Kyriazopoulou Panagiotopoulou S, McRae JF, Darbandi SF, Knowles D, Li YI, Kosmicki JA, Arbelaez J, Cui W, Schwartz GB, Chow ED, Kanterakis E, Gao H, Kia A, Batzoglou S, Sanders SJ, Farh KKH. Predicting Splicing from Primary Sequence with Deep Learning. *Cell* Elsevier; 2019;**176**:535–548.

62.	Ioannidis NM, Rothstein JH, Pejaver V, Middha S, McDonnell SK, Baheti S, Musolf A, Li Q, Holzinger E, Karyadi D, Cannon-Albright LA, Teerlink CC, Stanford JL, Isaacs WB, Xu J, Cooney KA, Lange EM, Schleutker J, Carpten JD, Powell IJ, Cussenot O, Cancel-Tassin G, Giles GG, MacInnis RJ, Maier C, Hsieh C-L, Wiklund F, Catalona WJ, Foulkes WD, Mandal D, et al. REVEL: An Ensemble Method for Predicting the Pathogenicity of Rare Missense Variants. *Am J Hum Genet* 2016;**99**:877–885.

63.	Purcell S, Neale B, Todd-Brown K, Thomas L, Ferreira MAR, Bender D, Maller J, Sklar P, Bakker PIW de, Daly MJ, Sham PC. PLINK: a tool set for whole-genome association and population-based linkage analyses. *Am J Hum Genet* 2007;**81**:559–575.

64.	Eilbeck K, Lewis SE, Mungall CJ, Yandell M, Stein L, Durbin R, Ashburner M. The Sequence Ontology: a tool for the unification of genome annotations. *Genome Biol* 2005;**6**:R44.

65.	Howe KL, Achuthan P, Allen J, Allen J, Alvarez-Jarreta J, Amode MR, Armean IM, Azov AG, Bennett R, Bhai J, Billis K, Boddu S, Charkhchi M, Cummins C, Da Rin Fioretto L, Davidson C, Dodiya K, El Houdaigui B, Fatima R, Gall A, Garcia Giron C, Grego T, Guijarro-Clarke C, Haggerty L, Hemrom A, Hourlier T, Izuogu OG, Juettemann T, Kaikala V, Kay M, et al. Ensembl 2021. *Nucleic Acids Res* 2021;**49**:D884–D891.

66.	Ingles J, Goldstein J, Thaxton C, Caleshu C, Corty EW, Crowley SB, Dougherty K, Harrison SM, McGlaughon J, Milko L V., Morales A, Seifert BA, Strande N, Thomson K, Peter Van Tintelen J, Wallace K, Walsh R, Wells Q, Whiffin N, Witkowski L, Semsarian C, Ware JS, Hershberger RE, Funke B. Evaluating the Clinical Validity of Hypertrophic Cardiomyopathy Genes. *Circ Genomic Precis Med* 2019;**12**:57–64.

67.	Jordan E, Peterson L, Ai T, Asatryan B, Bronicki L, Brown E, Celeghin R, Edwards M, Fan J, Ingles J, James CA, Jarinova O, Johnson R, Judge DP, Lahrouchi N, Lekanne Deprez RH, Lumbers RT, Mazzarotto F, Medeiros Domingo A, Miller RL, Morales A, Murray B, Peters S, Pilichou K, Protonotarios A, Semsarian C, Shah P, Syrris P, Thaxton C, Tintelen JP van, et al. Evidence-Based Assessment of Genes in Dilated Cardiomyopathy. *Circulation* 2021;**144**:7–19.

68.	Nagy E, Maquat LE. A rule for termination-codon position within intron-containing genes: when nonsense affects RNA abundance. *Trends Biochem Sci* England; 1998;**23**:198–199.

69.	Kent WJ, Sugnet CW, Furey TS, Roskin KM, Pringle TH, Zahler AM, Haussler a. D. The Human Genome Browser at UCSC. *Genome Res* 2002;**12**:996–1006.

70.	Whiffin N, Walsh R, Govind R, Edwards M, Ahmad M, Zhang X, Tayal U, Buchan R, Midwinter W, Wilk AE, Najgebauer H, Francis C, Wilkinson S, Monk T, Brett L, O’Regan DP, Prasad SK, Morris-Rosendahl DJ, Barton PJR, Edwards E, Ware JS, Cook SA. CardioClassifier: disease- and gene-specific computational decision support for clinical genome interpretation. *Genet Med* 2018;**20**:1246–1254.

71.	CRAN - Package ratesci.

72.	Noether GE. Two Confidence Intervals for the Ratio of Two Probabilities and Some Measures of Effectiveness. *J Am Stat Assoc* Taylor & Francis; 1957;**52**:36–45.

73.	Laud PJ. Equal-tailed confidence intervals for comparison of rates. *Pharm Stat* England; 2017;**16**:334–348.

74.	Donner A, Zou GY. Closed-form confidence intervals for functions of the normal mean and standard deviation. *Stat Methods Med Res* England; 2012;**21**:347–359.

75.	Li Y, Koval JJ, Donner A, Zou GY. Interval estimation for the area under the receiver operating characteristic curve when data are subject to error. *Stat Med* England; 2010;**29**:2521–2531.

76.	Laud P, Dane A. Confidence intervals for the difference between independent binomial proportions: comparison using a graphical approach and moving averages. *Pharm Stat* 2014;**13 5**:294–308.

77.	Gart JJ, Nam J. Approximate interval estimation of the ratio of binomial parameters: a review and corrections for skewness. *Biometrics* United States; 1988;**44**:323–338.

78.	Ng CT, Chee TS, Ling LF, Lee YP, Ching CK, Chua TSJ, Cheok C, Ong HY. Prevalence of hypertrophic cardiomyopathy on an electrocardiogram-based pre-participation screening programme in a young male South-East Asian population: Results from the Singapore Armed Forces Electrocardiogram and Echocardiogram screening protocol. *Europace* 2011;**13**:883–888.

79.	Corrado D, Basso C, Schiavon M, Thiene G. Screening for hypertrophic cardiomyopathy in young athletes. *N Engl J Med* United States; 1998;**339**:364–369.
